# Metabolomic Age (MileAge) predicts health and lifespan: a comparison of multiple machine learning algorithms

**DOI:** 10.1101/2024.02.10.24302617

**Authors:** Julian Mutz, Raquel Iniesta, Cathryn M Lewis

## Abstract

**Background:** Molecular ageing clocks estimate an individual’s biological age. Our aim was to compare multiple machine learning algorithms for developing ageing clocks from nuclear magnetic resonance (NMR) spectroscopy metabolomics data. To validate how well each ageing clock predicted age-related morbidity and lifespan, we assessed their associations with multiple health indicators (e.g., telomere length and frailty) and all-cause mortality.

**Methods:** The UK Biobank is a multicentre observational health study of middle-aged and older adults. The Nightingale Health platform was used to quantify 168 circulating plasma metabolites at the baseline assessment from 2006 to 2010. We trained and internally validated 17 machine learning algorithms including regularised regression, kernel-based methods and ensembles. Metabolomic age (MileAge) delta was defined as the difference between predicted and chronological age.

**Results:** The sample included 101,359 participants (mean age = 56.53 years, SD = 8.10). Most metabolite levels varied by chronological age. The nested cross-validation mean absolute error (MAE) ranged from 5.31 to 6.36 years. 31.76% of participants had an age-bias adjusted MileAge more than one standard deviation (3.75 years) above or below the mean. A Cubist rule-based regression model overall performed best at predicting health outcomes. The all-cause mortality hazard ratio (HR) comparing individuals with a MileAge delta more than one standard deviation above and below the mean was HR = 1.52 (95% CI 1.41-1.64, *p* < 0.001) over a median follow-up of 13.87 years. Individuals with an older MileAge were frailer, had shorter telomeres, were more likely to have a chronic illness and rated their health worse.

**Conclusions:** Metabolomic ageing clocks derived from multiple machine learning algorithms were robustly associated with health indicators and mortality. Our metabolomic ageing clock (MileAge) derived from a Cubist rule-based regression model can be incorporated in research, and may find applications in health assessments, risk stratification and proactive health tracking.

## Introduction

Chronological age, the time elapsed since birth, is a powerful predictor of health and disease (Mutz, Roscoe, & Lewis, 2021). However, there is considerable heterogeneity in health status, lifestyle and the physical signs of ageing between individuals of the same chronological age. This variability may partly reflect individual differences in biological ageing, which is the process of accumulating molecular and cellular damage that results in a progressive decline in physiological functioning (Moqri et al., 2023). While our chronological age cannot be altered, biological ageing trajectories in humans may be modifiable, or even reversible. Therefore, developing reliable measures of biological age is an important priority in biomedical research and population health.

Although there is no single biological marker of biological ageing, several hallmarks such as telomere length shortening have been identified (López-Otín, Blasco, Partridge, Serrano, & Kroemer, 2023). Clinical and population studies of age-related biological changes have also examined physiological measures of grip strength and cardiovascular function (Mutz, Hoppen, Fabbri, & Lewis, 2022; Mutz & Lewis, 2021; Mutz, Young, & Lewis, 2022), blood- based biomarkers (Nakamura, Miyao, & Ozeki, 1988), inflammatory markers (Franceschi, Garagnani, Parini, Giuliani, & Santoro, 2018) and frailty (Hoogendijk et al., 2019).

Molecular “omics” and neuroimaging data such as DNA methylation (Hannum et al., 2013; Horvath & Raj, 2018; Lu et al., 2019) and structural magnetic resonance imaging (Cole & Franke, 2017) have facilitated the development of biological ageing clocks (Rutledge, Oh, & Wyss-Coray, 2022; Solovev, Shaposhnikov, & Moskalev, 2020). Ageing clocks are usually developed using machine learning algorithms that identify relationships between chronological age and molecular data. The difference between predicted age, which approximates biological age, and chronological age is associated with health outcomes (Macdonald-Dunlop et al., 2022). Ageing clocks provide a more holistic picture of a person’s health and are conceptually easier to understand than most individual molecular markers as they are expressed in unit of years.

Population-scale metabolomics, the study of small molecules, i.e., metabolites, within cells, tissues or organisms, is increasingly incorporated into biological ageing research (Panyard, Yu, & Snyder, 2022). Metabolites are the products of metabolism, for example when food is converted to energy. While many initial metabolomics studies were limited to few metabolites and small samples, technological advancements have enabled the population- scale profiling of multiple molecular pathways (Soininen, Kangas, Würtz, Suna, & Ala- Korpela, 2015). Simultaneously quantifying hundreds or thousands of metabolites can provide unprecedented snapshots of an individual’s physiological state. Metabolomic profiles predict many common incident diseases (Buergel et al., 2022) and mortality risk (Deelen et al., 2019). Over the past decade, studies have characterised associations between chronological age and metabolomic biomarkers (Lawton et al., 2008; Menni et al., 2013; Yu et al., 2012). The first study to develop a “metabolite-derived age variable” showed that a panel of 22 metabolites explained 59% of the variance in chronological age. A linear combination of these metabolites was correlated with age-related clinical measures independent of chronological age (Menni et al., 2013). The first study to develop a biological ageing clock from metabolomics data showed that the difference between predicted and chronological age, metabolomic age delta, was associated with a higher disease burden and higher mortality (Hertel et al., 2016). Analyses in other samples, for example the Airwave Health Monitoring Study in the UK (Robinson et al., 2020) and the Dutch BBMRI-NL consortium (van den Akker et al., 2020), have since replicated some of these findings.

Metabolomics is amongst the most powerful omics data for biological age estimation (Solovev et al., 2020) and prediction of disease (Macdonald-Dunlop et al., 2022).

The aim of this study was to compare multiple machine learning algorithms for developing ageing clocks from nuclear magnetic resonance (NMR) spectroscopy metabolomics data in more than 100,000 participants in the UK Biobank (Bycroft et al., 2018). These data provide an unprecedented resource to develop ageing clocks and represent one of the largest single NMR metabolomics databases to date. To validate how well each ageing clock predicted age- related morbidity and lifespan, and captured biological signal beyond that approximated by chronological age (Hertel et al., 2019), we assessed their associations with multiple health indicators (e.g., telomere length and frailty) and all-cause mortality.

## Methods

### Study population

The UK Biobank is a prospective health study of over 500,000 UK residents aged 37–73 who were recruited between 2006 and 2010. Individuals registered with the UK National Health Service (NHS) and living within a 25-mile (∼40 km) radius of one of 22 assessment centres were invited to participate (Bycroft et al., 2018). Participants provided data on their sociodemographic characteristics, health behaviours and medical history, underwent physical examination and had blood and urine samples taken. There is extensive record linkage, for example with national death registries, hospital inpatient records and primary care data.

### Metabolomic biomarker quantification

Nuclear magnetic resonance (NMR) spectroscopy metabolomic biomarkers were quantified in non-fasting blood plasma samples taken at the baseline assessment. The Nightingale Health platform ascertains 168 circulating metabolites using a high-throughput standardized protocol for sample quality control, preparation, data storage and automated analyses (Würtz et al., 2017). The metabolites span multiple pathways, including lipoprotein lipids in 14 subclasses, circulating fatty acids and fatty acid compositions, as well as low-molecular weight metabolites, such as amino acids, ketone bodies and glycolysis metabolites. Most measures are highly correlated (*r* > 0.9) with routine clinical chemistry assays (Würtz et al., 2017). For further details on sample preparation and quality control procedures, see https://biobank.ndph.ox.ac.uk/ukb/ukb/docs/nmrm_companion_doc.pdf. We used the first release of metabolomics data (March 2021) on a random subset of 118,019 participants.

### Machine learning

We evaluated 17 machine learning algorithms, including regularised linear regression, latent variable modelling, instance-based learning, non-parametric regression, kernel-based methods, tree-based models, rule-based models and ensemble methods (Panel 1). To internally validate each algorithm in predicting chronological age from plasma metabolites, we implemented 10×5 nested cross-validation (Figure 1a). Nested cross-validation is preferred for internal validation over other existing approaches as it provides more accurate error estimation (Bates, Hastie, & Tibshirani). We split the data into 10 folds of equal size, to which individuals were allocated at random while preserving the chronological age distribution of the full analytical sample (Figure 1b).

**Figure 1.**
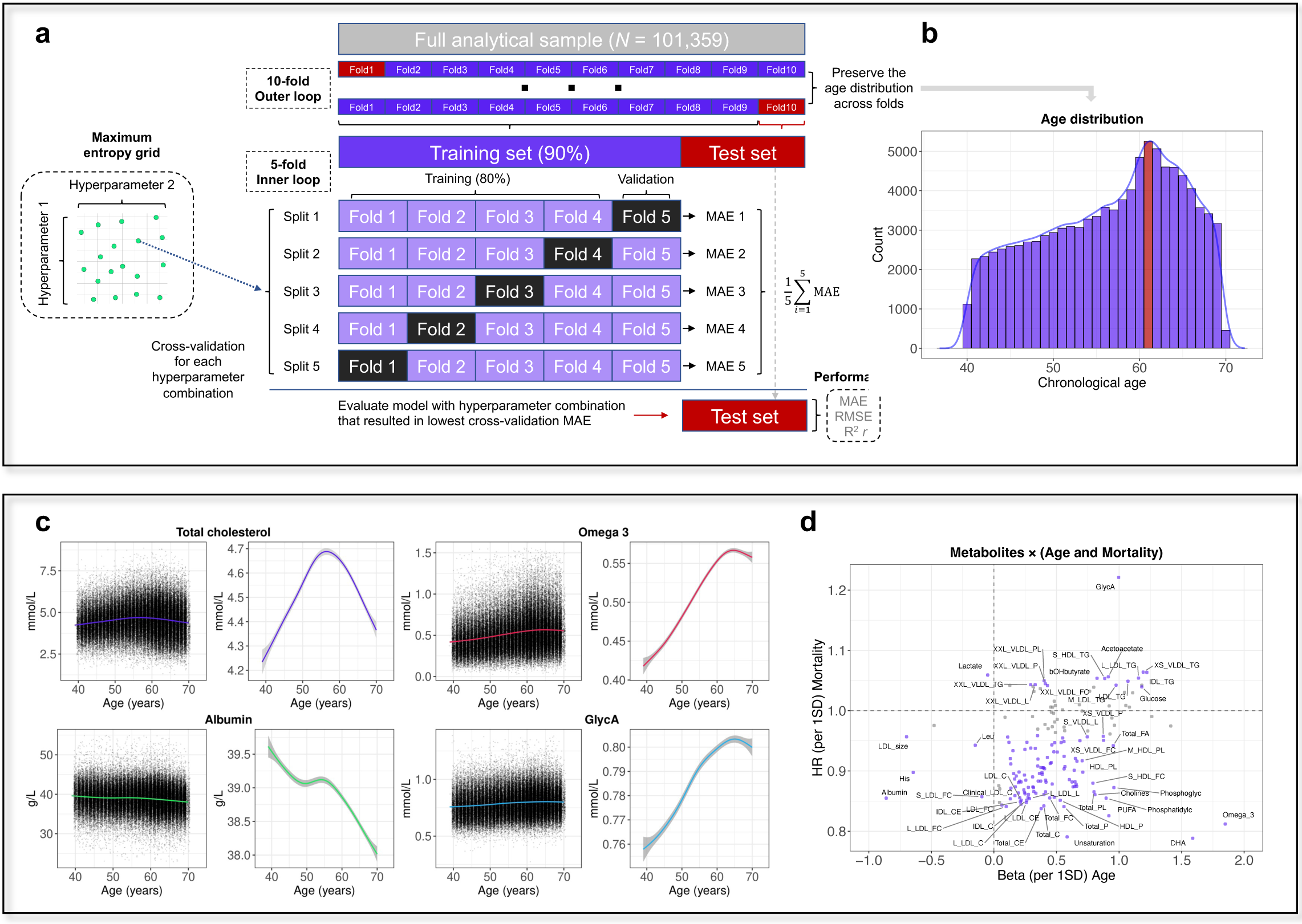
a, Overview of the nested cross-validation approach. MAE = mean absolute error; RMSE = root-mean-square error. **b,** Histogram of the chronological age distribution of the full analytical sample. The statistical mode (age = 61 years) is shown in red. **c,** Distribution of metabolite levels by chronological age, showing scatter plots of all observations and smooth curves (note the difference in y-axis scale). The smooth curves were estimated using generalised additive models, with shaded areas corresponding to 95% confidence intervals. GlycA = glycoprotein acetyls. **d**, Scatter plot showing the hazard ratio (HR) for all-cause mortality and the beta for chronological age associated with a one standard deviation (SD) difference in metabolite levels. Metabolites that had statistically significant associations with both chronological age and all-cause mortality are shown in purple.

**Panel 1.** Overview of the machine learning algorithms used in this study.

Ridge regression: linear regression model with a penalty term (L_2_ regularization) to shrink the magnitude of coefficient estimates towards zero (Hoerl & Kennard, 1970).

<u>Least Absolute Shrinkage and Selection Operator (LASSO)</u>: linear regression model with a penalty term (L_1_ regularization) to shrink the magnitude of coefficient estimates towards zero. This technique can result in sparse models as variable selection is performed by reducing some coefficient estimates exactly equal to zero (Tibshirani, 1996).

<u>Elastic net</u>: linear regression model that combines the L_1_ and L_2_ penalty terms of LASSO and Ridge regression. This technique reduces the magnitude of some coefficient estimates towards zero and can perform variable selection by reducing some coefficient estimates exactly equal to zero (Zou & Hastie, 2005).

<u>Partial least squares regression (PLSR)</u>: latent variable model that extracts a set of latent factors that best explain the covariance between the predictors and outcome. These factors are then used as predictors in a linear regression model (Wold, Sjöström, & Eriksson, 2001).

<u>K-nearest neighbors (KNN)</u>: instance-based learning model which uses the weighted average outcome of the *k* nearest data points to make predictions. In this context, “nearest” is usually determined by a distance metric such as the Minkowski distance (Cover & Hart, 1967).

<u>Multivariate adaptive regression splines (MARS)</u>: non-parametric regression model that creates piecewise linear approximations of the relationship between the predictors and outcome. This technique can model non-linear associations and interactions between the predictors (J. Friedman, H., 1991).

MARS ensemble: ensemble method that combines the predictions of multiple MARS models to improve the predictive accuracy and stability of the model (Kuhn & Johnson, 2013a).

<u>Support vector regression (SVR)</u>: kernel-based method that seeks to identify a hyperplane that best models the relationships between the predictors and outcome. This variation of the support vector machine algorithm (Boser, Guyon, & Vapnik, 1992) was adapted for regression and can employ a range of kernels to transform the data to a higher-dimensional space, allowing for complex non-linear associations (Drucker, Burges, Kaufman, Smola, & Vapnik, 1996). We tested linear, polynomial and radial basis function kernels.

<u>Regression tree</u>: technique that models the relationship between the predictors and outcome by creating a tree-like structure of decision rules based on values of the predictors. Decision trees are interpretable and can incorporate non-linear relationships and higher-order interactions in the data (Breiman, Friedman, Olshen, & Stone, 1984).

<u>Bagging</u>: ensemble method that combines the predictions of multiple regression trees. Bootstrapped aggregating (“bagging”) involves training regression trees on multiple, randomly selected (“bootstrapped”) samples of the data. Bagging can improve the stability and accuracy of decision tree models (Breiman, 1996).

<u>Random forest</u>: ensemble method that combines predictions of multiple regression trees. Each tree is trained on a random subset of the data and, at each split, a subset of predictors, reducing the correlation between decision trees in the ensemble (Breiman, 2001).

Extreme gradient boosting (XGBoost): ensemble method that builds decision trees sequentially by implementing a gradient descent algorithm that seeks to minimize errors from previous models while increasing the influence (“boosting”) of highly predictive models.

More complex models are penalised through L_1_ and L_2_ regularization to avoid overfitting (T. Chen & Guestrin, 2016).

<u>Bayesian additive regression trees (BART)</u>: ensemble method that uses Bayesian techniques to iteratively construct and update multiple decision trees. Regularization priors force each tree to explain only a subset of the relationship between the predictors and outcome, thereby preventing overfitting (Chipman, George, & McCulloch, 2010).

<u>Cubist rule-based regression</u>: ensemble model that that derives rules from decision trees and fits linear regression models for the subset of the data defined by each rule. The model incorporates boosting techniques and may adjust predictions based on *k*-nearest neighbors (Kuhn & Johnson, 2013b; Quinlan, 1992).

<u>RuleFit ensemble</u>: ensemble method that uses a tree-based model (XGBoost) to predict an outcome and subsequently derives rules. LASSO is then used to select the most predictive rules, resulting in a sparse linear model (J. H. Friedman & Popescu, 2008).

For each iteration of the outer loop of the nested cross-validation, 9/10 folds combined served as the training set and the tenth fold served as the test set. The 90% training sets were further divided into five equal size sets, and we performed 5-fold cross validation to empirically identify, for each algorithm, the hyperparameter combination that resulted in the lowest cross-validation mean absolute error (MAE). Tuning grids were set up using a maximum entropy space-filling design. The size of each tuning grid was determined by the number of available hyperparameters, type of hyperparameter (continuous, discrete or categorical) and computational constraints. We tested up to ten values for each hyperparameter, resulting in tuning grid sizes between ten (for Ridge regression) and 3125 (for XGBoost). Further details, including pre-processing requirements, are available in Table S1. The model specifications with the lowest 5-fold cross-validation MAE were subsequently fit in the 90% training sets, and performance was assessed by calculating the MAE, root-mean-square error (RMSE), Pearson correlation coefficient (*r*) and the coefficient of determination (*R^2^*) in the 10% test sets. We also examined the average magnitude of discrepancy in predictive performance between the training and test sets, extrapolation beyond the chronological age range in the data and the computing hours required for hyperparameter tuning for each model.

### Metabolomic ageing clocks

Individual-level age predictions for all participants were obtained by aggregating the predictions of the ten test sets of the outer loop of the nested cross-validation. Metabolomic age delta (MileAge delta) was calculated as the difference between predicted and chronological age, with positive values representing an older predicted than chronological age and negative values representing a younger predicted than chronological age. Given that ageing clocks overestimate age in young individuals and underestimate age in older individuals, we regressed predicted age (MileAge) on chronological age and used the resulting intercept (β) and slope coefficient (α) estimates to apply a statistical correction to the age prediction: MileAge (age bias adjusted) = MileAge + [Age - (α × Age + β)J (de Lange & Cole, 2020).

### Health indicators and mortality

We tested associations between MileAge delta (adj.) and multiple health indicators: having a long-standing illness, disability or infirmity (yes/no), self-rated health (“poor”, “fair”, “good” or “excellent”) and overall health status (unhealthy/healthy) derived from 81 cancer and 443 non-cancer illnesses (Mutz & Lewis, 2022; Mutz et al., 2021). Next we examined associations with the frailty phenotype and frailty index (Mutz, Choudhury, Zhao, & Dregan, 2022). The frailty phenotype summarises data on weight loss, exhaustion, physical activity, walking speed and hand-grip strength. The frailty index was derived from 49 variables obtained at the baseline assessment, including cardiometabolic, cranial, immunological, musculoskeletal, respiratory and sensory traits, well-being, infirmity, cancer and pain. We also tested associations between MileAge delta (adj.) and telomere length, measured using a validated quantitative polymerase chain reaction assay that expresses telomere length as the ratio of the telomere repeat copy number (T) relative to a single-copy gene (S) that encodes haemoglobin subunit beta. T/S ratio is proportional to an individual’s average telomere length (Lai, Wright, & Shay, 2018). Finally, we examined prospective associations with all-cause mortality. The date of death was obtained through linkage with national death registries, NHS Digital (England and Wales) and the NHS Central Register (Scotland). The censoring date was 30 November 2022.

### Exclusion criteria

Women who were pregnant or unsure that they were pregnant at the time of assessment were excluded from the analysis given that their metabolite profiles likely changed during pregnancy. Participants for whom their genetic and self-reported sex did not match were also excluded as this may indicate poor data quality. We also excluded individuals with missing metabolite data or potential outlier metabolite values, defined as values 4× the interquartile range (IQR) above or below the median.

### Statistical analyses

All data processing and analyses were performed in R (version 4.2).

Sample characteristics were summarised using means and standard deviations or counts and percentages. Generalised additive models were used to explore the relationship between chronological age and metabolite levels. We further conducted metabolome-wide association analyses of chronological age and all-cause mortality to identify metabolites that were statistically significantly associated with chronological age and mortality (at *P* < 0.05/168). Correlations between the predicted age derived from each machine learning model were estimated using Pearson’s correlation coefficient.

Cross-sectional associations between MileAge delta (adj.) and the frailty index and telomere length were estimated using ordinary least squares regression. Associations between MileAge delta and having a long-standing illness and overall health status were estimated using logistic regression. Association between MileAge delta and the frailty phenotype and self-rated health were estimated using ordinal logistic regression. For each health indicator, higher values corresponded to worse health. For the health association analyses, we fitted minimally adjusted models that included chronological age and sex as covariates. For the prospective analyses of all-cause mortality, we calculated person-years of follow-up and the median duration of follow-up of censored individuals. Survival probabilities by MileAge delta were estimated using the Kaplan-Meier (KM) method (Kaplan & Meier, 1958) and we calculated log-rank *p*-values. Hazard ratios (HRs) and 95% confidence intervals were estimated using Cox proportional hazards models (Cox, 1972). Age in years was used as the underlying time axis, with age 40 as the start of follow-up. Across both cross-sectional and prospective analyses, we defined MileAge delta subgroups by standard deviation from the mean.

Individuals with a MileAge delta equal to or smaller than one standard deviation below the mean were the reference group. To discern how this analytical decision might impact results, and to enable comparability with other studies, we also report associations with all-cause mortality for all models with subgroups defined by the bottom and top 10% of the distribution as well as negative and positive values. Finally, we used generalised additive models and spline functions to explore the relationship between MileAge delta as a continuous variable and health indicators and all-cause mortality, respectively.

For the Cubist rule-based regression model, which across most analyses performed best at predicting health outcomes, we performed additional analyses. We calculated variable importance scores to identify metabolites that strongly contributed to MileAge. We explored associations between MileAge delta and all-cause mortality stratified by sex, self-rated health and chronological age group (39-49, 50-59 and 60-71 years). Finally, we assessed the performance of our ageing clock by benchmarking it against other ageing markers: (a) we estimated the hazard ratio for all-cause mortality by MileAge delta and other ageing marker (grip strength, telomere length and the frailty index) subgroups defined by standard deviation from the mean, adjusted for chronological age and sex; (b) we calculated the C-index and 95% confidence intervals for chronological age + sex (as the base model) and for each ageing marker added separately to the model, with time (in days) since the baseline assessment as the underlying time axis.

## Results

### Sample characteristics

Of the 118,019 participants with metabolomics data, 110,730 had complete information on all metabolites (Figure S1). After removing individuals with potential outlier metabolite values, inconsistencies between self-reported and genetic sex or possible pregnancies, the analytical sample included 101,359 participants (Table 1). The mean chronological age was 56.44 years (SD = 8.12), with the most common age being 61 years (Figure 1b). Most metabolite levels varied by chronological age (Figure 1c), showing considerable evidence of non-linear relationships (Figures S2-S33).

**Table 1.** Sample characteristics.

|  | MileAge delta (adj.) |  |  |  |
| --- | --- | --- | --- | --- |
|  | Full sample<br>(N=101359) | ≤ 1SD below<br>the mean<br>(n=16204) | Middle<br>(n=69166) | ≥ 1SD above<br>the mean<br>(n=15989) |
| Age; mean (SD) | 56.44 (8.12) | 55.85 (8.33) | 56.75 (8.08) | 55.69 (8.00) |
| Sex |  |  |  |  |
| Female | 54484 (53.8%) | 8062 (49.8%) | 37002 (53.5%) | 9420 (58.9%) |
| Male | 46875 (46.2%) | 8142 (50.2%) | 32164 (46.5%) | 6569 (41.1%) |
| Ethnicity |  |  |  |  |
| White | 95672 (94.4%) | 15163 (93.6%) | 65486 (94.7%) | 15023 (94.0%) |
| Mixed-race | 574 (0.6%) | 103 (0.6%) | 386 (0.6%) | 85 (0.5%) |
| Black | 1529 (1.5%) | 233 (1.4%) | 975 (1.4%) | 321 (2.0%) |
| Asian | 1953 (1.9%) | 375 (2.3%) | 1265 (1.8%) | 313 (2.0%) |
| Chinese | 291 (0.3%) | 77 (0.5%) | 184 (0.3%) | 30 (0.2%) |
| Other | 880 (0.9%) | 173 (1.1%) | 564 (0.8%) | 143 (0.9%) |
| Missing<sup>1</sup> | 460 (0.5%) | 80 (0.5%) | 306 (0.4%) | 74 (0.5%) |
| Highest qualification |  |  |  |  |
| None | 17084 (16.9%) | 2551 (15.7%) | 11868 (17.2%) | 2665 (16.7%) |
| O levels/GCSEs/CSEs | 27008 (26.6%) | 4237 (26.1%) | 18435 (26.7%) | 4336 (27.1%) |
| A levels/NVQ/HND/HNC<sup>2</sup> | 23235 (22.9%) | 3637 (22.4%) | 15961 (23.1%) | 3637 (22.7%) |
| Degree | 32826 (32.4%) | 5577 (34.4%) | 22091 (31.9%) | 5158 (32.3%) |
| Missing<sup>1</sup> | 1206 (1.2%) | 202 (1.2%) | 811 (1.2%) | 193 (1.2%) |
| Household income<sup>3</sup> |  |  |  |  |
| Very low | 19422 (19.2%) | 2863 (17.7%) | 13265 (19.2%) | 3294 (20.6%) |
| Low | 22159 (21.9%) | 3487 (21.5%) | 15137 (21.9%) | 3535 (22.1%) |
| Medium | 22634 (22.3%) | 3831 (23.6%) | 15417 (22.3%) | 3386 (21.2%) |
| High | 17767 (17.5%) | 3095 (19.1%) | 12027 (17.4%) | 2645 (16.5%) |
| Very high | 4725 (4.7%) | 819 (5.1%) | 3188 (4.6%) | 718 (4.5%) |
| Missing<sup>1</sup> | 14652 (14.5%) | 2109 (13.0%) | 10132 (14.6%) | 2411 (15.1%) |
| Cohabitation |  |  |  |  |
| With partner | 73896 (72.9%) | 12054 (74.4%) | 50748 (73.4%) | 11094 (69.4%) |
| Single | 8341 (8.2%) | 1418 (8.8%) | 5467 (7.9%) | 1456 (9.1%) |
| Missing<sup>1</sup> | 19122 (18.9%) | 2732 (16.9%) | 12951 (18.7%) | 3439 (21.5%) |
| Smoking status |  |  |  |  |
| Never | 55643 (54.9%) | 9149 (56.5%) | 37876 (54.8%) | 8618 (53.9%) |
| Former | 34805 (34.3%) | 5342 (33.0%) | 23838 (34.5%) | 5625 (35.2%) |
| Current | 10416 (10.3%) | 1634 (10.1%) | 7119 (10.3%) | 1663 (10.4%) |
| Missing<sup>1</sup> | 495 (0.5%) | 79 (0.5%) | 333 (0.5%) | 83 (0.5%) |
| Menopause<sup>4</sup> |  |  |  |  |
| No | 12986 (12.8%) | 2887 (17.8%) | 8096 (11.7%) | 2003 (12.5%) |
| Yes | 32745 (32.3%) | 4046 (25.0%) | 23034 (33.3%) | 5665 (35.4%) |
| Missing<sup>1</sup> | 55628 (54.9%) | 9271 (57.2%) | 38036 (55.0%) | 8321 (52.0%) |
| Morbidity count |  |  |  |  |
| None | 23788 (23.5%) | 4459 (27.5%) | 16311 (23.6%) | 3018 (18.9%) |
| One | 26923 (26.6%) | 4581 (28.3%) | 18615 (26.9%) | 3727 (23.3%) |
| Two | 20535 (20.3%) | 3195 (19.7%) | 14011 (20.3%) | 3329 (20.8%) |
| Three | 13157 (13.0%) | 1857 (11.5%) | 9001 (13.0%) | 2299 (14.4%) |
| Four | 7553 (7.5%) | 989 (6.1%) | 5077 (7.3%) | 1487 (9.3%) |
| Five+ | 9387 (9.3%) | 1119 (6.9%) | 6141 (8.9%) | 2127 (13.3%) |
| Missing<sup>1</sup> | 16 (0.0%) | 4 (0.0%) | 10 (0.0%) | 2 (0.0%) |
| Fasting time<sup>5</sup>; mean (SD) | 3.75 (2.40) | 3.74 (2.47) | 3.74 (2.39) | 3.79 (2.38) |
| Body mass index<sup>6</sup>; mean (SD) | 27.43 (4.73) | 26.17 (4.07) | 27.41 (4.57) | 28.80 (5.59) |

*Note:* GCSEs = general certificate of secondary education; CSE = certificate of secondary education; NVQ = national vocational qualification; HND = higher national diploma; HNC = higher national certificate. MileAge delta (adj.) derived from the Cubist rule-based regression model. <sup>1</sup>Missing data may also include “do not know”, “prefer not to answer”, “not applicable” or related responses. <sup>2</sup>Also includes ‘other professional qualifications’. <sup>3</sup>Annual household income groups: very low (<£18,000), low (£18,000–£30,999), middle (£31,000–£51,999), high (£52,000–£100,000) and very high (>£100,000). <sup>4</sup>*n* = 59519 females. <sup>5</sup>*n* = 3 missing. <sup>6</sup>*n* = 345 missing.

### Metabolite-wide associations

165/168 metabolites were associated with chronological age (*p* < 0.05/168). While most metabolite levels were elevated in older individuals (e.g., Omega 3, citrate and glucose), seven, including albumin, glycine and histidine, were negatively associated with chronological age (Figure S34; Additional file 1). Amongst the 119 metabolites associated with all-cause mortality, GlycA was most strongly associated with a higher mortality hazard (HR = 1.22, 95% CI 1.20-1.25, *p* < 0.001), whereas the degree of unsaturation, docosahexaenoic acid (DHA) and Omega 3 were strongly associated with a lower mortality hazard (Figure S35; Additional file 2). Notably, 116 metabolites associated with chronological age also predicted mortality, with GlycA, Omega 3 and DHA amongst the most strongly associated metabolites of both age and mortality (Figure 1d). Between 87% and 95% of the metabolites that were statistically significantly associated with other health indicators (e.g., frailty and health status) were associated with chronological age (Figure S36). The exception was telomere length for which only 53% of statistically significant metabolites were shared with chronological age.

### Predictive model performance

Predictive performance estimates in the 90% training (*n* = 91,222 to 91,226) and 10% test sets are shown in Table S3. The nested cross-validation mean absolute error (MAE) in the test sets (*n* = 10,133 to 10,137) ranged from 5.31 to 6.36 years, with the support vector regression with a radial basis function (SVM radial) performing best and the MARS ensemble performing worst. The root-mean-square error (RMSE) ranged from 6.60 to 7.58 years. Correlation coefficients between predicted and chronological age ranged from 0.36 to 0.59, with *R^2^* values between 0.13 and 0.35. The difference in predictive performance between the training and test sets, i.e., the model’s optimism which may indicate poor generalization to unseen data, was generally low (e.g., MAE_difference_ < 0.15 for 10/17 models). However, certain tree-based models (bagging, random forest and XGBoost) and the *k*-nearest neighbors model overfit the training data (MAE_difference_ = -1.51 to -5.83), with correlation coefficients between predicted and chronological age of *r* > 0.8 in the training sets (Figure S40). Figure 2a shows the nested cross-validation MAEs for all models. For comparison, drawing random samples from a uniform distribution between the sample’s minimum and maximum chronological age, i.e., a random prediction model, resulted in a MAE = 9.79.

**Figure 2.**
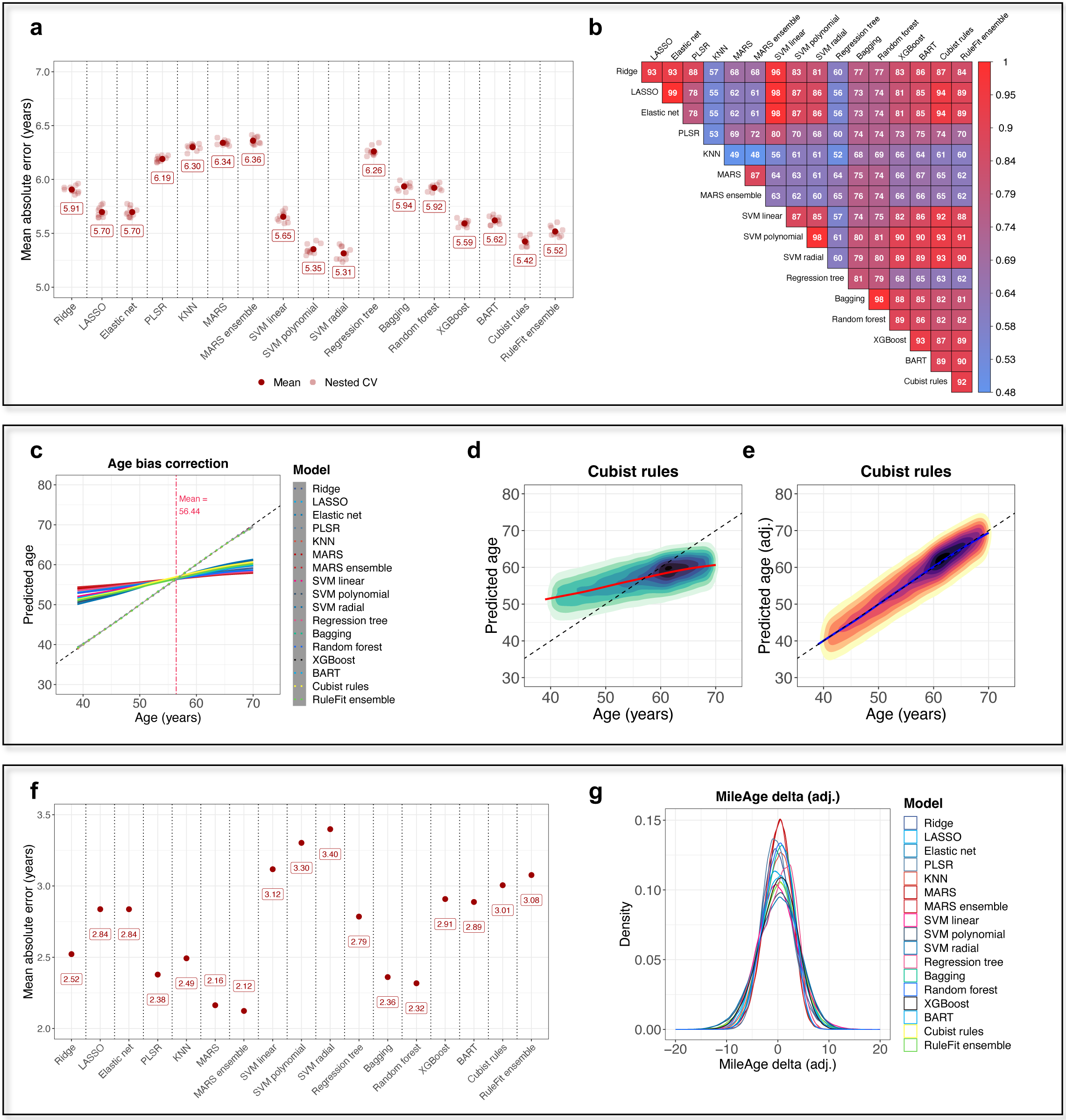
a, Nested cross-validation mean absolute error (MAE) for all models with tuned hyperparameter values in the 10% hold-out test sets. CV = cross-validation. **b,** Heatmap of Pearson’s correlation coefficient (*r*) between the predicted age values for all models. Estimates shown were multiplied by 100. **c,** Line plot showing the correlation between predicted age and chronological age for all models before (solid lines) and after (dotted lines) applying a statistical correction to the predicted age to remove the age bias (i.e., the systematic overestimation of age in young individuals and underestimation of age in older individuals). **d, e,** 2D density plots showing the correlation between predicted age derived from the Cubist rule-based regression model and chronological age before and after age bias correction. Observations beyond y-axis limits of 30 to 80 not shown. **f,** Mean absolute error for all models with tuned hyperparameter values calculated in the full sample after age bias correction. **g,** Density plot showing the distribution of MileAge

Predicting the sample mean, i.e., a null model, resulted in a MAE = 6.96. Additional plots showing other performance measures (MAE, RMSE, *r* and *R^2^*) in the training and test sets are presented in the supplement (Figures S37-S43). There were moderate to high correlations between the predicted age values of the various models (*r* = 0.48 to > 0.99) (Figure 2b). High correlations (*r* > 0.87) amongst the most accurate models suggest they capture similar patterns in the data. Omega 3, albumin and citrate were amongst the most important contributors to predictive accuracy (Figure S44).

### Age bias correction

All models overestimated the age of young individuals and underestimated the age of older individuals (Figure S44). Applying a statistical correction (see Methods) to the predicted age values removed this bias (Figures 2c-e and S45). The predicted age values were originally within the chronological age range (39 to 70 years) of the sample for most individuals.

Across *all* models, 0.23% (*n* = 238) and 0.96% (*n* = 976) of predictions were below or above the minimum and maximum age, respectively (Table S4). More predicted age values were outside the chronological age range (up to 8.27%, *n* = 8382 for a single model) after the age bias correction. Re-calculating the predictive performance estimates after the age bias correction suggested higher accuracy (MAE = 2.12 to 3.40; Figure 2f). The overall performance rankings across the models inverted, with the models that originally predicted chronological age best showing reduced accuracy (Figures S46-S48). The age bias adjusted MileAge delta ranged from -18.94 years younger to 16.05 years older for the Cubist rule-based regression model, with 15.99% (*n* = 16,204) and 15.77% (*n* = 15,989) of the sample having a MileAge delta (adj.) of at least 3.75 years below or above the mean (Figure 2g; Table S5).

### Associations with health indicators

Descriptive statistics for the health indicators are shown in Table S6. Having an older predicted than chronological age was associated with higher frailty index scores across all models (Table S7). This extended to the frailty phenotype for all models when comparing individuals with a MileAge delta (adj.) more than one standard deviation above and below the mean, and for 12/17 models when comparing the middle of the distribution to individuals with MileAge delta (adj.) values more than one standard deviation below the mean (Table S8). For telomere length, we observed a group difference for 12/17 models when comparing the tails of the distribution, and for 9/17 models when comparing the middle of the distribution to the reference group (Table S9). Having an older predicted than chronological age was generally associated with chronic illness and poor self-rated health (Tables S10- S12). An exception to this pattern were the MARS models, for which an older predicted than chronological age was associated with *longer* telomeres, and for which there was little evidence of statistically significant differences in health between individuals with MileAge delta (adj.) values in the middle of the distribution and the reference group.

MileAge delta (adj.) derived from the Cubist rule-based regression model was most strongly associated with most health indicators (Figure 3a). Individuals with a MileAge delta (adj.) greater than one standard deviation above the mean had higher frailty index scores than individuals with a MileAge delta (adj.) smaller than one standard deviation below the mean (β = 0.023, 95% CI 0.021–0.024, *p* < 0.001). This group difference was approximately equivalent to an 18.3-year chronological age difference in frailty index scores (β = 0.023 divided by β = 0.001255 derived from a linear model, y _frailty index_ ∼ x _chronological age_).

**Figure 3.**
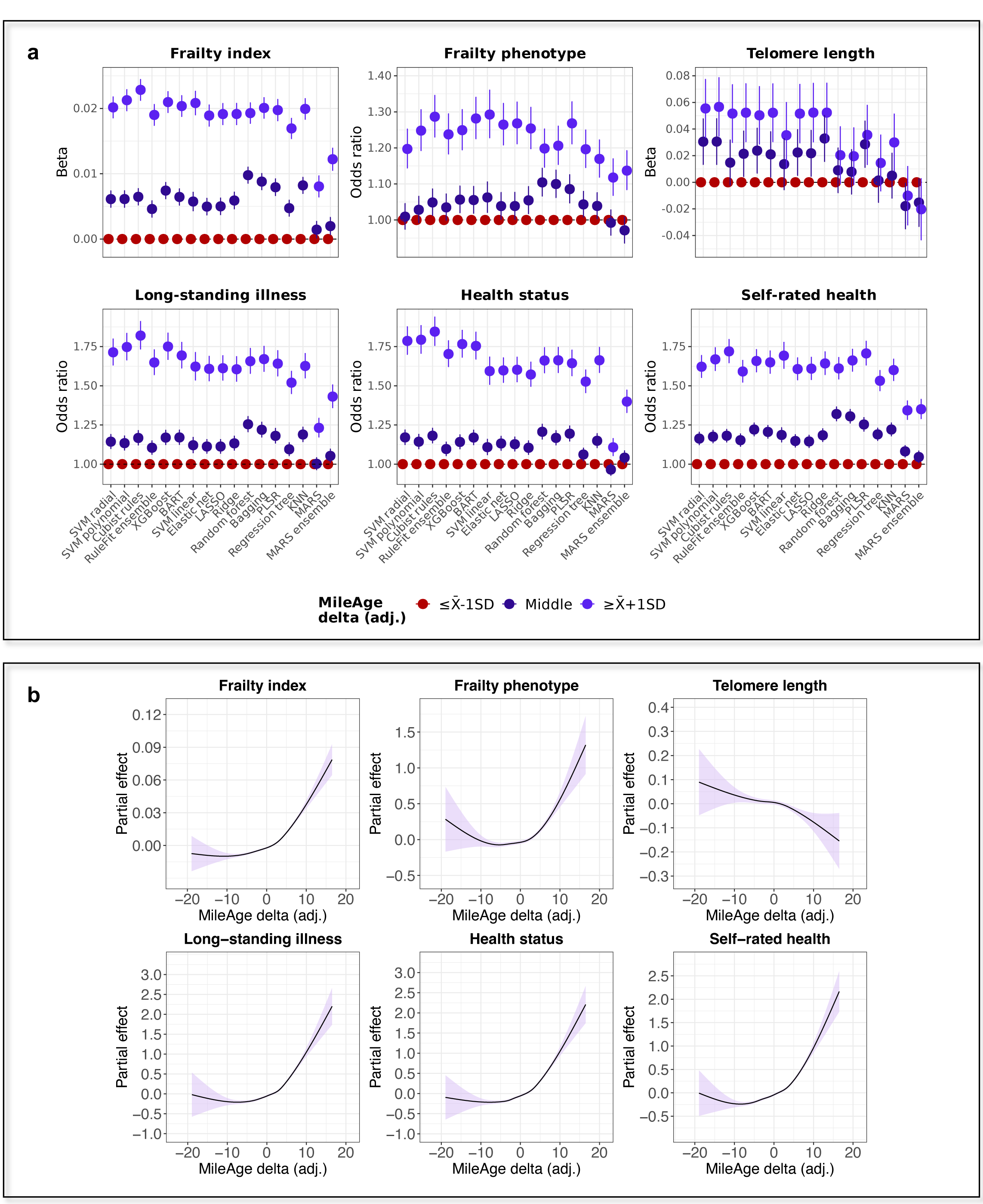
a, Associations between MileAge delta (adj.) and health indicators for all models. Models were adjusted for chronological age and sex. Reference group: individuals with a MileAge delta (adj.) smaller than one standard deviation below the mean. See Panel 1 for model abbreviations. **b,** Partial effect plots of generalised additive models of the association between health indicators and MileAge delta (adj.). Models

Individuals with an older predicted than chronological age were also more likely to be physically frail (OR = 1.29, 95% CI 1.23–1.35, *p* < 0.001) and had shorter telomeres (β = 0.052, 95% CI 0.030 to 0.073, *p* < 0.001) equivalent to a 2.2-year chronological age difference in telomere length. Such individuals were also more likely to have a long-standing illness (OR = 1.82, 95% CI 1.73–1.91, *p* < 0.001), poorer health status (OR = 1.85, 95% CI 1.76–1.94, *p* < 0.001) and worse self-rated health (OR = 1.72, 95% CI 1.65–1.80, *p* < 0.001). Generalised additive models showed that positive MileAge delta (adj.) values, indicating accelerated biological ageing, were robustly associated with unfavourable health (Figure 3b), whereas negative MileAge delta (adj.) values were only weakly associated with favourable health, a pattern that was consistent across most models (Figures S49-S54).

### Predicting mortality

The median duration of follow-up of censored individuals was 13.87 years (IQR = 1.37 years), with 1,361,970 person-years of follow-up. There were 8113 deaths amongst 101,274 participants (*n* = 85 missing). MileAge (adj.) was strongly associated with all-cause mortality, comparable to chronological age (Figures 4a-e). In the prospective analyses we examined the age bias adjusted MileAge delta and models were adjusted for chronological age and sex, with age (in years) as the underlying time axis (Figure S55 and Table S13).

**Figure 4.**
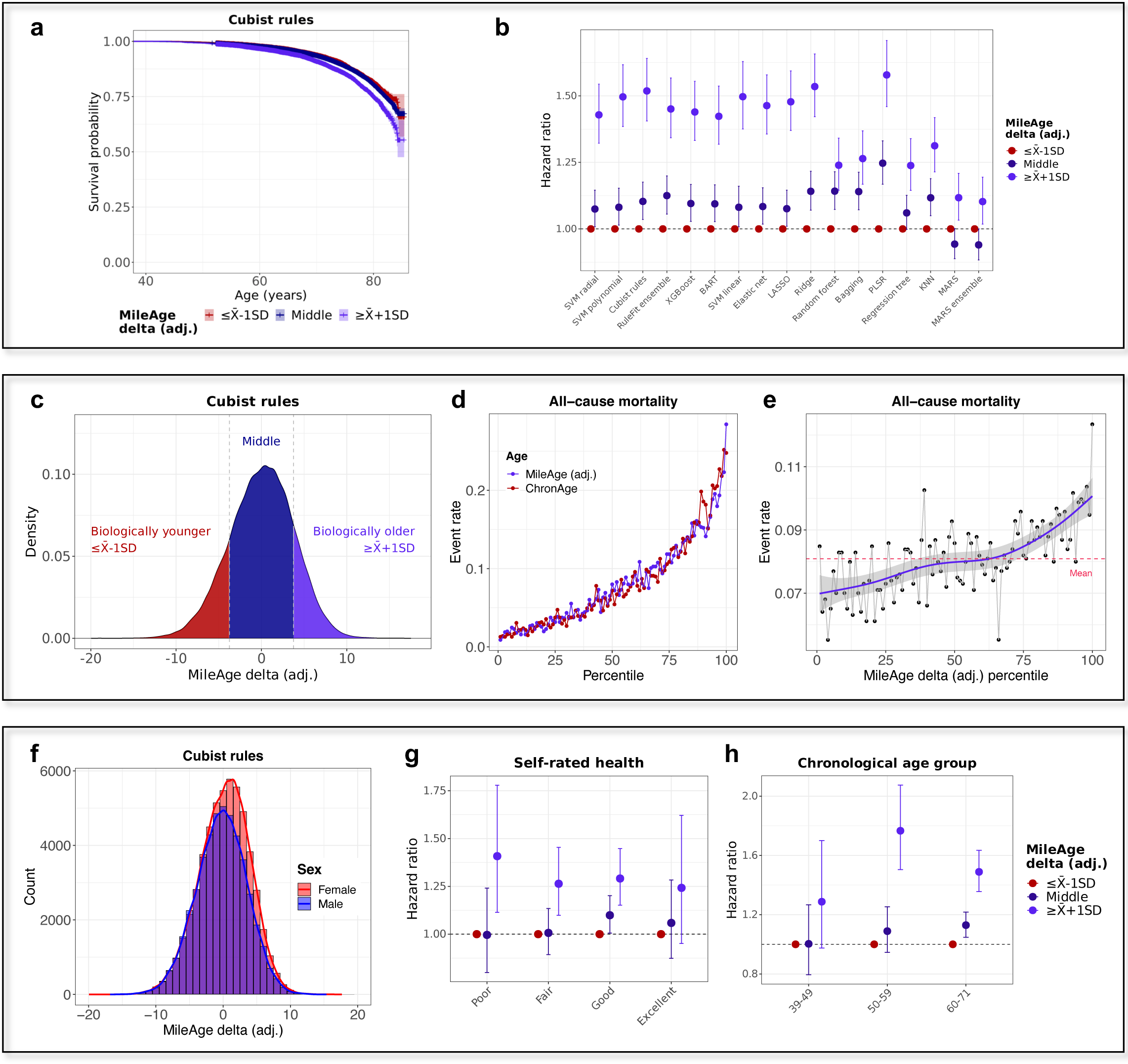
a, Kaplan-Meier plot showing survival probabilities for all-cause mortality by MileAge delta (adj.) derived from the Cubist rule-based regression model. Log-rank test *p*-value < 0.001. **b,** Hazard ratios and 95% confidence intervals from Cox proportional hazards models by MileAge delta (adj.) for all models. Models were adjusted for chronological age and sex. Age (in years) was used as the underlying time axis. Reference group: individuals with a MileAge delta (adj.) smaller than one standard deviation below the mean. See Panel 1 for model abbreviations. **c,** Density plot showing the distribution of MileAge delta (adj.) derived from the Cubist rule-based regression model. **d, e,** All-cause mortality rate by percentile of chronological age, MileAge (adj.) and MileAge delta (adj.) derived from the Cubist rule-based regression model. **f,** Histogram showing the distribution of MileAge delta (adj.) derived from the Cubist rule-based regression model, stratified by sex. **g, h,** Hazard ratios and 95% confidence intervals from Cox proportional hazards models for all-cause mortality by MileAge delta (adj.) derived from the Cubist rule-based regression model, stratified by self-rated health and chronological age group.

For the Cubist rule-based regression model, the hazard ratio (HR) comparing individuals with a MileAge delta (adj.) greater than one standard deviation above and below the mean was HR = 1.52 (95% CI 1.41-1.64, *p* < 0.001). Individuals with a MileAge delta (adj.) between one standard deviation above and below the mean had a statistically significantly higher mortality risk for 14/17 models (*p* between 0.03 and < 0.001) (Table S14). Comparing the bottom and top 10% of the MileAge delta (adj.) distribution, instead of one standard deviation above and below the mean, resulted in greater differences (e.g., HR = 1.63, 95% CI 1.48-1.79, *p* < 0.001 for the Cubist model). Individuals in between the tails of the distribution had a higher mortality risk compared to the bottom 10% for 14/17 models (Figure S56 and Table S15).

When comparing individuals with a positive and negative MileAge delta (adj.), we found that individuals with an older predicted than chronological age had a higher mortality risk for all models except the MARS models (Figure S57 and Table S16). Modelling the mortality hazard as a spline function of MileAge delta (adj.) suggested that positive values were strongly associated with a higher mortality hazard, while there was little evidence that negative values were associated with a lower mortality hazard (Figure S58).

Females had slightly higher MileAge delta (adj.) values than males (Figure 4f), a pattern which we observed across all models (Figure S59). The mortality hazard of individuals with a MileAge delta (adj.) greater than one standard deviation above the mean was elevated in both females (HR = 1.26, 95% CI 1.11-1.43, *p* < 0.001) and males (HR = 1.73, 95% CI 1.57-1.91, *p* < 0.001) (Figures S60-S61; Table S17). The difference in mortality between individuals with a MileAge delta (adj.) in the middle of the distribution and the reference group was only statistically significant in males (HR = 1.14, 95% CI 1.05-1.23, *p* = 0.002). Stratifying the sample by self-rated health, the mortality hazard of individuals with a MileAge delta (adj.) greater than one standard deviation above the mean was higher in all strata (e.g., HR = 1.41, 95% CI 1.11-1.78, *p* < 0.001 for poor self-rated health) except for individuals with excellent health (Figure 4g and Table S17). The mortality hazard of individuals with a MileAge delta (adj.) greater than one standard deviation above the mean was higher in the age groups above 50 years (Figures S62 and 4h; Table S17).

### Comparison with other ageing markers

A comparison of the all-cause mortality hazard associated with MileAge delta (adj.) and other ageing marker subgroups defined by the standard deviation from the mean showed that the largest hazard ratio (HR = 2.90) was observed for the frailty index (Figure 5a). The smallest hazard ratio was observed for telomere length (HR = 1.31) (Table S18), with MileAge delta (adj.) (HR = 1.52) and grip strength (HR = 1.90) in-between. Adding each ageing marker individually as a continuous variable to a base model that included chronological age and sex improved prediction of all-cause mortality, with the best prediction observed for the frailty index (C-index 0.737, 95% CI 0.732 to 0.742 vs C-index 0.716, 95% CI 0.711 to 0.722 for the base model) (Figure 5b and Table S19). Modelling the mortality hazard as a spline function of the ageing markers, to identify potential non-linear effects, suggested that the all-cause mortality hazard was considerably higher in individuals with an older predicted than chronological age. For example, compared to the sample median MileAge delta (adj.), which was equivalent to no difference between predicted and chronological age, a MileAge delta (adj.) of 10 was associated with a HR of about 2.7, i.e., a 170% higher morality hazard (Figure 5c).

**Figure 5.**
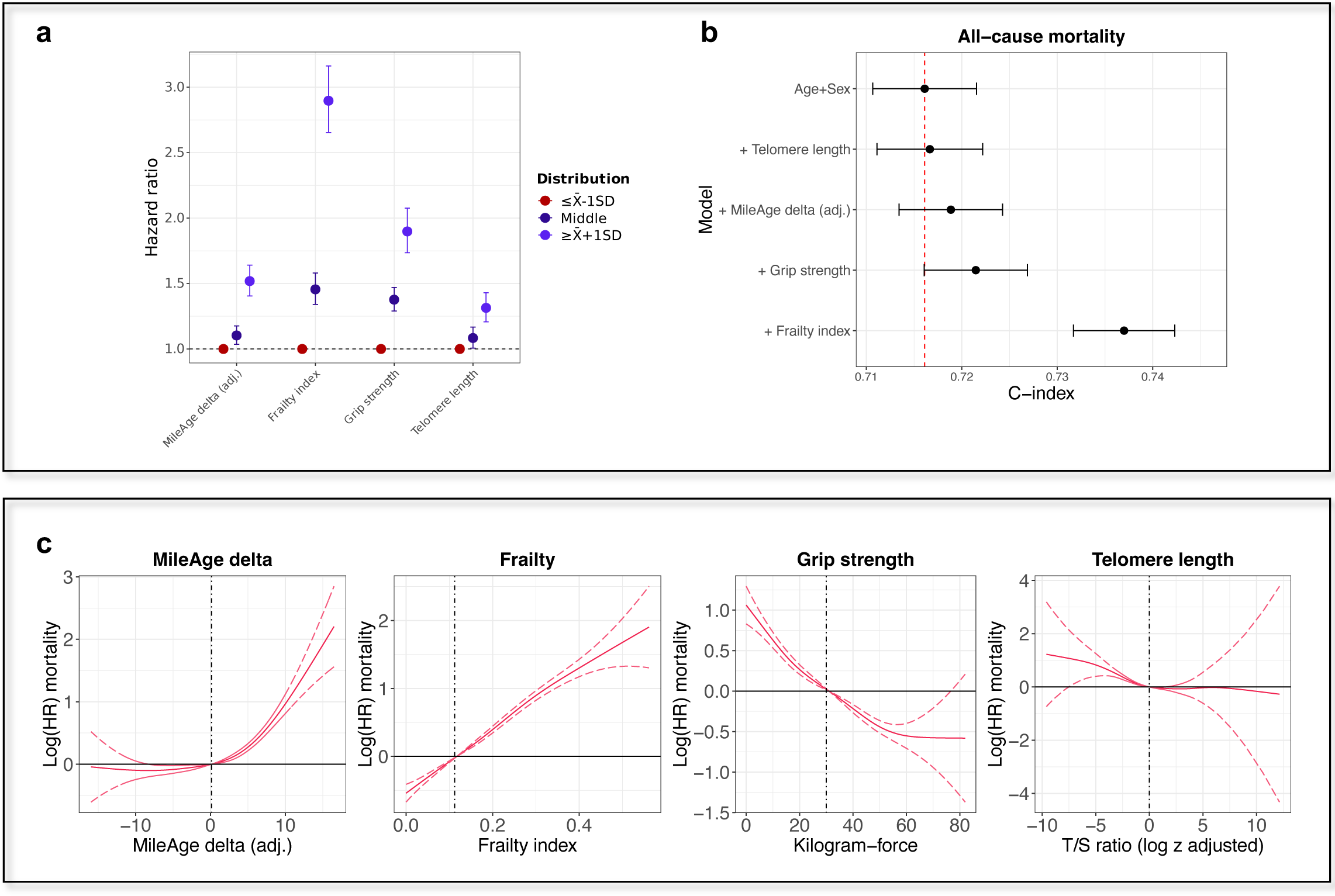
a, Hazard ratios (HR) and 95% confidence intervals from Cox proportional hazards models for all-cause mortality by MileAge delta (adj.), the frailty index, grip strength and telomere length. Models were adjusted for chronological age and sex. Age (in years) was used as the underlying time axis. Reference group: individuals with a score smaller than one standard deviation below the mean. **b,** C-index and 95% confidence intervals from Cox proportional hazards models for all-cause mortality for chronological age + sex (the base model) and for each ageing marker added separately to the base model. Time (in days) was used as the underlying time axis. **c,** Log(HRs) and 95% confidence intervals from Cox proportional hazards models for all-cause mortality. Models were adjusted for chronological age and sex. Age (in years) was used as the underlying time axis. Vertical lines indicate the median of the distribution which represents the reference for interpreting the estimates shown.

## Discussion

In 101,359 UK Biobank participants with Nightingale Health metabolomics data, we observed that most metabolite levels varied by chronological age, with considerable evidence of non-linear associations. Across the machine learning algorithms employed to develop ageing clocks from circulating plasma metabolites, the nested cross-validation mean absolute error between predicted age (MileAge) and chronological age ranged from 5.31 to 6.36 years (*R^2^* between 0.13 and 0.35). All models overestimated age in young individuals and underestimated age in older individuals. After applying a statistical correction to remove this age bias, 31.76% of participants had adjusted MileAge delta values of at least 3.75 years, highlighting the potential of metabolomic ageing clocks to differentiate between individuals of the same chronological age (Hertel et al., 2019). While there was a high degree of consistency across the top performing models such as support vector regression, tree-based and rule-based ensembles, the ageing clock derived from a Cubist rule-based regression model (MAE = 5.42) was most strongly associated with most health indicators. We observed across most models that individuals with an older predicted than chronological age, indicating accelerated biological ageing, were frailer, had shorter telomeres, were more likely to have a chronic illness, rated their health worse and had a higher mortality risk.

Multiple studies have developed biological ageing clocks trained on chronological age from metabolomics data. The seminal study by Menni et al. (2013) derived a metabolite age score as a linear combination of 22 plasma metabolites that were correlated with chronological age in 6055 twins, achieving an *R^2^* of 59% and a hazard ratio for all-cause mortality of HR = 1.08 per year. Hertel et al. (2016) tested a multivariable linear regression and a fractional polynomial model in 3611 participants, with the latter achieving a correlation between predicted and chronological age of *r* = 0.53 (RMSE = 11.19) in men and *r* = 0.61 (RMSE = 10.37) in women. Their metabolomic ageing clock included 59 urinary metabolites and was predictive of all-cause mortality (HR = 1.24 per SD). Robinson et al. (2020) developed several ageing clocks using elastic net models in 2238 participants, with correlations between predicted and chronological age of *r* = 0.45 (MAE = 4.17) to *r* = 0.83 (MAE = 6.49). Akker et al. (2020) developed an ageing clock, MetaboAge, from 56 metabolites in 18,716 participants from 24 community and hospital-based cohorts using a linear regression model, achieving a correlation of *r* = 0.65 and a median absolute error of 7.3. An independent external validation of this clock observed a notably lower correlation of *r* = 0.21 (Macdonald-Dunlop et al., 2022). The same study also developed a clock from NMR metabolomics data (81/86 metabolites selected) in 1643 individuals, which achieved a correlation of *r* = 0.74 but failed to replicate in a validation cohort, and another clock from mass spectrometry metabolomics (181/682 metabolites selected) in 861 individuals, which achieved a correlation of *r* = 0.81 (Macdonald-Dunlop et al., 2022). An updated ageing clock, MetaboAge 2.0, in the BBMRI-nl data that included 65 metabolites achieved an *R^2^* of 0.451 and 0.449 for a linear regression and elastic net model, respectively (Bizzarri et al., 2023).

Metabolomic ageing clocks trained on chronological age are generally less accurate than ageing clocks derived from other types of omics data (Rutledge et al., 2022). Nevertheless, our most predictive model (MAE = 5.31 years) had a similar accuracy as a deep learning ageing clock (MAE = 5.68) derived from blood markers, biometrics and sex in the China Health and Retirement Longitudinal Study. An elastic net model in the same data achieved a MAE = 6.19 years (Galkin et al., 2022), while the elastic net model in our study had a lower MAE = 5.70, likely due to the larger sample size. While a discussion of how well different ageing clocks predict health outcomes is beyond the scope of this study, the most widely tested epigenetic ageing clocks that were trained on chronological age are weak predictors of mortality (B. H. Chen et al., 2016; Fransquet, Wrigglesworth, Woods, Ernst, & Ryan, 2019). Second-generation ageing clocks are more predictive of mortality. For example, a one-year increase in PhenoAge, which was trained on physiological dysregulation, was associated with a 9% increase in mortality risk. Its epigenetic derivation, DNAm PhenoAge, was associated with a 4.5% increase in all-cause mortality (Levine et al., 2018). GrimAge, another popular mortality risk predictor that incorporates chronological age, sex and eight DNA methylation surrogate markers (seven for plasma proteins and one for smoking pack-years) strongly predicts mortality and age-related diseases (Lu et al., 2019). A recent clock developed from circulating biomarkers and trained to predict mortality in the UK Biobank (*n* = 307,000) using an elastic net Cox model (Bortz et al., 2023) yielded a 9.2% relative increase in prediction compared to the PhenoAge model. However, the improvement in prediction over a model that included chronological age and sex was modest (C-index 0.715 vs 0.762, compared to 0.716 vs 0.719 in our study; though note that our clock was not trained to predict mortality). These findings suggest that certain omics clocks capture physiologically relevant signals more, while first-generation epigenetic clocks are specifically good at predicting chronological time (Rutledge et al., 2022).

Although chronological age prediction is valuable in fields such as forensics (Vidaki et al., 2017), it is of limited use in population health and geroscience, given that chronological age is non-modifiable (Robinson & Lau, 2020). A perfect prediction model would merely tell us about chronological, not biological, age (Nakamura et al., 1988). It is the *difference* between predicted and chronological age (MileAge delta) that serves as an indicator of accelerated or decelerated ageing. A less accurate chronological age prediction does not necessarily indicate a worse biological age model (Hertel et al., 2019), hence we also tested algorithms that were expected to perform less well at predicting chronological age (e.g., bagging). Prior studies suggested that biological age estimates derived from physical activity (Pyrkov et al., 2018) or epigenetic data (Zhang et al., 2019) with higher predictive accuracy of chronological age were less predictive of all-cause mortality. Our study showed that the models that were most predictive of chronological age were generally also more strongly associated with health and mortality, though we note that the pattern of predictive accuracy largely inverted after we applied the age bias correction.

We report several additional analyses, including variable importance scores and benchmarking against other ageing markers, for the ageing clock derived from the Cubist rule-based regression model, which was most strongly associated with most health indicators. There are, however, several conclusions that can be drawn across most of the machine learning algorithms tested. First, the wide range of the MileAge delta values quantifies the latent patterns in the metabolomics data not captured by chronological age and suggests that our ageing clocks approximate (past) rate of biological ageing for people of the same chronological age. Second, the associations between most ageing clocks, health and mortality demonstrate that these clocks capture biologically relevant information, which may find applications in health tracking, nutrition or in clinical trials (e.g., sample stratification). A key finding across most ageing clock models tested here was that associations with health and mortality were stronger in individuals with an older predicted than chronological age and less so in individuals with a younger predicted than chronological age.

Several of the algorithms included in our comparison, including those that were most predictive of chronological age, mortality and health, allowed for non-linear relationships in the data, which were often not considered in previous studies (Panyard et al., 2022). A recent review found that most molecular ageing clocks were developed using linear models with regularization, whereas few used non-linear models (Xia, Wang, Yu, Chen, & Han, 2021).

Although it has been asserted that age-related physiological markers are linearly associated with age (Pyrkov et al., 2018), we have shown here and in previous studies (Mutz, Hoppen, et al., 2022; Mutz & Lewis, 2021; Mutz, Young, et al., 2022) that this does not apply to many biological markers. To enable comparability between studies, we provide a comprehensive set of statistical estimates and developed our ageing clock using metabolites measured in absolute concentrations instead of relative to other measures. Although it may be argued that quantification of a smaller number of metabolites would in principle be more feasible and convenient in clinical practice, all metabolites included in our model can be quantified from a single assay with minimal sample preparation required (Würtz et al., 2017).

## Limitations and future directions

Our study had certain limitations. Some algorithms that have previously been used to develop biological ageing clocks, for example deep learning (Zhavoronkov, Li, Ma, & Mamoshina, 2019), or approaches combining HSIC LASSO feature selection with non-linear support vector regression (Takahashi et al., 2020) were not tested, and could be explored in future studies. Our ageing clock provides a “systems level” indicator of age-related changes in metabolites; future clocks may be developed at the tissue or cellular level. Plasma samples may differ from other body fluids, e.g., serum, urine or cerebrospinal fluid. The metabolite coverage of the Nightingale Health platform is lipid focussed and mostly covers larger molecules, while there are potentially over 217,000 endogenous and exogenous molecules (Wishart et al., 2021). Nevertheless, this platform enables robust assessment of these metabolites in a single experiment (Würtz et al., 2017). Although more complex ageing clocks could be developed using technologies with wider metabolite coverage, prior analyses suggested that a majority of metabolites associated with age were related to lipid and amino acid pathways, both of which were included here (Menni et al., 2013). As in previous studies, we observed a systematic overestimation of age in young individuals and underestimation of age in older individuals (Nakamura et al., 1988). This bias is neither data nor method specific and may be explained by regression to the mean (Liang, Zhang, & Niu, 2019) of the training data (Jones, Lee, & Topol, 2022). To account for this bias, we have adjusted the predicted age and included chronological age as a covariate in the health association analyses (Xia et al., 2021). Longitudinal ageing metrics may be more robustly associated with certain health outcomes than cross-sectional ageing metrics, e.g., with physical and cognitive decline but not, for example, multimorbidity (Kuo et al., 2022). Future research should develop metabolomic ageing clocks from longitudinal data. Finally, the lack of independent data for external validation is a limitation.

## Conclusions

Metabolomic ageing clocks derived from multiple machine learning algorithms were robustly associated with health indicators and mortality. We found that our ageing clock (MileAge) derived from a Cubist rule-based regression model was overall most strongly associated with health indicators. Individuals with MileAge values greater than their chronological age, indicating accelerated biological ageing, were frailer, had shorter telomeres, were more likely to have a chronic illness, rated their health worse and had a higher mortality risk.

Metabolomic ageing clocks hold significant promise for research on lifespan and healthspan extension, as they provide a proxy of biological ageing that is potentially modifiable. These clocks may also help identify health risks before clinical symptoms emerge. As such, biological ageing clocks may contribute to health risk assessments, complementing clinical biomarkers. However, the utility of ageing clocks is not limited to risk stratification, but also in providing an intuitive, year-based metric for health tracking that may help individuals proactively engage with their health.

## Data Availability

The data used are available to all bona fide researchers for health-related research that is in the public interest, subject to an application process and approval criteria. Study materials are publicly available online at http://www.ukbiobank.ac.uk.

## Acknowledgments

This research is funded by the National Institute for Health and Care Research (NIHR) Maudsley Biomedical Research Centre at South London and Maudsley NHS Foundation Trust and King’s College London. The views expressed are those of the authors and not necessarily those of the NHS, the NIHR or the Department of Health and Social Care.

Computational analyses were supported by: King’s College London. (2023). King’s Computational Research, Engineering and Technology Environment (CREATE). Retrieved May 24, 2023, from https://doi.org/10.18742/rnvf-m076. This research has been conducted using data from UK Biobank, a major biomedical database. Data access permission has been granted under UK Biobank application 45514.

## Financial disclosures

CML is a member of the scientific advisory board of Myriad Neuroscience, has received speaker fees from SYNLAB and received consultancy fees from UCB. JM and RI declare no financial conflict of interest.

## Authorship contributions

JM conceived the idea of the study, acquired the data, carried out the analysis, interpreted the findings and wrote the manuscript. CML and RI interpreted the findings and reviewed the manuscript. All authors read and approved the final manuscript.

## Ethics

Ethical approval for the UK Biobank study has been granted by the National Information Governance Board for Health and Social Care and the NHS North West Multicentre Research Ethics Committee (11/NW/0382). No project-specific ethical approval is needed.

## Data sharing statement

The data used are available to all *bona fide* researchers for health-related research that is in the public interest, subject to an application process and approval criteria. Study materials are publicly available online at http://www.ukbiobank.ac.uk.

## Supplementary material

Supplementary information is available online.

## Notes

### Author Declarations

The data used are available to all bona fide researchers for health-related research that is in the public interest, subject to an application process and approval criteria. Study materials are publicly available online at http://www.ukbiobank.ac.uk. Ethical approval for the UK Biobank study has been granted by the National Information Governance Board for Health and Social Care and the NHS North West Multicentre Research Ethics Committee (11/NW/0382). No project-specific ethical approval is needed.

## References

1. Akker, E. B. v. d., Trompet, S., Wolf, J. J. H. B., Beekman, M., Suchiman, H. E. D., Deelen, J., . . . Slagboom, P. E. (2020). Metabolic Age Based on the BBMRI-NL 1H-NMR Metabolomics Repository as Biomarker of Age-related Disease. Circulation: Genomic and Precision Medicine, 13(5), 541–547. doi:doi:10.1161/CIRCGEN.119.002610

2. Bates, S., Hastie, T., & Tibshirani, R. Cross-Validation: What Does It Estimate and How Well Does It Do It? Journal of the American Statistical Association, 1–12. doi:10.1080/01621459.2023.2197686

3. Bizzarri, D., Reinders, M. J. T., Beekman, M., BBMRI-NL, Slagboom, P. E., & Akker, E. B. v. d. (2023). Technical report: A comprehensive comparison between different quantification versions of Nightingale Health’s 1H-NMR metabolomics platform. medRxiv, 2023.2007.2003.23292168. doi:10.1101/2023.07.03.23292168

4. Bortz, J., Guariglia, A., Klaric, L., Tang, D., Ward, P., Geer, M., . . . Joshi, P. K. (2023). Biological Age Estimation Using Circulating Blood Biomarkers. medRxiv, 2023.2002.2023.23285864. doi:10.1101/2023.02.23.23285864

5. Boser, B. E., Guyon, I. M., & Vapnik, V. N. (1992). *A training algorithm for optimal margin classifiers*. Paper presented at the Proceedings of the fifth annual workshop on Computational learning theory, Pittsburgh, Pennsylvania, USA. 10.1145/130385.130401

6. Breiman, L. (1996). Bagging predictors. Machine Learning, 24(2), 123–140. doi:10.1007/BF00058655

7. Breiman, L. (2001). Random Forests. Machine Learning, 45(1), 5–32. doi:10.1023/A:1010933404324

8. Breiman, L., Friedman, J., Olshen, R., & Stone, C. (1984). Classification and Regression Trees.

9. Buergel, T., Steinfeldt, J., Ruyoga, G., Pietzner, M., Bizzarri, D., Vojinovic, D., . . . Landmesser, U. (2022). Metabolomic profiles predict individual multidisease outcomes. Nature Medicine, 28(11), 2309–2320. doi:10.1038/s41591-022-01980-3

10. Bycroft, C., Freeman, C., Petkova, D., Band, G., Elliott, L. T., Sharp, K., . . . O’Connell, J. (2018). The UK Biobank resource with deep phenotyping and genomic data. Nature, 562(7726), 203–209. 10.1038/s41586-018-0579-z

11. Chen, B. H., Marioni, R. E., Colicino, E., Peters, M. J., Ward-Caviness, C. K., Tsai, P. C., . . . Horvath, S. (2016). DNA methylation-based measures of biological age: meta- analysis predicting time to death. Aging (Albany NY), 8(9), 1844–1865. doi:10.18632/aging.101020

12. Chen, T., & Guestrin, C. (2016). XGBoost: A Scalable Tree Boosting System. Paper presented at the Proceedings of the 22nd ACM SIGKDD International Conference on Knowledge Discovery and Data Mining, San Francisco, California, USA. 10.1145/2939672.2939785

13. Chipman, H. A., George, E. I., & McCulloch, R. E. (2010). BART: Bayesian additive regression trees. The Annals of Applied Statistics, 4(1), 266-298, 233. Retrieved from 10.1214/09-AOAS285

14. Cole, J. H., & Franke, K. (2017). Predicting Age Using Neuroimaging: Innovative Brain Ageing Biomarkers. Trends in Neurosciences, 40(12), 681–690. 10.1016/j.tins.2017.10.001

15. Cover, T., & Hart, P. (1967). Nearest neighbor pattern classification. IEEE Transactions on Information Theory, 13(1), 21–27. doi:10.1109/TIT.1967.1053964

16. Cox, D. R. (1972). Regression models and life=:Jtables. Journal of the Royal Statistical Society: Series B (Methodological), 34(2), 187–202.

17. de Lange, A. G., & Cole, J. H. (2020). Commentary: Correction procedures in brain-age prediction. Neuroimage Clin, 26, 102229. doi:10.1016/j.nicl.2020.102229

18. Deelen, J., Kettunen, J., Fischer, K., van der Spek, A., Trompet, S., Kastenmüller, G., . . . Slagboom, P. E. (2019). A metabolic profile of all-cause mortality risk identified in an observational study of 44,168 individuals. Nature Communications, 10(1), 3346. doi:10.1038/s41467-019-11311-9

19. Drucker, H., Burges, C. J., Kaufman, L., Smola, A., & Vapnik, V. (1996). Support Vector Regression Machines. Advances in neural information processing systems, 9.

20. Franceschi, C., Garagnani, P., Parini, P., Giuliani, C., & Santoro, A. (2018). Inflammaging: a new immune–metabolic viewpoint for age-related diseases. Nature Reviews Endocrinology, 14(10), 576–590. doi:10.1038/s41574-018-0059-4

21. Fransquet, P. D., Wrigglesworth, J., Woods, R. L., Ernst, M. E., & Ryan, J. (2019). The epigenetic clock as a predictor of disease and mortality risk: a systematic review and meta-analysis. Clinical Epigenetics, 11(1), 62. doi:10.1186/s13148-019-0656-7

22. Friedman, J., H. (1991). Multivariate Adaptive Regression Splines. The Annals of Statistics, 19(1), 1–67. doi:10.1214/aos/1176347963

23. Friedman, J. H., & Popescu, B. E. (2008). Predictive learning via rule ensembles. The Annals of Applied Statistics, 916-954.

24. Galkin, F., Kochetov, K., Koldasbayeva, D., Faria, M., Fung, H. H., Chen, A. X., & Zhavoronkov, A. (2022). Psychological factors substantially contribute to biological aging: evidence from the aging rate in Chinese older adults. Aging (Albany NY), 14(18), 7206–7222. doi:10.18632/aging.204264

25. Hannum, G., Guinney, J., Zhao, L., Zhang, L., Hughes, G., Sadda, S., . . . Zhang, K. (2013). Genome-wide Methylation Profiles Reveal Quantitative Views of Human Aging Rates. Molecular Cell, 49(2), 359–367. 10.1016/j.molcel.2012.10.016

26. Hertel, J., Frenzel, S., König, J., Wittfeld, K., Fuellen, G., Holtfreter, B., . . . Grabe, H. J. (2019). The informative error: A framework for the construction of individualized phenotypes. Statistical Methods in Medical Research, 28(5), 1427–1438. doi:10.1177/0962280218759138

27. Hertel, J., Friedrich, N., Wittfeld, K., Pietzner, M., Budde, K., Van der Auwera, S., . . . Grabe, H. J. (2016). Measuring Biological Age via Metabonomics: The Metabolic Age Score. Journal of Proteome Research, 15(2), 400–410. doi:10.1021/acs.jproteome.5b00561

28. Hoerl, A. E., & Kennard, R. W. (1970). Ridge Regression: Biased Estimation for Nonorthogonal Problems. Technometrics, 12(1), 55–67. doi:10.1080/00401706.1970.10488634

29. Hoogendijk, E. O., Afilalo, J., Ensrud, K. E., Kowal, P., Onder, G., & Fried, L. P. (2019). Frailty: implications for clinical practice and public health. The Lancet, 394(10206), 1365–1375. 10.1016/S0140-6736(19)31786-6

30. Horvath, S., & Raj, K. (2018). DNA methylation-based biomarkers and the epigenetic clock theory of ageing. Nature Reviews Genetics, 19, 371–384. 10.1038/s41576-018-0004-3

31. Jones, D. T., Lee, J., & Topol, E. J. (2022). Digitising brain age. The Lancet, 400(10357), 988. doi:10.1016/S0140-6736(22)01782-2

32. Kaplan, E. L., & Meier, P. (1958). Nonparametric estimation from incomplete observations. Journal of the American Statistical Association, 53(282), 457–481.

33. Kuhn, M., & Johnson, K. (2013a). Nonlinear Regression Models. In M. Kuhn & K. Johnson (Eds.), Applied Predictive Modeling (pp. 141–171). New York, NY: Springer New York.

34. Kuhn, M., & Johnson, K. (2013b). Regression Trees and Rule-Based Models. In M. Kuhn & K. Johnson (Eds.), Applied Predictive Modeling (pp. 173–220). New York, NY: Springer New York.

35. Kuo, P.-L., Schrack, J. A., Levine, M. E., Shardell, M. D., Simonsick, E. M., Chia, C. W., . . . Ferrucci, L. (2022). Longitudinal phenotypic aging metrics in the Baltimore Longitudinal Study of Aging. Nature Aging, 2(7), 635–643. doi:10.1038/s43587-022-00243-7

36. Lai, T.-P., Wright, W. E., & Shay, J. W. (2018). Comparison of telomere length measurement methods. Philosophical Transactions of the Royal Society B: Biological Sciences, 373(1741), 20160451.

37. Lawton, K. A., Berger, A., Mitchell, M., Milgram, K. E., Evans, A. M., Guo, L., . . . Milburn, M. V. (2008). Analysis of the adult human plasma metabolome. Pharmacogenomics, 9(4), 383–397. doi:10.2217/14622416.9.4.383

38. Levine, M. E., Lu, A. T., Quach, A., Chen, B. H., Assimes, T. L., Bandinelli, S., . . . Horvath, S. (2018). An epigenetic biomarker of aging for lifespan and healthspan. Aging (Albany NY), 10(4), 573–591. doi:10.18632/aging.101414

39. Liang, H., Zhang, F., & Niu, X. (2019). Investigating systematic bias in brain age estimation with application to post-traumatic stress disorders. Human Brain Mapping, 40(11), 3143–3152. 10.1002/hbm.24588

40. López-Otín, C., Blasco, M. A., Partridge, L., Serrano, M., & Kroemer, G. (2023). Hallmarks of aging: An expanding universe. Cell, 186(2), 243–278. doi:10.1016/j.cell.2022.11.001

41. Lu, A. T., Quach, A., Wilson, J. G., Reiner, A. P., Aviv, A., Raj, K., . . . Horvath, S. (2019). DNA methylation GrimAge strongly predicts lifespan and healthspan. Aging (Albany NY), 11(2), 303–327. doi:10.18632/aging.101684

42. Macdonald-Dunlop, E., Taba, N., Klarić, L., Frkatović, A., Walker, R., Hayward, C., . . . Joshi, P. K. (2022). A catalogue of omics biological ageing clocks reveals substantial commonality and associations with disease risk. Aging (Albany NY), 14(2), 623–659. doi:10.18632/aging.203847

43. Menni, C., Kastenmüller, G., Petersen, A. K., Bell, J. T., Psatha, M., Tsai, P.-C., . . . Valdes, A. M. (2013). Metabolomic markers reveal novel pathways of ageing and early development in human populations. International Journal of Epidemiology, 42(4), 1111–1119. doi:10.1093/ije/dyt094

44. Moqri, M., Herzog, C., Poganik, J. R., Justice, J., Belsky, D. W., Higgins-Chen, A., . . . Gladyshev, V. N. (2023). Biomarkers of aging for the identification and evaluation of longevity interventions. Cell, 186(18), 3758–3775. doi:10.1016/j.cell.2023.08.003

45. Mutz, J., Choudhury, U., Zhao, J., & Dregan, A. (2022). Frailty in individuals with depression, bipolar disorder and anxiety disorders: longitudinal analyses of all-cause mortality. medRxiv, 2022.2002.2023.22271065. doi:10.1101/2022.02.23.22271065

46. Mutz, J., Hoppen, T. H., Fabbri, C., & Lewis, C. M. (2022). Anxiety disorders and age- related changes in physiology. The British Journal of Psychiatry, 1-10. doi:10.1192/bjp.2021.189

47. Mutz, J., & Lewis, C. M. (2021). Lifetime depression and age-related changes in body composition, cardiovascular function, grip strength and lung function: sex-specific analyses in the UK Biobank. Aging, 13(13), 17038–17079.

48. Mutz, J., & Lewis, C. M. (2022). Cross-classification between self-rated health and health status: longitudinal analyses of all-cause mortality and leading causes of death in the UK. Scientific Reports, 12(1), 459. 10.1038/s41598-021-04016-x

49. Mutz, J., Roscoe, C. J., & Lewis, C. M. (2021). Exploring health in the UK Biobank: associations with sociodemographic characteristics, psychosocial factors, lifestyle and environmental exposures. BMC Medicine, 19(1), 240. doi:10.1186/s12916-021-02097-z

50. Mutz, J., Young, A. H., & Lewis, C. M. (2022). Age-related changes in physiology in individuals with bipolar disorder. Journal of Affective Disorders, 296, 157–168. 10.1016/j.jad.2021.09.027

51. Nakamura, E., Miyao, K., & Ozeki, T. (1988). Assessment of biological age by principal component analysis. Mechanisms of Ageing and Development, 46(1), 1–18. 10.1016/0047-6374(88)90109-1

52. Panyard, D. J., Yu, B., & Snyder, M. P. (2022). The metabolomics of human aging: Advances, challenges, and opportunities. Science Advances, 8(42), eadd6155. doi:doi:10.1126/sciadv.add6155

53. Pyrkov, T. V., Slipensky, K., Barg, M., Kondrashin, A., Zhurov, B., Zenin, A., . . . Fedichev, P. O. (2018). Extracting biological age from biomedical data via deep learning: too much of a good thing? Scientific Reports, 8(1), 5210. doi:10.1038/s41598-018-23534-9

54. Quinlan, J. R. (1992). *Learning with continuous classes.* Paper presented at the 5th Australian joint conference on artificial intelligence.

55. Robinson, O., Chadeau Hyam, M., Karaman, I., Climaco Pinto, R., Ala-Korpela, M., Handakas, E., . . . Vineis, P. (2020). Determinants of accelerated metabolomic and epigenetic aging in a UK cohort. Aging Cell, 19(6), e13149. 10.1111/acel.13149

56. Robinson, O., & Lau, C. E. (2020). Measuring biological age using metabolomics. Aging (Albany NY), 12(22), 22352–22353. doi:10.18632/aging.104216

57. Rutledge, J., Oh, H., & Wyss-Coray, T. (2022). Measuring biological age using omics data. Nature Reviews Genetics. doi:10.1038/s41576-022-00511-7

58. Soininen, P., Kangas, A. J., Würtz, P., Suna, T., & Ala-Korpela, M. (2015). Quantitative Serum Nuclear Magnetic Resonance Metabolomics in Cardiovascular Epidemiology and Genetics. Circulation: Cardiovascular Genetics, 8(1), 192–206. doi:doi:10.1161/CIRCGENETICS.114.000216

59. Solovev, I., Shaposhnikov, M., & Moskalev, A. (2020). Multi-omics approaches to human biological age estimation. Mechanisms of Ageing and Development, 185, 111192. 10.1016/j.mad.2019.111192

60. Takahashi, Y., Ueki, M., Yamada, M., Tamiya, G., Motoike, I. N., Saigusa, D., . . . Tomita, H. (2020). Improved metabolomic data-based prediction of depressive symptoms using nonlinear machine learning with feature selection. Translational Psychiatry, 10(1), 157. doi:10.1038/s41398-020-0831-9

61. Tibshirani, R. (1996). Regression Shrinkage and Selection Via the Lasso. Journal of the Royal Statistical Society: Series B (Methodological), 58(1), 267–288. 10.1111/j.2517-6161.1996.tb02080.x

62. Vidaki, A., Ballard, D., Aliferi, A., Miller, T. H., Barron, L. P., & Syndercombe Court, D. (2017). DNA methylation-based forensic age prediction using artificial neural networks and next generation sequencing. Forensic Science International: Genetics, 28, 225–236. doi:10.1016/j.fsigen.2017.02.009

63. Wishart, D. S., Guo, A., Oler, E., Wang, F., Anjum, A., Peters, H., . . . Gautam, V. (2021). HMDB 5.0: the Human Metabolome Database for 2022. Nucleic Acids Research, 50(D1), D622–D631. doi:10.1093/nar/gkab1062

64. Wold, S., Sjöström, M., & Eriksson, L. (2001). PLS-regression: a basic tool of chemometrics. Chemometrics and Intelligent Laboratory Systems, 58(2), 109–130. 10.1016/S0169-7439(01)00155-1

65. Würtz, P., Kangas, A. J., Soininen, P., Lawlor, D. A., Davey Smith, G., & Ala-Korpela, M. (2017). Quantitative Serum Nuclear Magnetic Resonance Metabolomics in Large-Scale Epidemiology: A Primer on -Omic Technologies. American Journal of Epidemiology, 186(9), 1084–1096. doi:10.1093/aje/kwx016

66. Xia, X., Wang, Y., Yu, Z., Chen, J., & Han, J.-D. J. (2021). Assessing the rate of aging to monitor aging itself. Ageing Research Reviews, 69, 101350. 10.1016/j.arr.2021.101350

67. Yu, Z., Zhai, G., Singmann, P., He, Y., Xu, T., Prehn, C., . . . Wang-Sattler, R. (2012). Human serum metabolic profiles are age dependent. Aging Cell, 11(6), 960–967. 10.1111/j.1474-9726.2012.00865.x

68. Zhang, Q., Vallerga, C. L., Walker, R. M., Lin, T., Henders, A. K., Montgomery, G. W., . . . Visscher, P. M. (2019). Improved precision of epigenetic clock estimates across tissues and its implication for biological ageing. Genome Medicine, 11(1), 54. doi:10.1186/s13073-019-0667-1

69. Zhavoronkov, A., Li, R., Ma, C., & Mamoshina, P. (2019). Deep biomarkers of aging and longevity: from research to applications. Aging (Albany NY), 11(22), 10771–10780. doi:10.18632/aging.102475

70. Zou, H., & Hastie, T. (2005). Regularization and variable selection via the elastic net. Journal of the Royal Statistical Society: Series B (Statistical Methodology), 67(2), 301–320. 10.1111/j.1467-9868.2005.00503.x

