## Supplementary material for "Metabolomic Age (MileAge) predicts health and lifespan: a comparison of multiple machine learning algorithms"

This material accompanies the article

Authors: Julian **M**utz, Raquel **I**niesta and Cathryn M **Le**wis

#### 1. Hyperparameter tuning

**Table S1**. Details hyperparameter tuning

| | | Pre-processing | | | Hyperparameters | | | |
|---|---|---|---|---|---|---|---|---|
| Algorithm | Engine | Normalize | Zero variance | None | Fixed | Tuned | Grid size | Racing |
| Ridge | glmnet | × | | | 1 | 1 | 10 | |
| LASSO | glmnet | × | | | 1 | 1 | 10 | |
| Elastic net | glmnet | × | | | 2 | | 100 | |
| PLSR | mixOmics | × | × | | 2 | | 40 | |
| KNN | kknn | × | | | 3 | | 125 | × |
| MARS | earth | | | × | 3 | | 48 | × |
| MARS ensemble | earth | | | × | 3 | 1 | 48 | |
| SVM linear | kernlab | × | | | 2 | | 100 | |
| SVM polynomial | kernlab | × | | | 4 | | 375 | × |
| SVM radial | kernlab | × | | | 3 | | 125 | × |
| Regression tree | rpart | | | × | 3 | | 1000 | |
| Bagging | rpart | | | × | 3 | 1 | 125 | × |
| Random forest | ranger | | | × | 3 | | 125 | × |
| XGBoost | xgboost | | | × | 5 | 3 | 3125 | × |
| BART | dbarts | | | × | 3 | 1 | 125 | × |
| Cubist rules | Cubist | | | × | 3 | | 125 | × |
| RuleFit ensemble | xrf | | | × | 5 | 3 | 3125 | × |

*Note:* See Panel 1 for model abbreviations.

#### 2. UK Biobank data fields

**Table S2**. UK Biobank data fields

| Data field | Variable name |
|---|---|
| *Sample characteristics* | |
| 31 | Sex |
| 53 | Date of attending assessment centre |
| 54 | UK Biobank assessment centre |
| 74 | Fasting time |
| 738 | Average total household income before tax |
| 2724 | Had menopause |
| 3140 | Pregnant |
| 6138[1] | Qualifications |
| 6141[1] | How are people in household related to participant |
| 21000[1] | Ethnic background |
| 21003 | Age when attended assessment centre |
| 22001 | Genetic sex |
| 20116 | Smoking status |
| 23104 | Body mass index (BMI) |
| *Health indicators* | |
| 134[1] | Number of self-reported cancers |
| 135[1] | Number of self-reported non-cancer illnesses |
| 2178 | Overall health rating |
| 2188 | Long-standing illness, disability of infirmity |
| 20001[1] | Cancer code, self-reported |
| 20002[1] | Non-cancer illness code, self-reported |
| 40000 | Date of death |
| 22192 | Z-adjusted T/S log |

*Note:* [1]These variables were further processed and categories used in the present study are shown in the main body of the text.

Details on the data fields used to derive the frailty index and frailty phenotype can be found in the below publication:

Mutz, J., Choudhury, U., Zhao, J., & Dregan, A. (2022). Frailty in individuals with depression, bipolar disorder and anxiety disorders: longitudinal analyses of all-cause mortality. *BMC Medicine, 20*(1), 274. doi:10.1186/s12916-022-02474-2

Details on the data fields for the Nightingale Health nuclear magnetic resonance (NMR) spectroscopy metabolomic biomarkers can be found in the UK Biobank showcase:

https://biobank.ndph.ox.ac.uk/showcase/label.cgi?id=220

#### 3. Sample flowchart

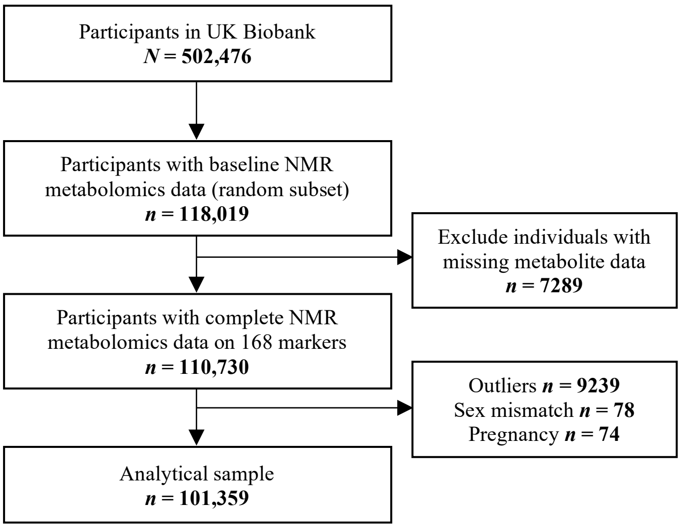

**Figure S1**. Study sample flowchart. Outliers were defined as metabolite values 4×IQR above or below the median. NMR = nuclear magnetic resonance; IQR = interquartile range.

#### 4. Distributions of metabolite levels by chronological age

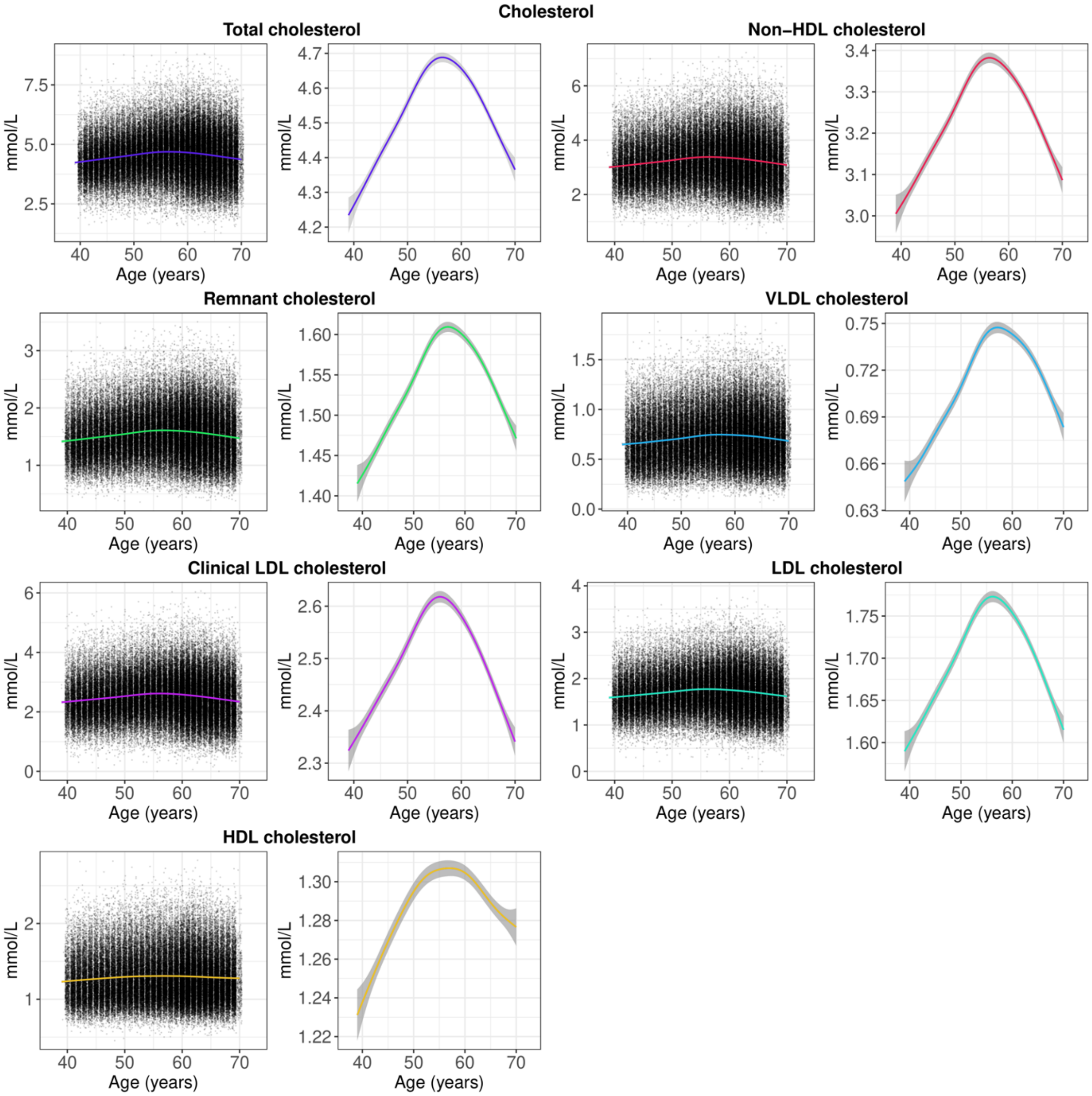

**Figure S2**. Distribution of cholesterol levels by chronological age, showing scatter plots of all observations and smooth curves (note the difference in y-axis scale). The smooth curves were estimated using generalised additive models, with shaded areas corresponding to 95% confidence intervals. HDL = high-density lipoprotein; VLDL = very low-density lipoprotein; LDL = low-density lipoprotein.

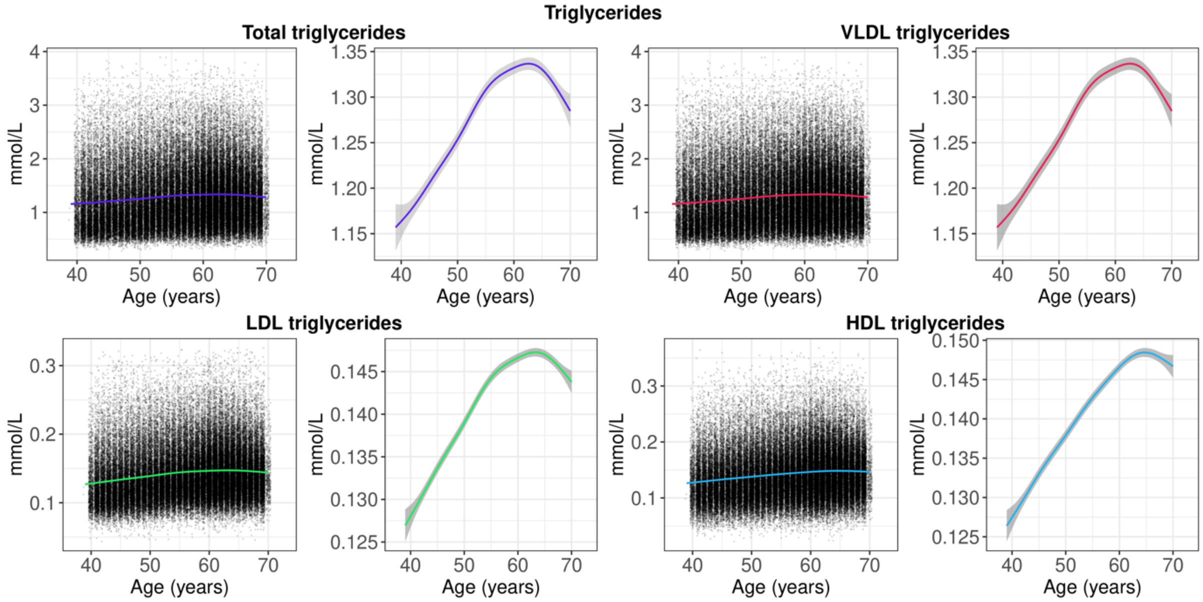

**Figure S3**. Distribution of triglyceride levels by chronological age, showing scatter plots of all observations and smooth curves (note the difference in y-axis scale). The smooth curves were estimated using generalised additive models, with shaded areas corresponding to 95% confidence intervals. VLDL = very low-density lipoprotein; LDL = low-density lipoprotein; HDL = high-density lipoprotein.

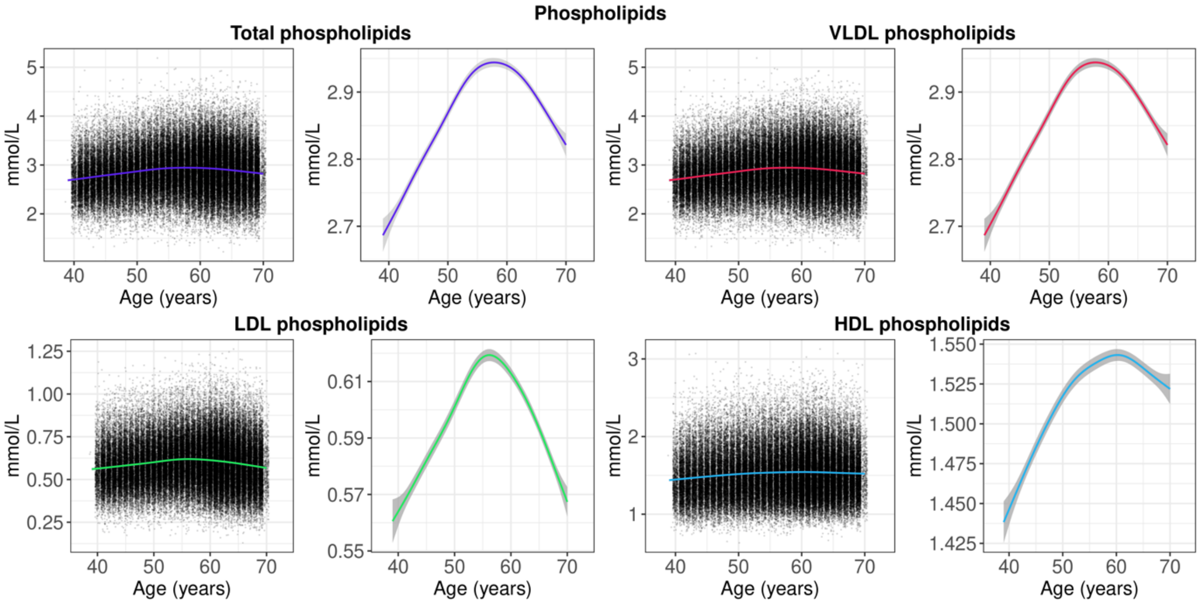

**Figure S4**. Distribution of phospholipid levels by chronological age, showing scatter plots of all observations and smooth curves (note the difference in y-axis scale). The smooth curves were estimated using generalised additive models, with shaded areas corresponding to 95% confidence intervals. VLDL = very low-density lipoprotein; LDL = low-density lipoprotein; HDL = high-density lipoprotein.

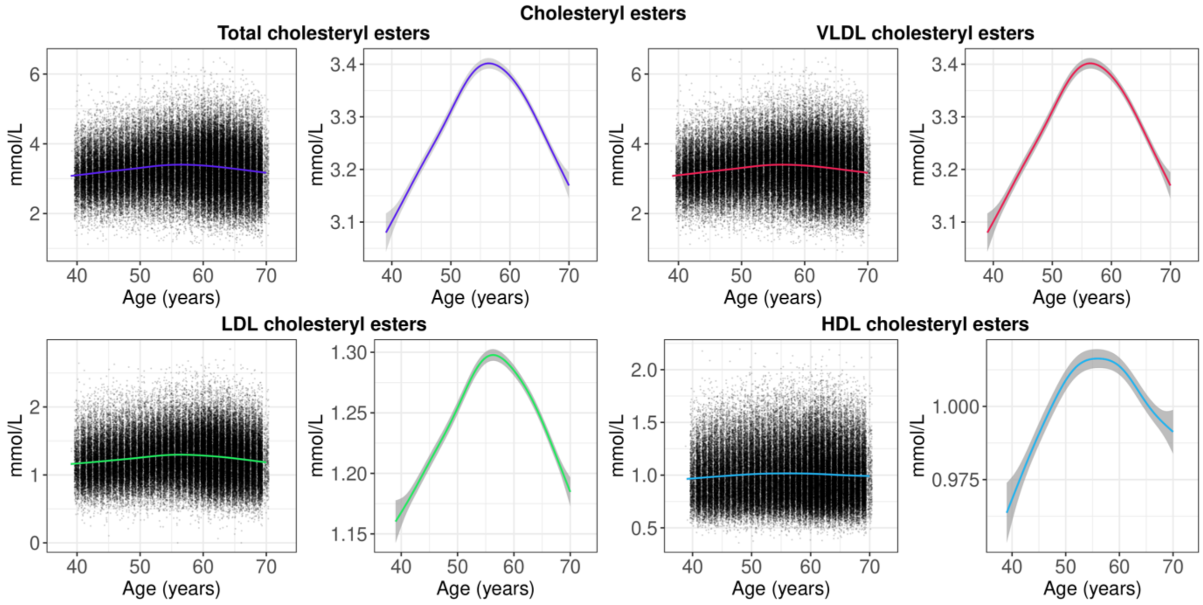

**Figure S5**. Distribution of cholesteryl ester levels by chronological age, showing scatter plots of all observations and smooth curves (note the difference in y-axis scale). The smooth curves were estimated using generalised additive models, with shaded areas corresponding to 95% confidence intervals. VLDL = very low-density lipoprotein; LDL = low-density lipoprotein; HDL = high-density lipoprotein.

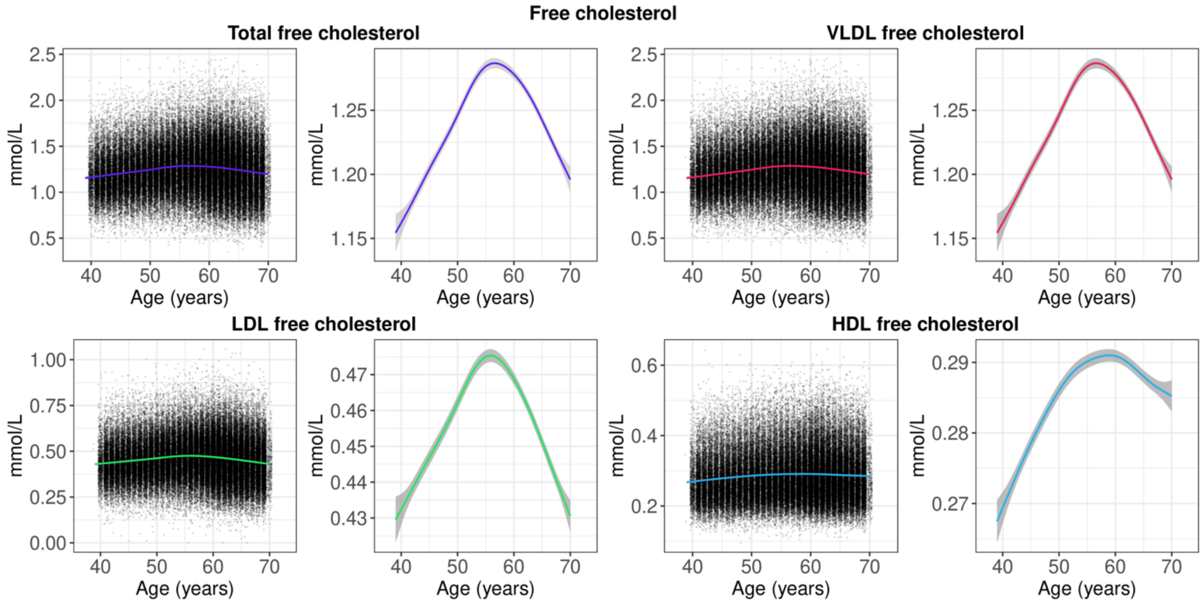

**Figure S6**. Distribution of free cholesterol levels by chronological age, showing scatter plots of all observations and smooth curves (note the difference in y-axis scale). The smooth curves were estimated using generalised additive models, with shaded areas corresponding to 95% confidence intervals. VLDL = very low-density lipoprotein; LDL = low-density lipoprotein; HDL = high-density lipoprotein.

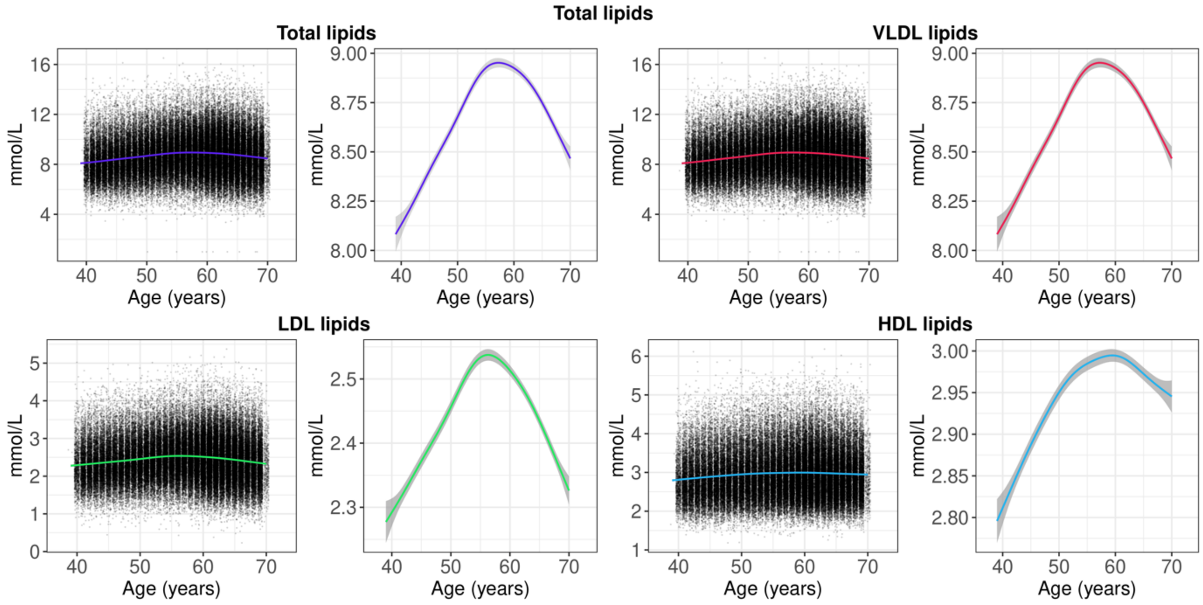

**Figure S7**. Distribution of total lipid levels by chronological age, showing scatter plots of all observations and smooth curves (note the difference in y-axis scale). The smooth curves were estimated using generalised additive models, with shaded areas corresponding to 95% confidence intervals. VLDL = very low-density lipoprotein; LDL = low-density lipoprotein; HDL = high-density lipoprotein.

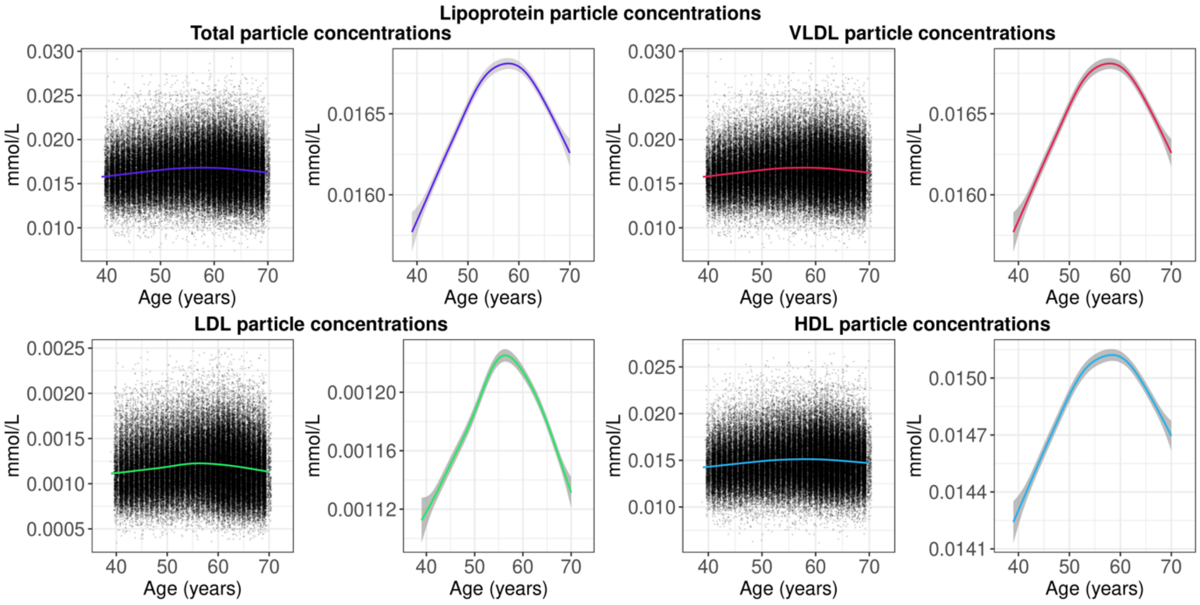

**Figure S8**. Distribution of lipoprotein particle concentrations by chronological age, showing scatter plots of all observations and smooth curves (note the difference in y-axis scale). The smooth curves were estimated using generalised additive models, with shaded areas corresponding to 95% confidence intervals. VLDL = very low-density lipoprotein; LDL = low-density lipoprotein; HDL = high-density lipoprotein.

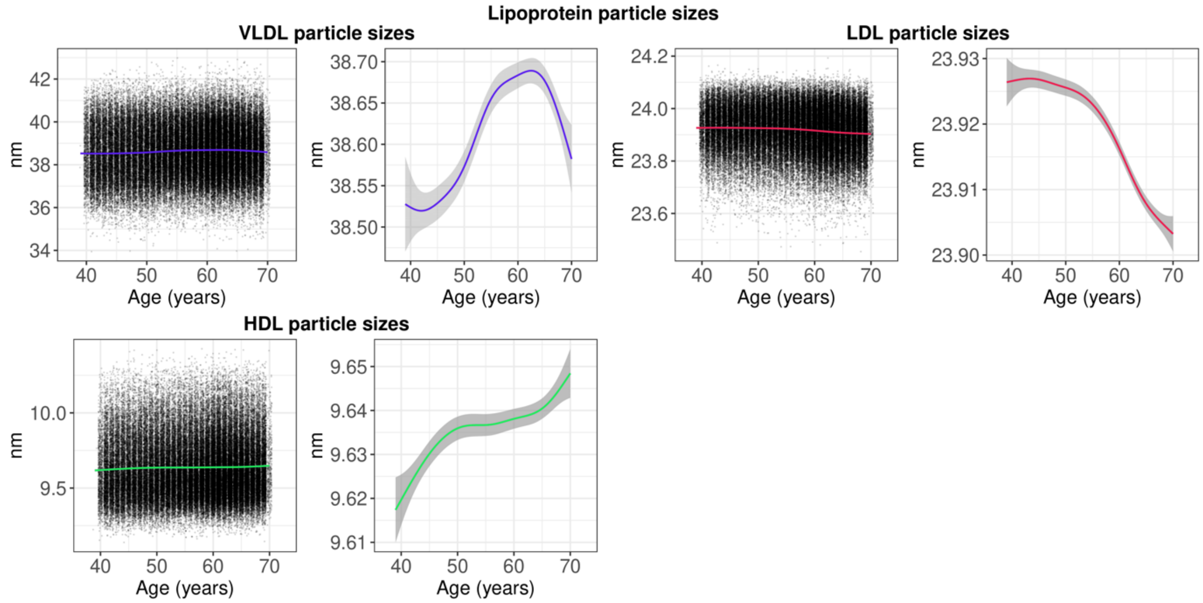

**Figure S9**. Distribution of lipoprotein particle sizes by chronological age, showing scatter plots of all observations and smooth curves (note the difference in y-axis scale). The smooth curves were estimated using generalised additive models, with shaded areas corresponding to 95% confidence intervals. VLDL = very low-density lipoprotein; LDL = low-density lipoprotein; HDL = high-density lipoprotein.

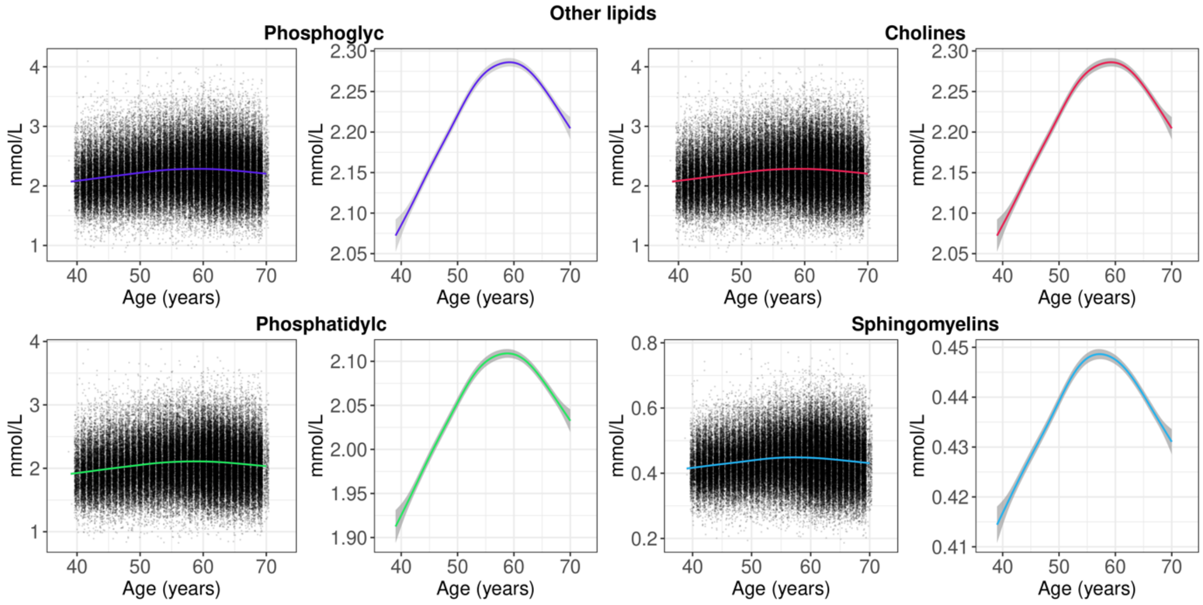

**Figure S10**. Distribution of other lipid levels by chronological age, showing scatter plots of all observations and smooth curves (note the difference in y-axis scale). The smooth curves were estimated using generalised additive models, with shaded areas corresponding to 95% confidence intervals.

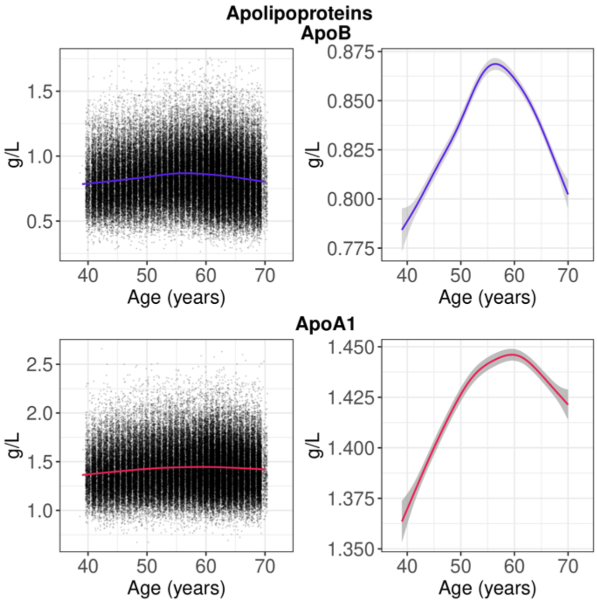

**Figure S11**. Distribution of apolipoprotein levels by chronological age, showing scatter plots of all observations and smooth curves (note the difference in y-axis scale). The smooth curves were estimated using generalised additive models, with shaded areas corresponding to 95% confidence intervals. ApoB = apolipoprotein B; ApoA1 = apolipoprotein A1.

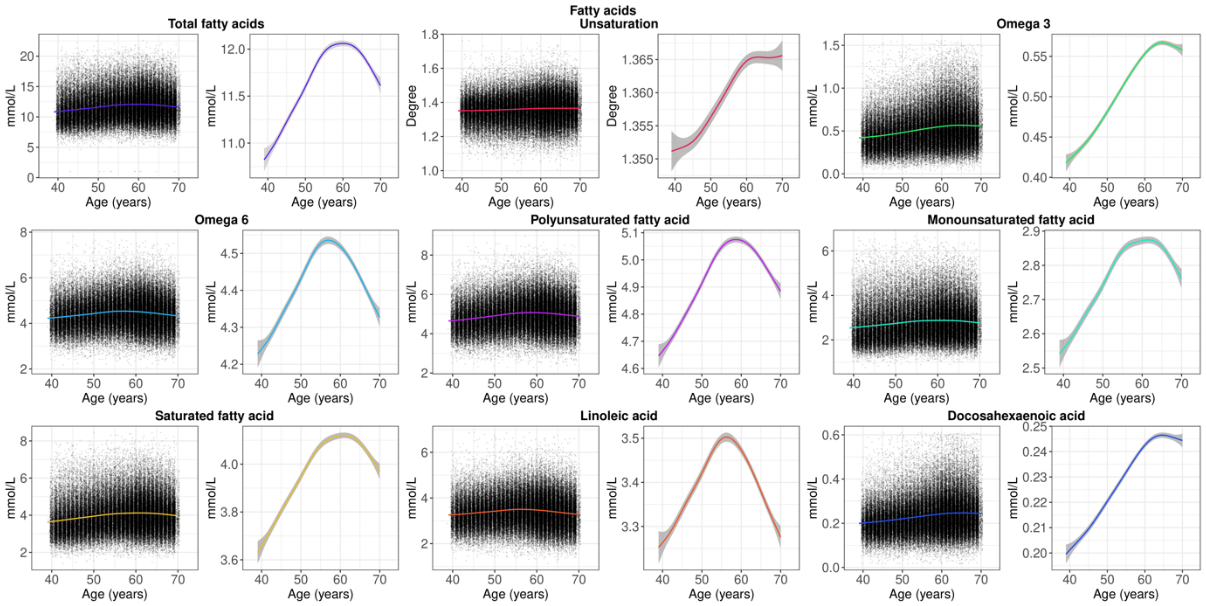

**Figure S12**. Distribution of fatty acid levels by chronological age, showing scatter plots of all observations and smooth curves (note the difference in y-axis scale). The smooth curves were estimated using generalised additive models, with shaded areas corresponding to 95% confidence intervals.

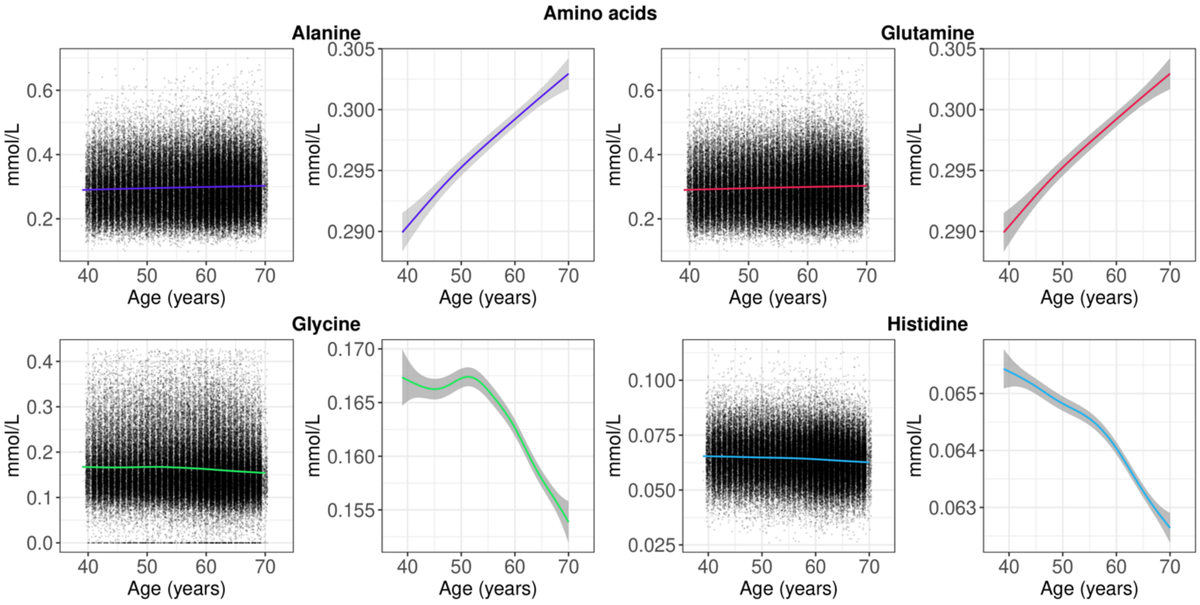

**Figure S13**. Distribution of amino acid levels by chronological age, showing scatter plots of all observations and smooth curves (note the difference in y-axis scale). The smooth curves were estimated using generalised additive models, with shaded areas corresponding to 95% confidence intervals.

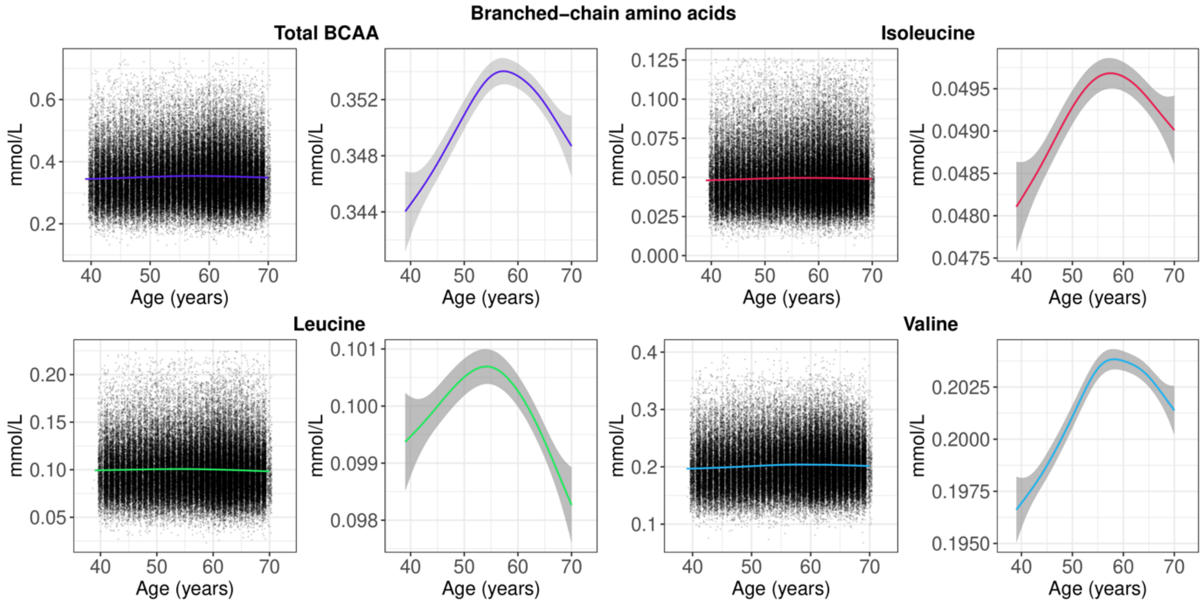

**Figure S14**. Distribution of branched-chain amino acid levels by chronological age, showing scatter plots of all observations and smooth curves (note the difference in y-axis scale). The smooth curves were estimated using generalised additive models, with shaded areas corresponding to 95% confidence intervals. BCAA = branched-chain amino acids.

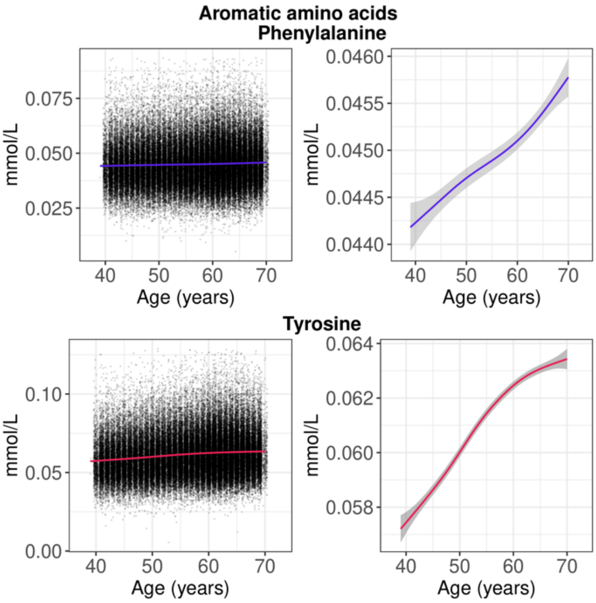

**Figure S15**. Distribution of aromatic amino acid levels by chronological age, showing scatter plots of all observations and smooth curves (note the difference in y-axis scale). The smooth curves were estimated using generalised additive models, with shaded areas corresponding to 95% confidence intervals.

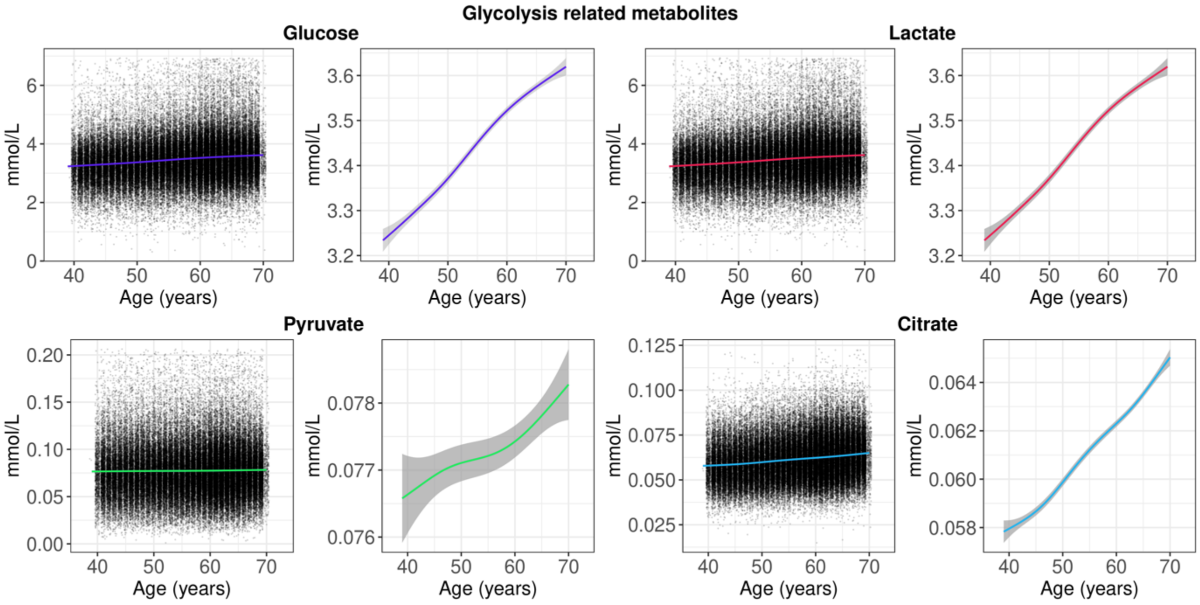

**Figure S16**. Distribution of glycolysis related metabolite levels by chronological age, showing scatter plots of all observations and smooth curves (note the difference in y-axis scale). The smooth curves were estimated using generalised additive models, with shaded areas corresponding to 95% confidence intervals.

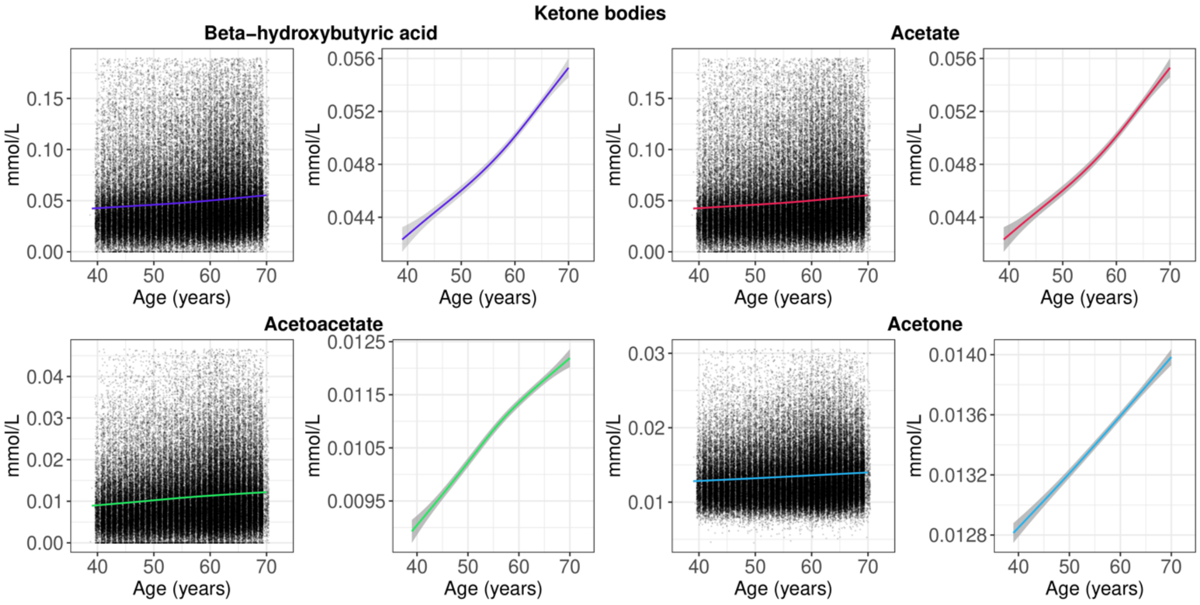

**Figure S17**. Distribution of ketone body levels by chronological age, showing scatter plots of all observations and smooth curves (note the difference in y-axis scale). The smooth curves were estimated using generalised additive models, with shaded areas corresponding to 95% confidence intervals.

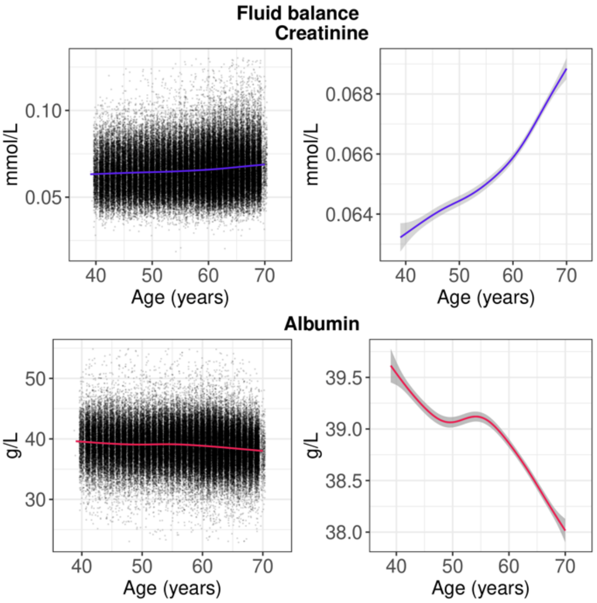

**Figure S18**. Distribution of metabolites involved in fluid balance by chronological age, showing scatter plots of all observations and smooth curves (note the difference in y-axis scale). The smooth curves were estimated using generalised additive models, with shaded areas corresponding to 95% confidence intervals.

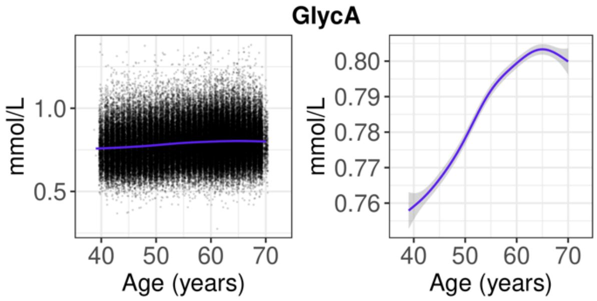

**Figure S19**. Distribution of glycoprotein acetyl levels by chronological age, showing scatter plots of all observations and smooth curves (note the difference in y-axis scale). The smooth curves were estimated using generalised additive models, with shaded areas corresponding to 95% confidence intervals. GlycA = glycoprotein acetyls.

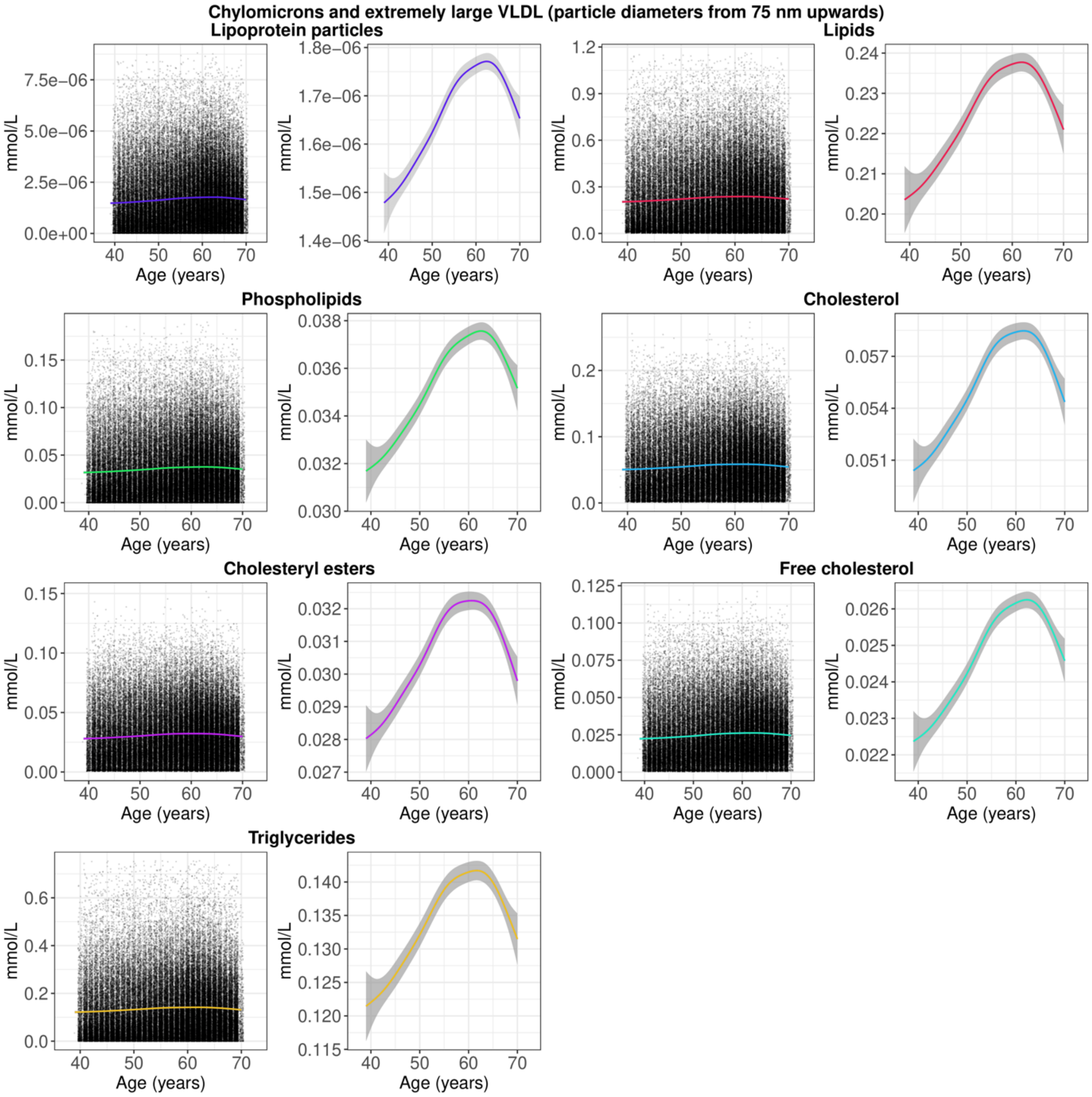

**Figure S20**. Distribution of chylomicrons and extremely large VLDL (particle diameters from 75 nm upwards) by chronological age, showing scatter plots of all observations and smooth curves (note the difference in y-axis scale). The smooth curves were estimated using generalised additive models, with shaded areas corresponding to 95% confidence intervals. VLDL = very low-density lipoprotein.

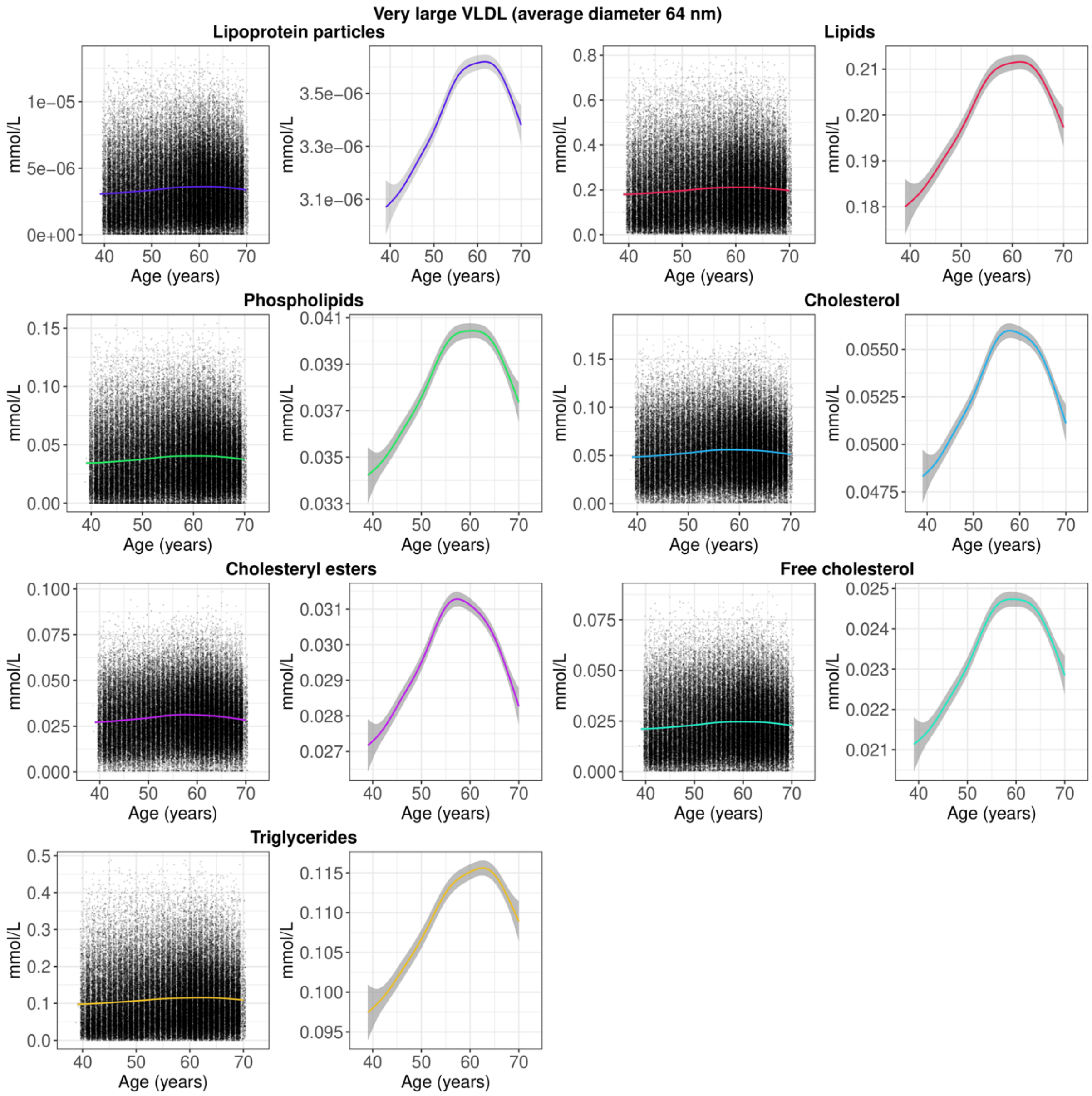

**Figure S21**. Distribution of very large VLDL (average diameter 64 nm) by chronological age, showing scatter plots of all observations and smooth curves (note the difference in y-axis scale). The smooth curves were estimated using generalised additive models, with shaded areas corresponding to 95% confidence intervals. VLDL = very low-density lipoprotein.

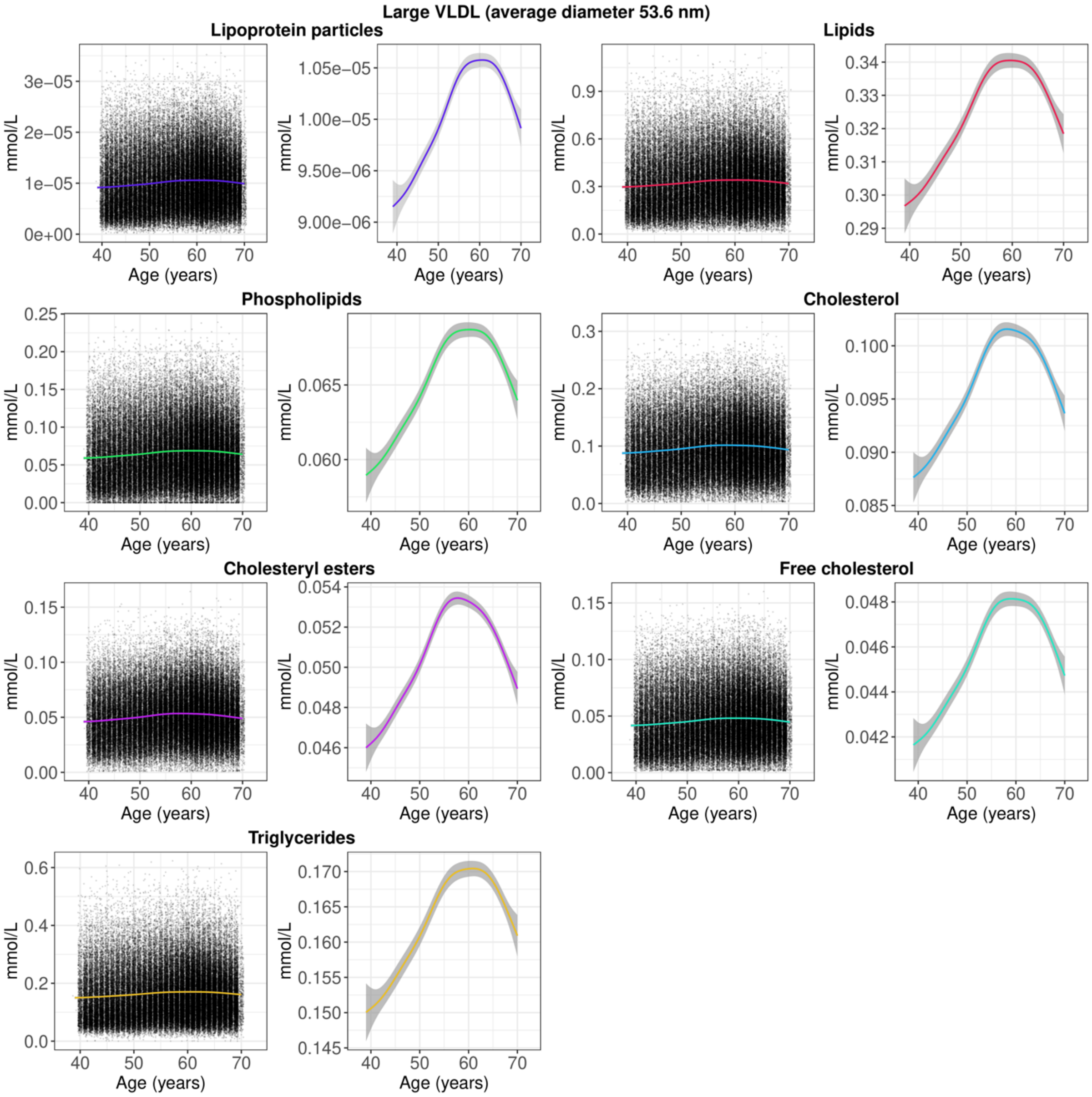

**Figure S22**. Distribution of large VLDL (average diameter 53.6 nm) by chronological age, showing scatter plots of all observations and smooth curves (note the difference in y-axis scale). The smooth curves were estimated using generalised additive models, with shaded areas corresponding to 95% confidence intervals. VLDL = very low-density lipoprotein.

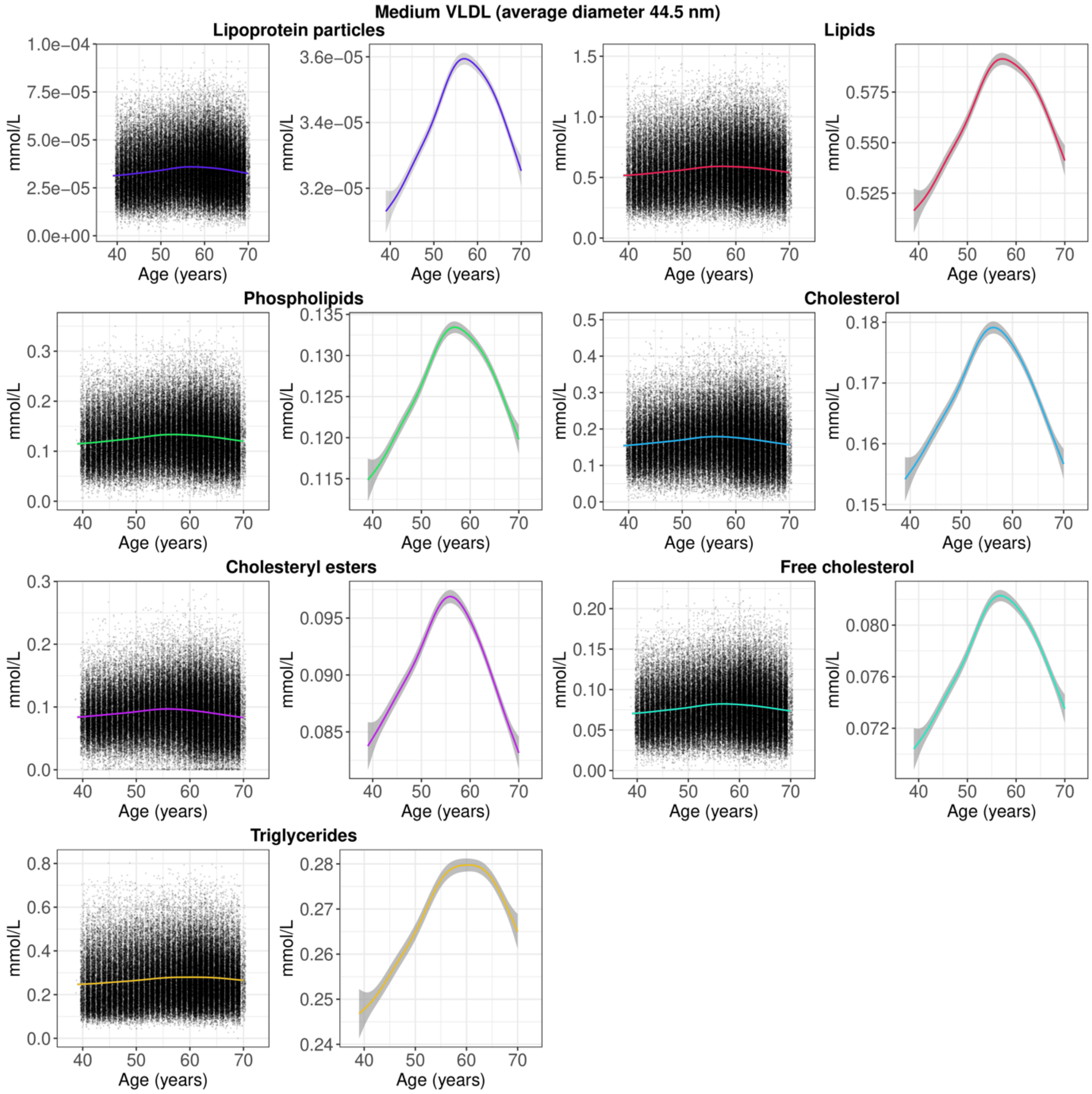

**Figure S23**. Distribution of medium VLDL (average diameter 44.5 nm) by chronological age, showing scatter plots of all observations and smooth curves (note the difference in y-axis scale). The smooth curves were estimated using generalised additive models, with shaded areas corresponding to 95% confidence intervals. VLDL = very low-density lipoprotein.

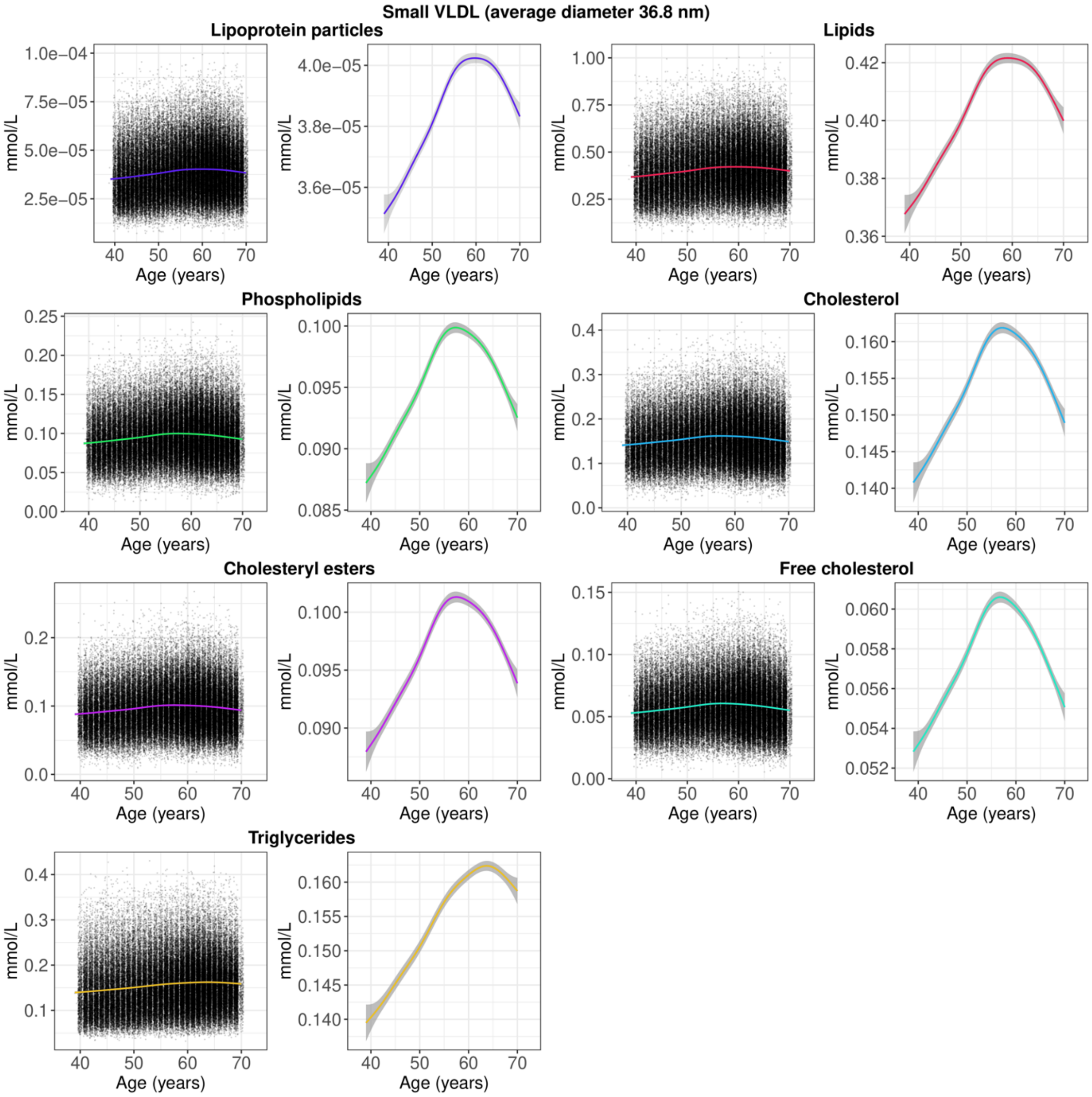

**Figure S24**. Distribution of small VLDL (average diameter 36.8 nm) by chronological age, showing scatter plots of all observations and smooth curves (note the difference in y-axis scale). The smooth curves were estimated using generalised additive models, with shaded areas corresponding to 95% confidence intervals. VLDL = very low-density lipoprotein.

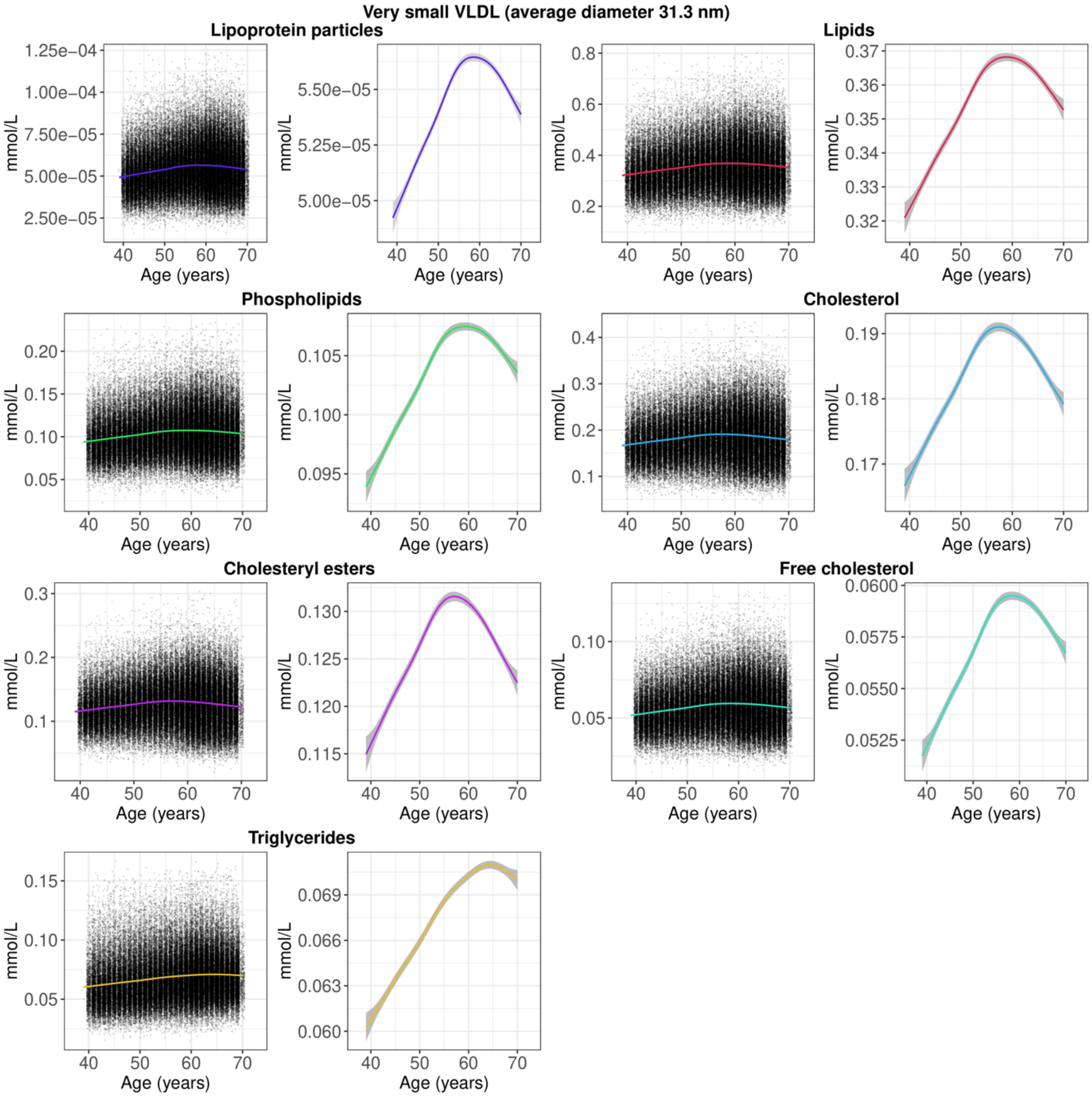

**Figure S25**. Distribution of very small VLDL (average diameter 31.3 nm) by chronological age, showing scatter plots of all observations and smooth curves (note the difference in y-axis scale). The smooth curves were estimated using generalised additive models, with shaded areas corresponding to 95% confidence intervals. VLDL = very low-density lipoprotein.

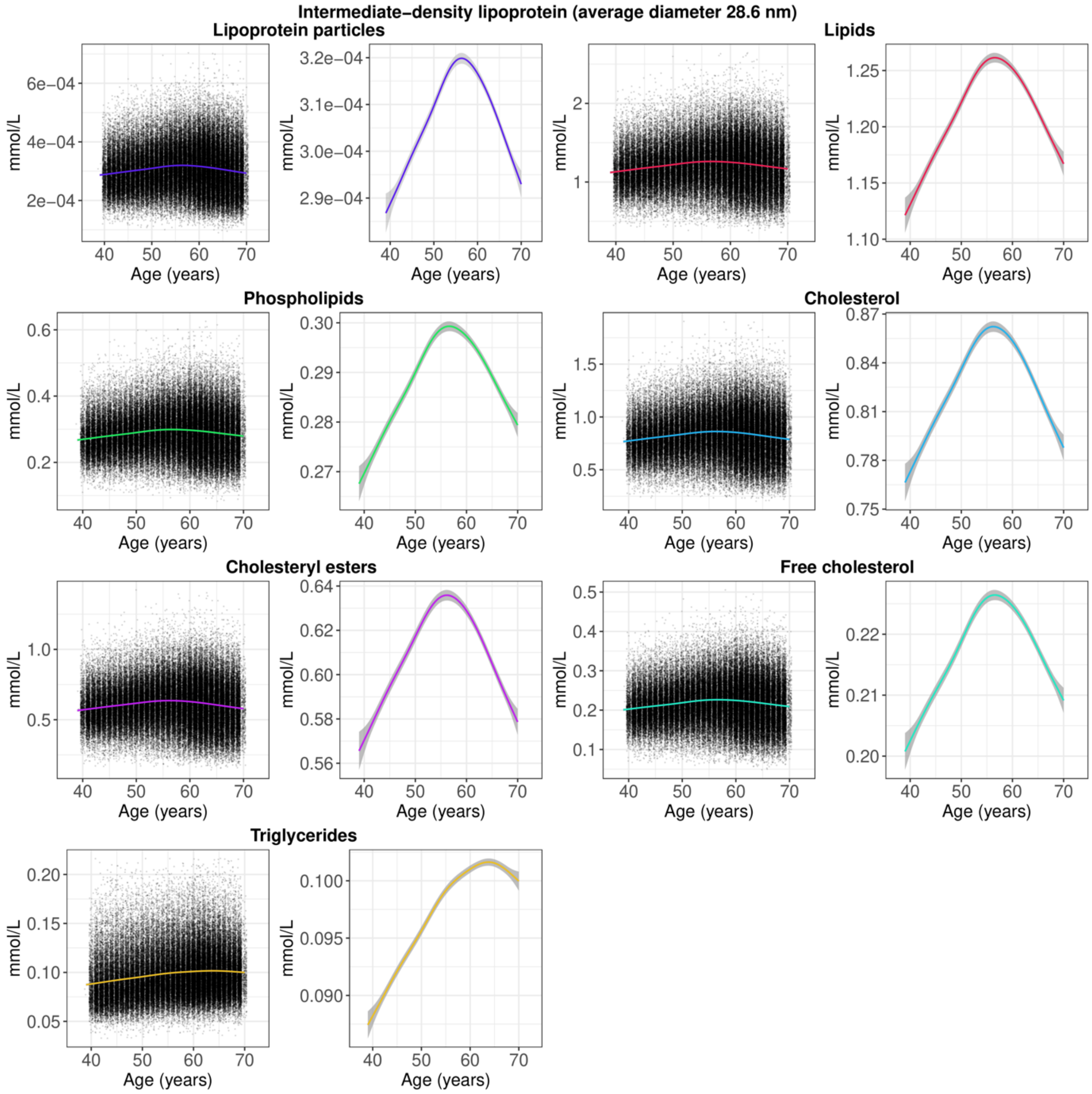

**Figure S26**. Distribution of intermediate-density lipoprotein (average diameter 28.6 nm) by chronological age, showing scatter plots of all observations and smooth curves (note the difference in y-axis scale). The smooth curves were estimated using generalised additive models, with shaded areas corresponding to 95% confidence intervals.

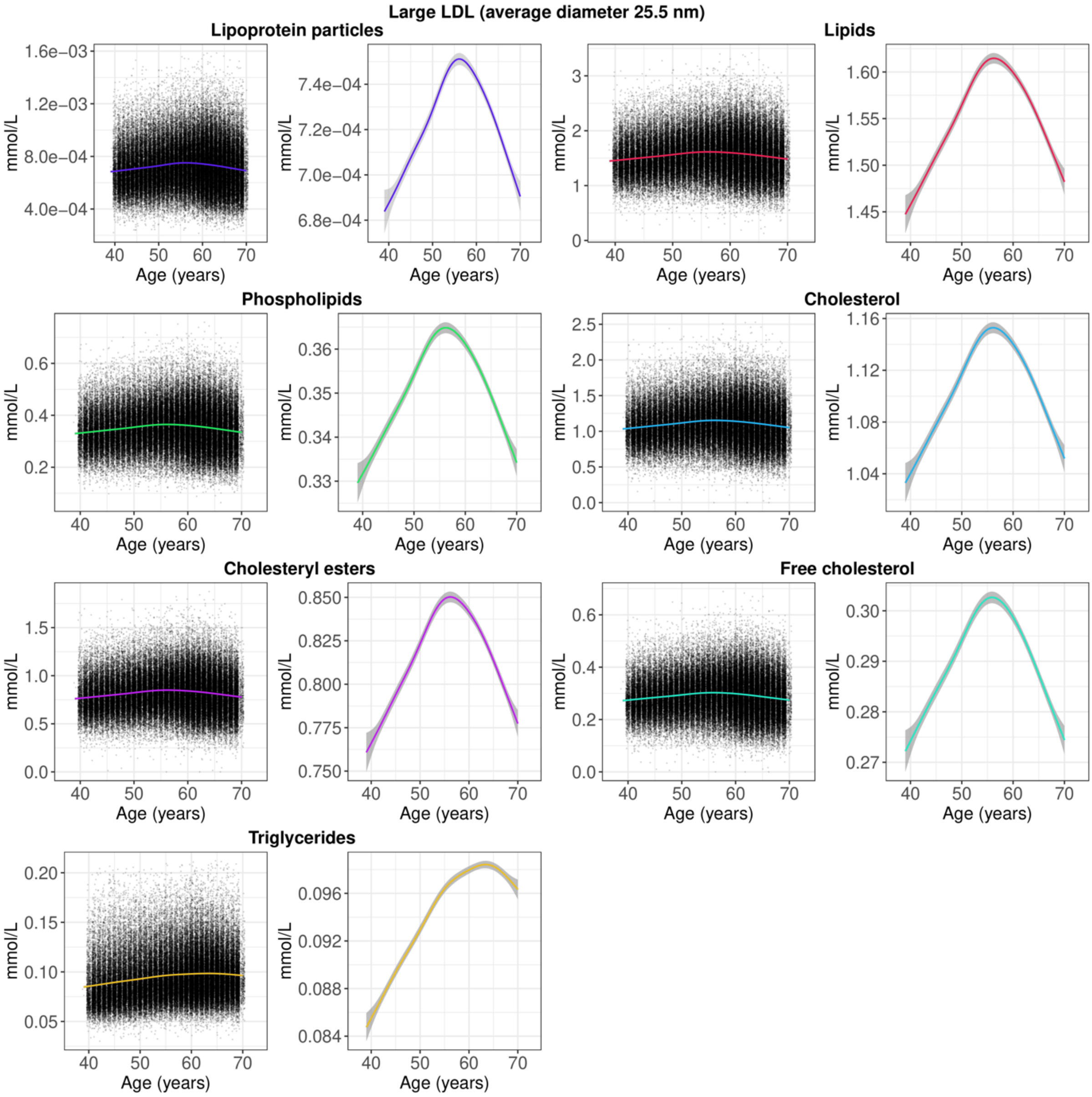

**Figure S27**. Distribution of large LDL (average diameter 25.5 nm) by chronological age, showing scatter plots of all observations and smooth curves (note the difference in y-axis scale). The smooth curves were estimated using generalised additive models, with shaded areas corresponding to 95% confidence intervals. LDL = low-density lipoprotein.

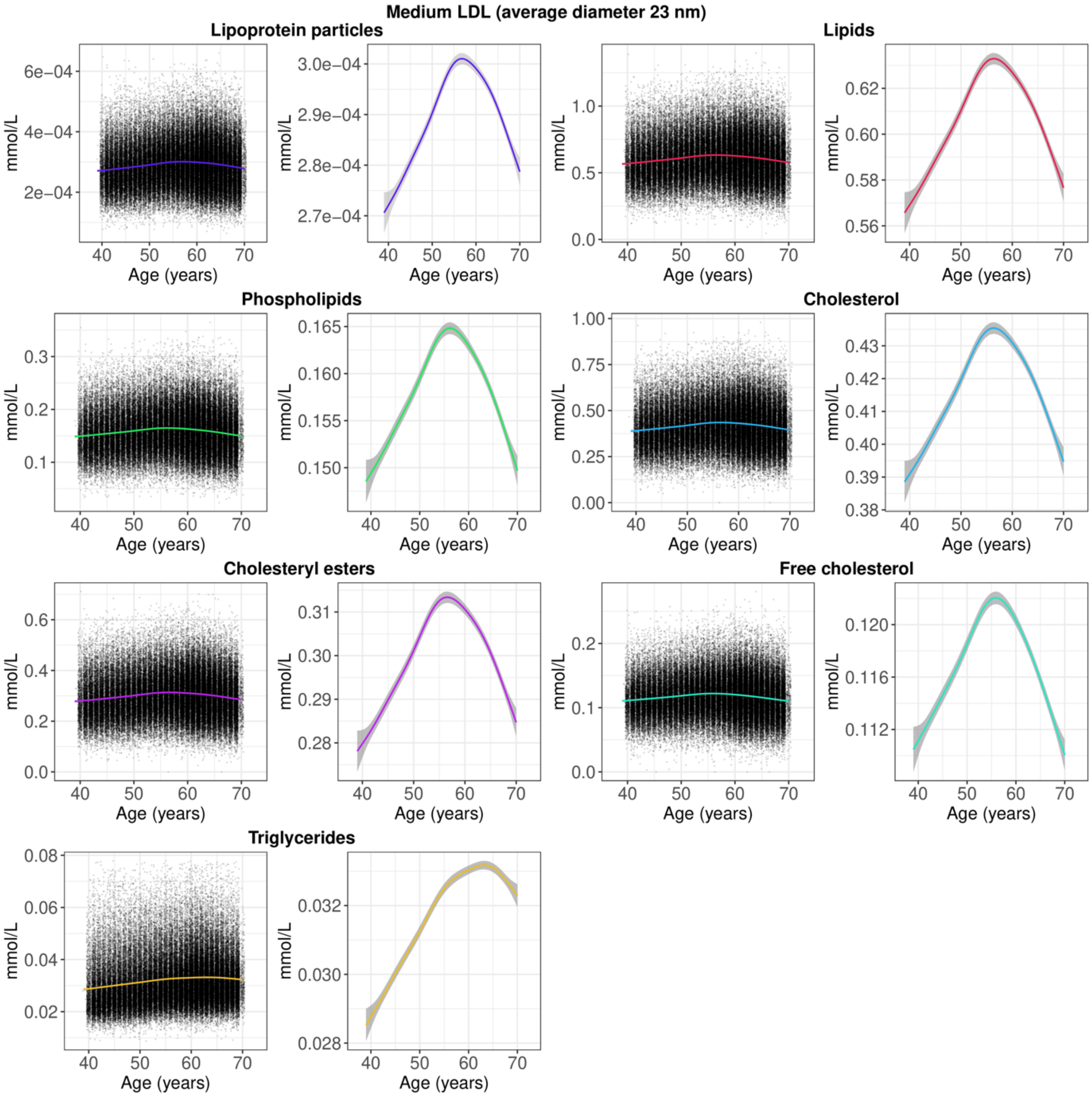

**Figure S28**. Distribution of medium LDL (average diameter 23 nm) by chronological age, showing scatter plots of all observations and smooth curves (note the difference in y-axis scale). The smooth curves were estimated using generalised additive models, with shaded areas corresponding to 95% confidence intervals. LDL = low-density lipoprotein.

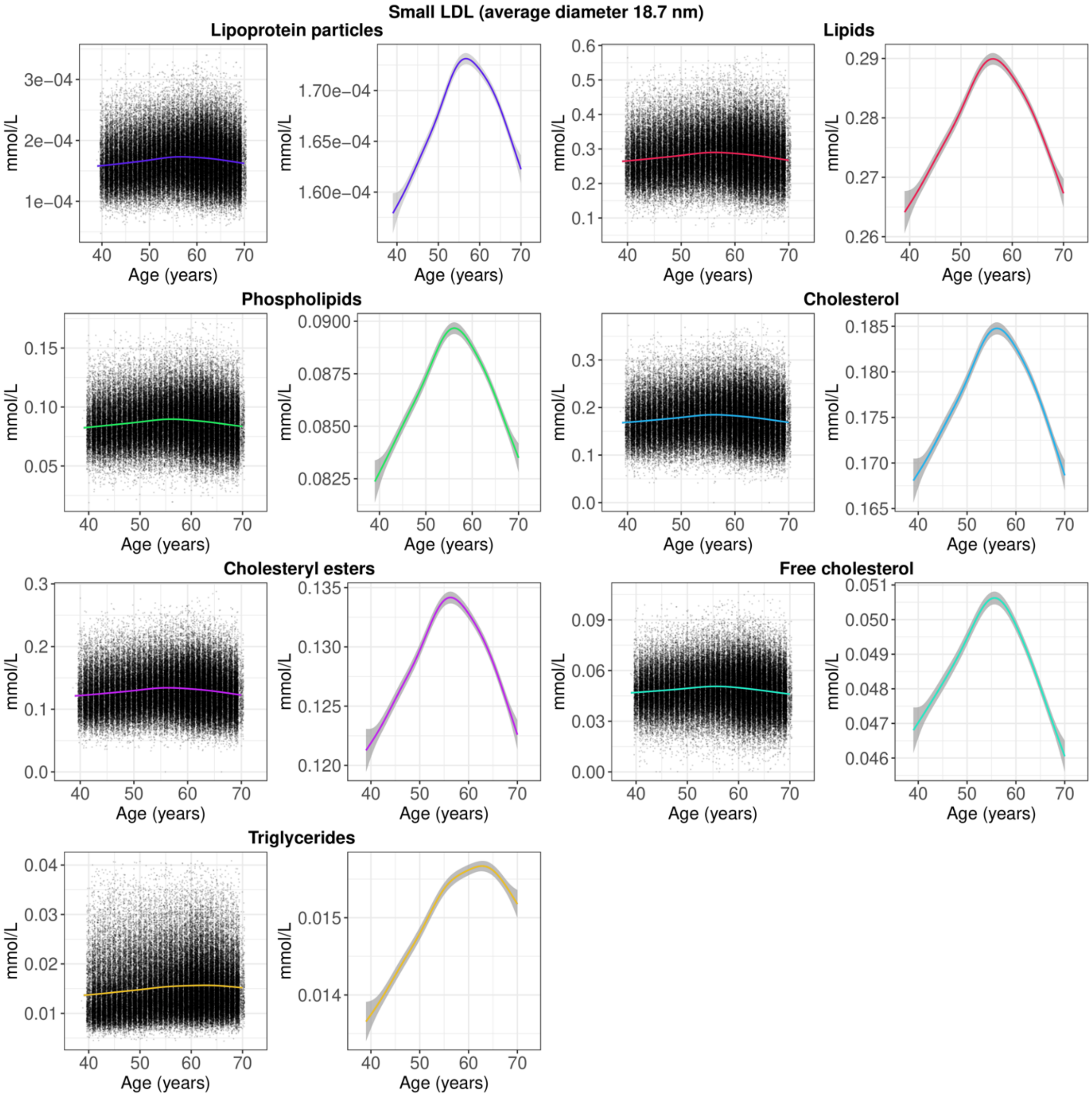

**Figure S29**. Distribution of small LDL (average diameter 18.7 nm) by chronological age, showing scatter plots of all observations and smooth curves (note the difference in y-axis scale). The smooth curves were estimated using generalised additive models, with shaded areas corresponding to 95% confidence intervals. LDL = low-density lipoprotein.

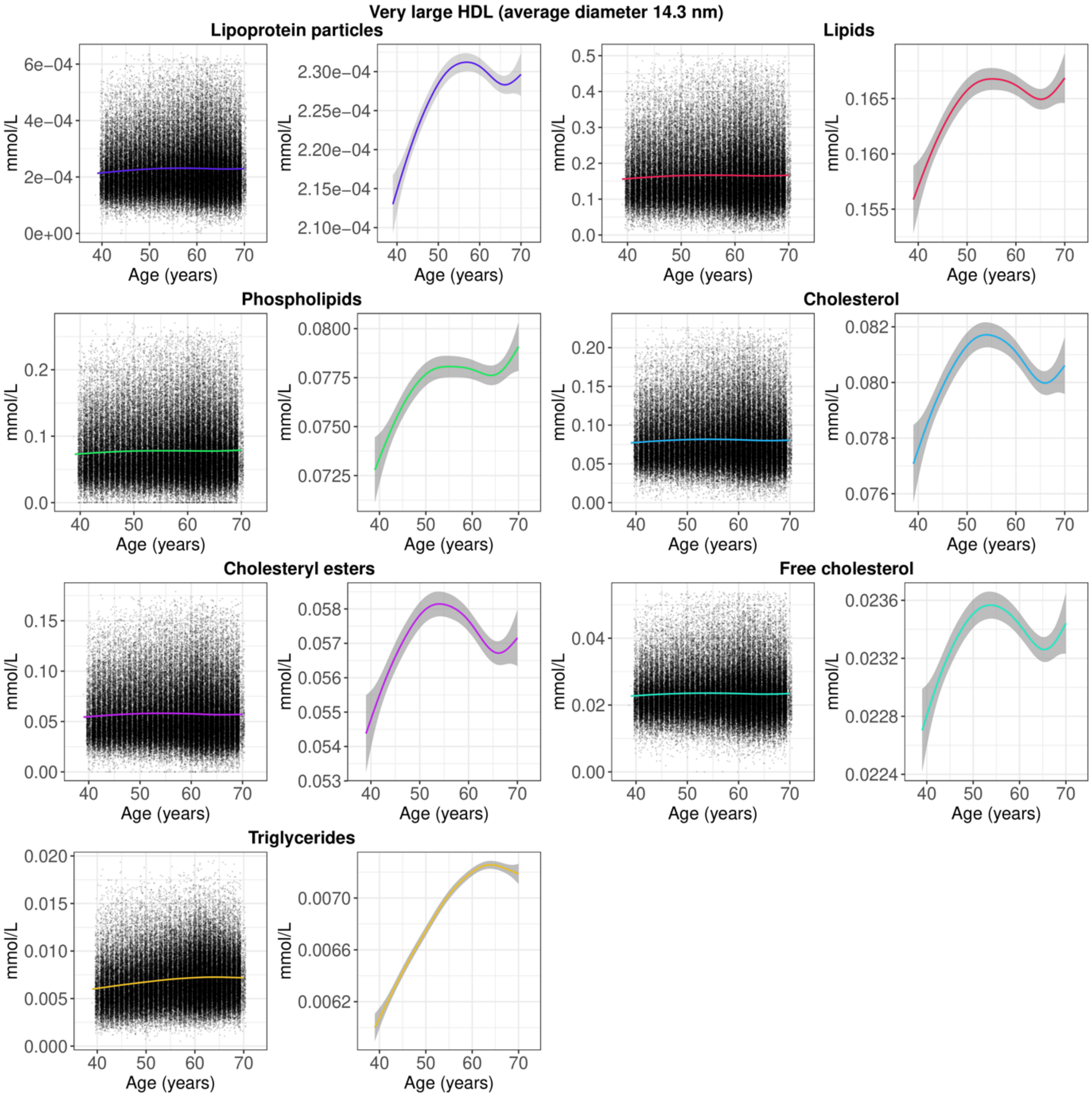

**Figure S30**. Distribution of very large HDL (average diameter 14.3 nm) by chronological age, showing scatter plots of all observations and smooth curves (note the difference in y-axis scale). The smooth curves were estimated using generalised additive models, with shaded areas corresponding to 95% confidence intervals. HDL = high-density lipoprotein.

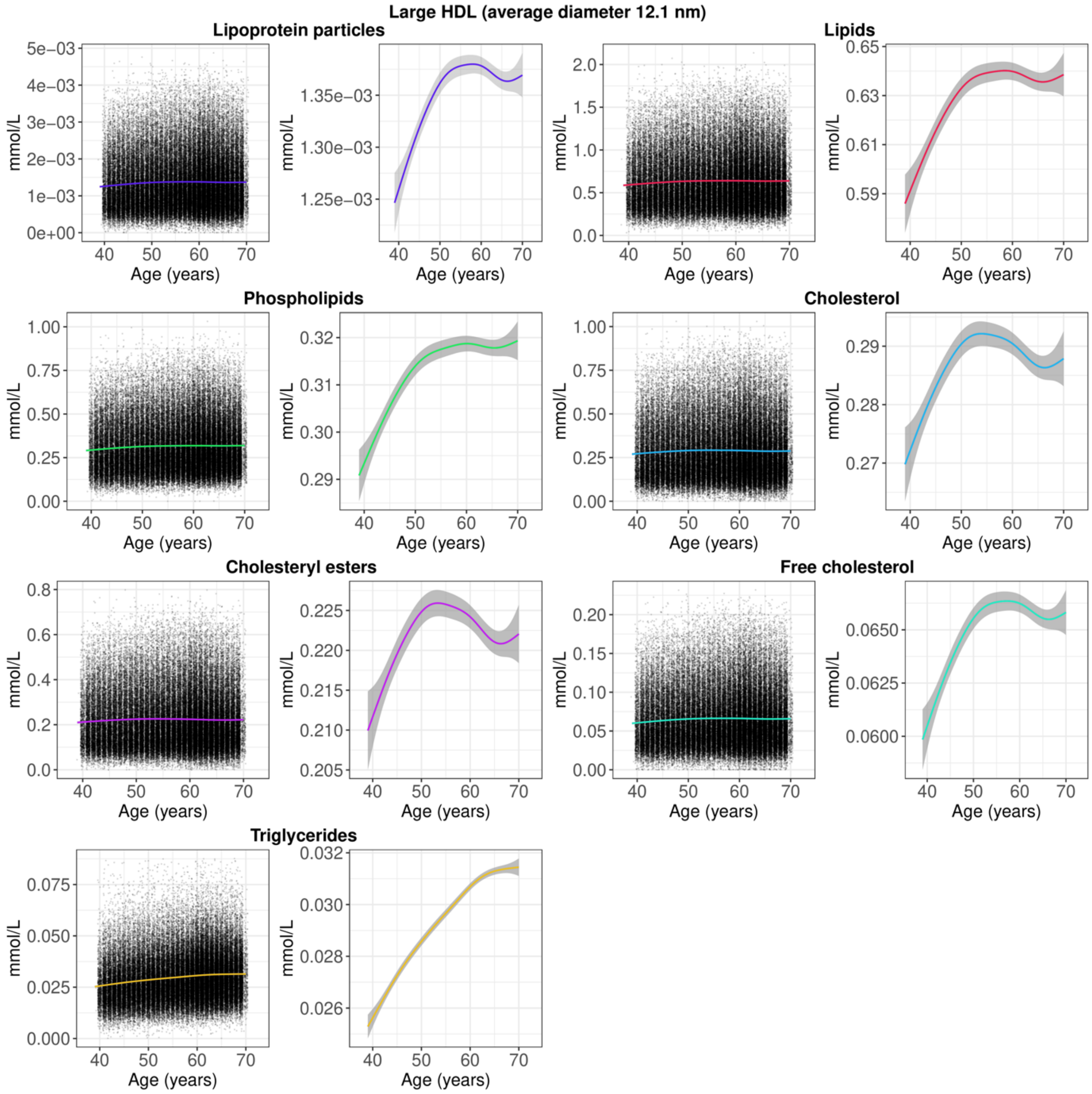

**Figure S31**. Distribution of large HDL (average diameter 12.1 nm) by chronological age, showing scatter plots of all observations and smooth curves (note the difference in y-axis scale). The smooth curves were estimated using generalised additive models, with shaded areas corresponding to 95% confidence intervals. HDL = high-density lipoprotein.

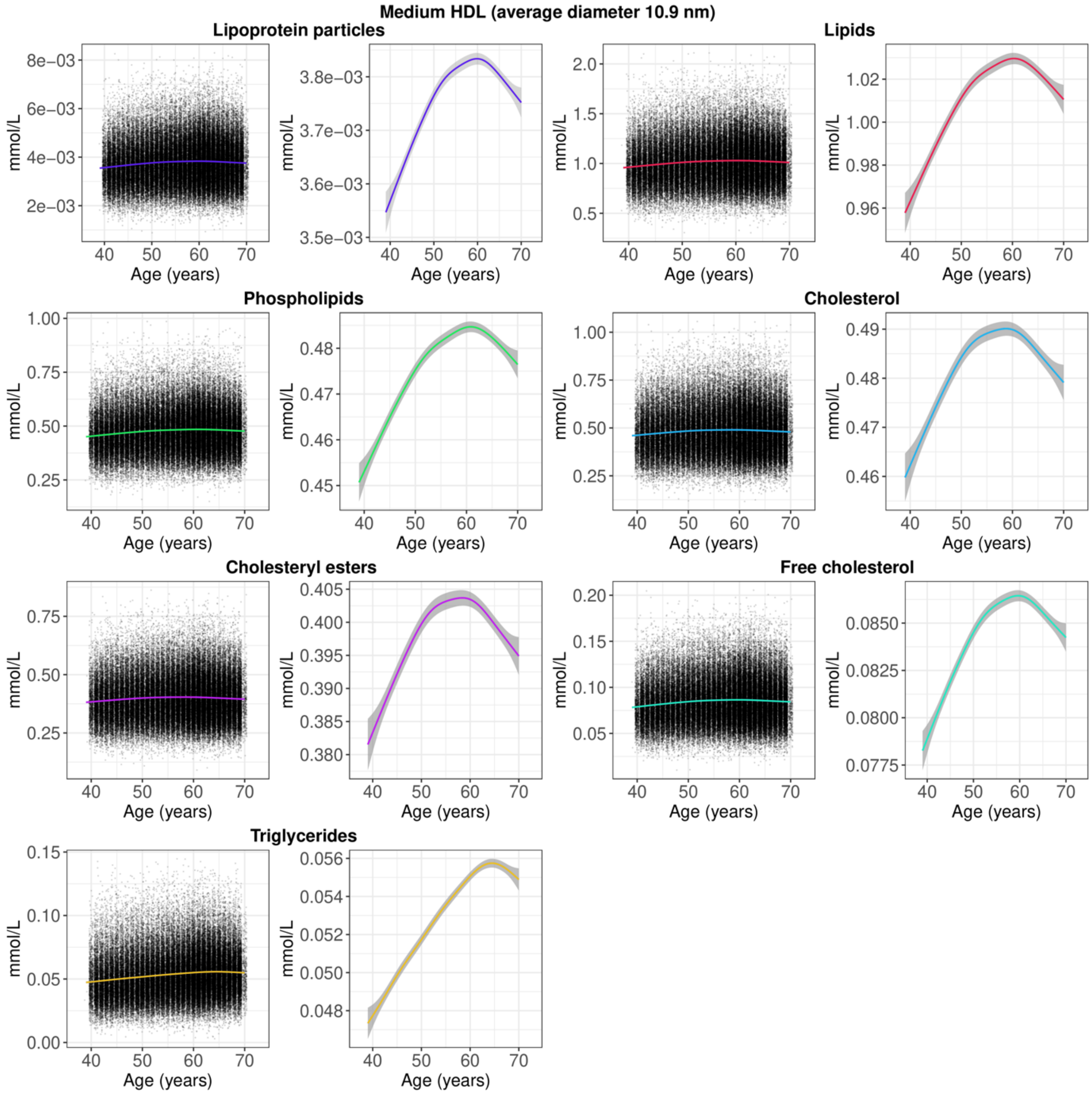

**Figure S32**. Distribution of medium HDL (average diameter 10.9 nm) by chronological age, showing scatter plots of all observations and smooth curves (note the difference in y-axis scale). The smooth curves were estimated using generalised additive models, with shaded areas corresponding to 95% confidence intervals. HDL = high-density lipoprotein.

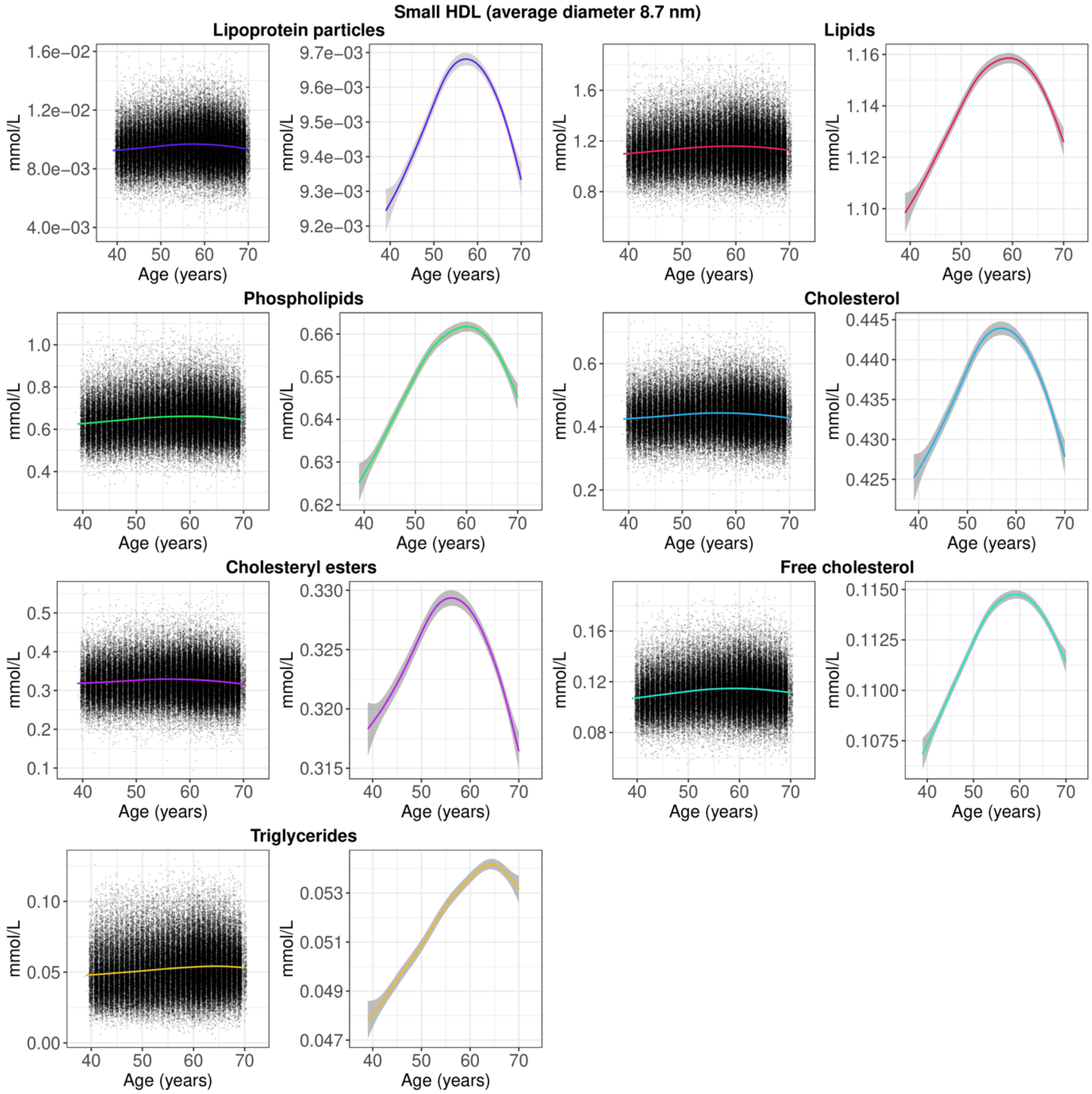

**Figure S33**. Distribution of small HDL (average diameter 8.7 nm) by chronological age, showing scatter plots of all observations and smooth curves (note the difference in y-axis scale). The smooth curves were estimated using generalised additive models, with shaded areas corresponding to 95% confidence intervals. HDL = high-density lipoprotein.

### 5. Metabolite-wide associations

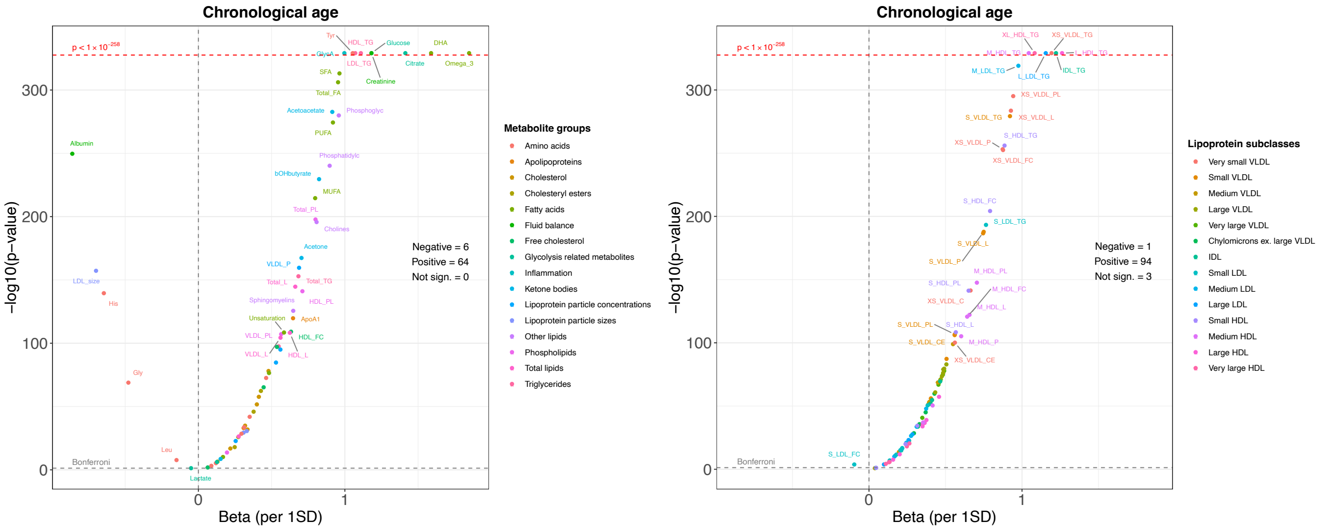

**Figure S34**. Volcano plots showing associations between chronological age and metabolite levels. Metabolites were standardised to have a mean equal to zero and a standard deviation of one. Associations were estimated using ordinary least squares regression. Models were adjusted for sex. *P*-values were -$\log_{10}$ transformed. *P*-values $< 1 \times 10^{-258}$ shown at the top of the plot could not be precisely estimated. SD = standard deviation; VLDL = very low-density lipoprotein; IDL = intermediate-density lipoprotein; LDL = low-density lipoprotein; HDL = high-density lipoprotein.

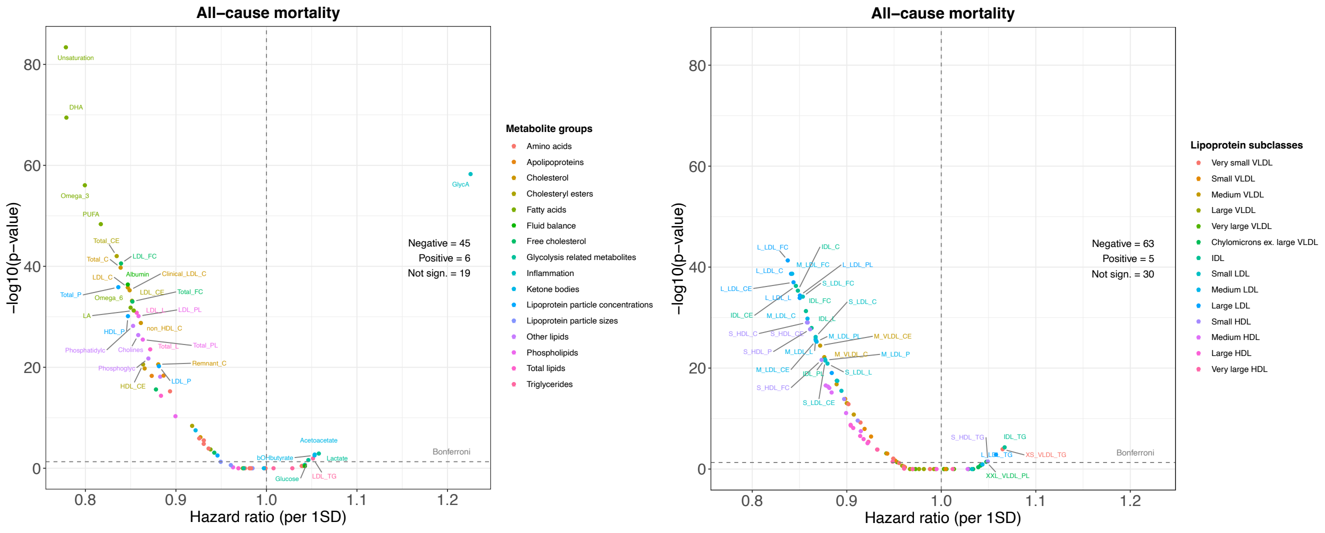

**Figure S35**. Volcano plots showing associations between metabolite levels and all-cause mortality. Metabolites were standardised to have a mean equal to zero and a standard deviation of one. Hazard ratios were estimated using Cox proportional hazards models adjusted for sex, with age (in years) as the underlying time axis. *P*-values were $-\log_{10}$ transformed. SD = standard deviation; VLDL = very low-density lipoprotein; IDL = intermediate-density lipoprotein; LDL = low-density lipoprotein; HDL = high-density lipoprotein.

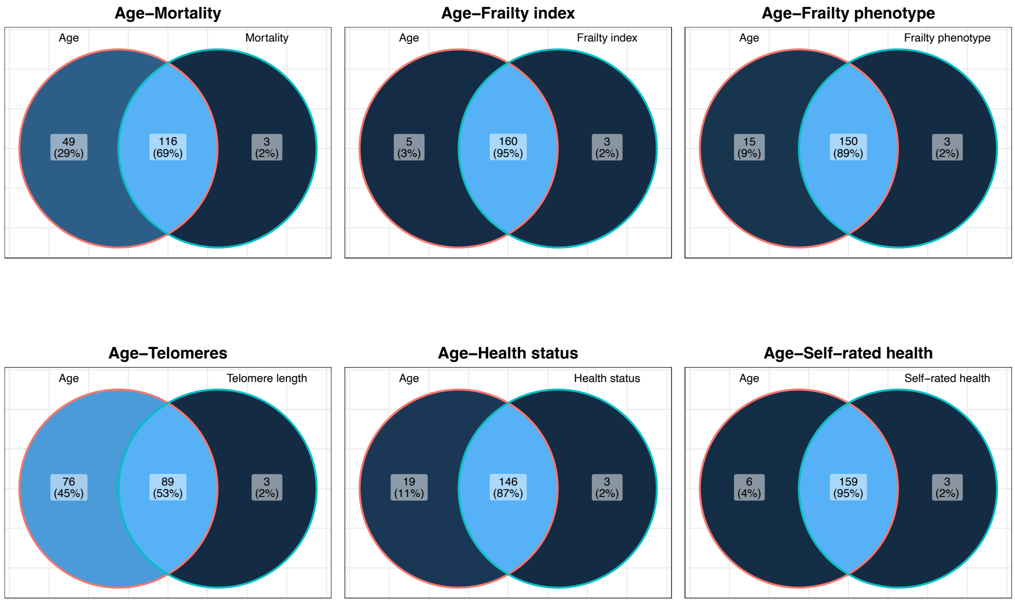

**Figure S36**. Venn diagrams showing the overlap between metabolites that were statistically significantly ($P < 0.05/168$) associated with chronological age and all-cause mortality or health indicators.

#### 6. Predictive model performance

**Table S3**. Predictive performance for all models with tuned hyperparameters

| | MAE | | | RMSE | | | $r$ | | | $R^2$ | | |
|---|---|---|---|---|---|---|---|---|---|---|---|---|
| Model | Train | Test | Diff | Train | Test | Diff | Train | Test | Diff | Train | Test | Diff |
| SVM radial | 5.05 | 5.31 | -0.268 | 6.35 | 6.60 | -0.247 | 0.62 | 0.59 | 0.037 | 0.39 | 0.35 | 0.045 |
| SVM polynomial | 5.22 | 5.35 | -0.135 | 6.52 | 6.63 | -0.117 | 0.60 | 0.58 | 0.018 | 0.36 | 0.34 | 0.022 |
| Cubist rules | 5.22 | 5.42 | -0.200 | 6.44 | 6.68 | -0.241 | 0.61 | 0.57 | 0.042 | 0.38 | 0.33 | 0.050 |
| RuleFit ensemble | 5.39 | 5.52 | -0.128 | 6.61 | 6.78 | -0.162 | 0.58 | 0.55 | 0.029 | 0.34 | 0.30 | 0.033 |
| XGBoost | 4.08 | 5.59 | -1.513 | 5.02 | 6.84 | -1.817 | 0.81 | 0.54 | 0.273 | 0.66 | 0.29 | 0.371 |
| BART | 5.50 | 5.62 | -0.122 | 6.70 | 6.86 | -0.154 | 0.57 | 0.54 | 0.030 | 0.32 | 0.29 | 0.033 |
| SVM linear | 5.64 | 5.65 | -0.010 | 6.93 | 6.94 | -0.008 | 0.53 | 0.52 | 0.001 | 0.28 | 0.28 | 0.001 |
| Elastic net | 5.69 | 5.70 | -0.005 | 6.91 | 6.92 | -0.006 | 0.52 | 0.52 | 0.001 | 0.27 | 0.27 | 0.001 |
| LASSO | 5.69 | 5.70 | -0.006 | 6.92 | 6.92 | -0.006 | 0.52 | 0.52 | 0.001 | 0.27 | 0.27 | 0.001 |
| Ridge | 5.90 | 5.91 | -0.003 | 7.11 | 7.11 | -0.003 | 0.48 | 0.48 | 0.001 | 0.24 | 0.23 | 0.001 |
| Random forest | 2.42 | 5.92 | -3.503 | 2.94 | 7.12 | -4.174 | 0.98 | 0.49 | 0.487 | 0.95 | 0.24 | 0.713 |
| Bagging | 4.15 | 5.94 | -1.782 | 5.00 | 7.15 | -2.141 | 0.86 | 0.48 | 0.377 | 0.73 | 0.23 | 0.504 |
| PLSR | 6.19 | 6.19 | 0.002 | 7.41 | 7.41 | 0.001 | 0.41 | 0.41 | 0.000 | 0.17 | 0.17 | 0.000 |
| Regression tree | 5.99 | 6.26 | -0.269 | 7.25 | 7.58 | -0.335 | 0.45 | 0.37 | 0.083 | 0.20 | 0.13 | 0.068 |
| KNN | 0.47 | 6.30 | -5.827 | 0.57 | 7.58 | -7.007 | 0.98 | 0.36 | 0.616 | 0.96 | 0.13 | 0.830 |
| MARS | 6.34 | 6.34 | 0.003 | 7.57 | 7.57 | 0.003 | 0.36 | 0.36 | -0.001 | 0.13 | 0.13 | -0.001 |
| MARS ensemble | 6.36 | 6.36 | -0.003 | 7.58 | 7.58 | -0.003 | 0.36 | 0.36 | 0.001 | 0.13 | 0.13 | 0.001 |

*Note:* MAE = mean absolute error; RMSE = root mean squared error; $r$ = correlation coefficient; $R^2$ = coefficient of determination. Models shown in ascending order by the lowest nested cross-validation MAE across the 10% hold-out test sets. See Panel 1 for model abbreviations.

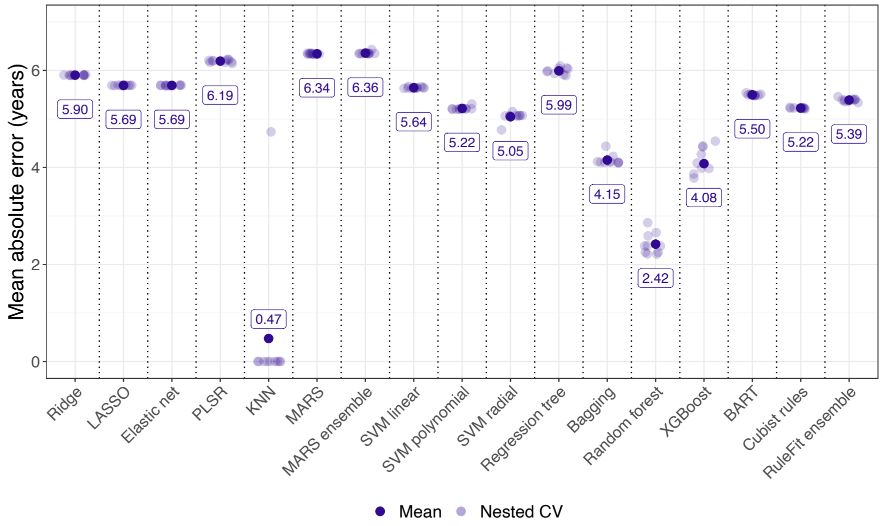

**Figure S37**. Nested cross-validation mean absolute error (MAE) for all models with tuned hyperparameter values in the 90% training sets. See Panel 1 for model abbreviations.

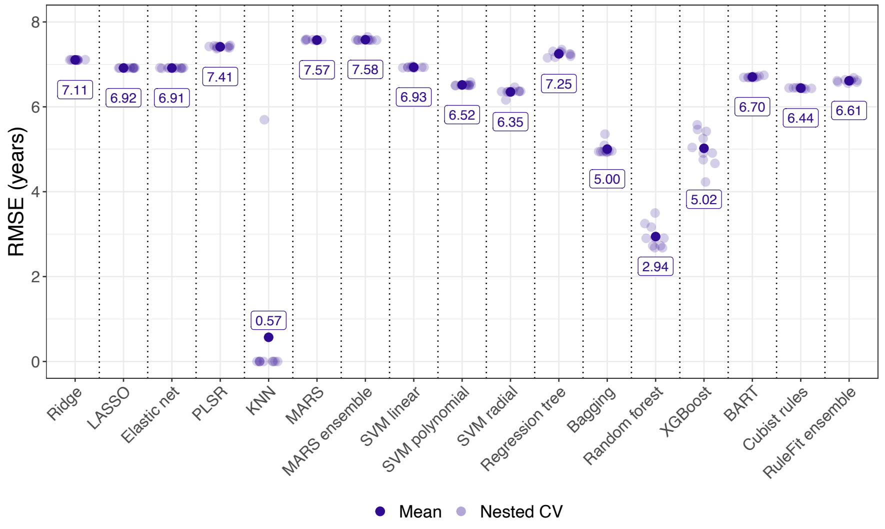

**Figure S38**. Nested cross-validation root-mean-square error (RMSE) for all models with tuned hyperparameter values in the 90% training sets. See Panel 1 for model abbreviations.

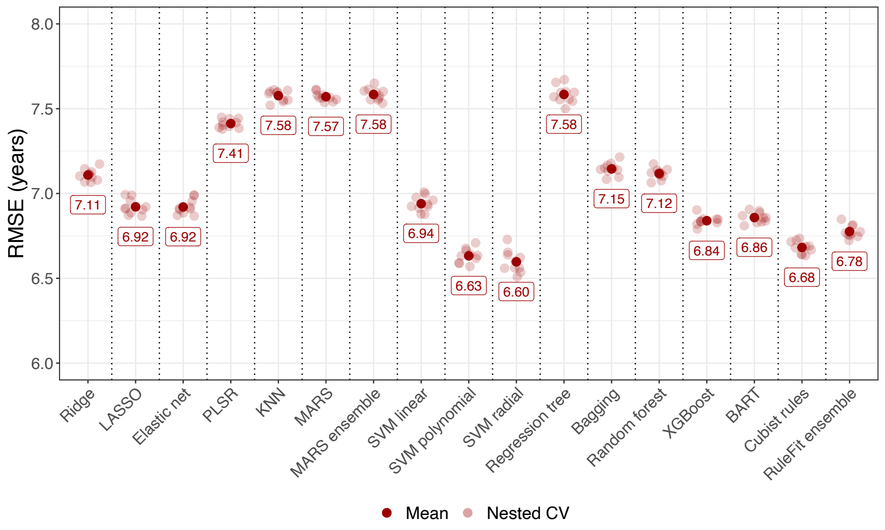

**Figure S39**. Nested cross-validation root-mean-square error (RMSE) for all models with tuned hyperparameter values in the 10% hold-out test sets. See Panel 1 for model abbreviations.

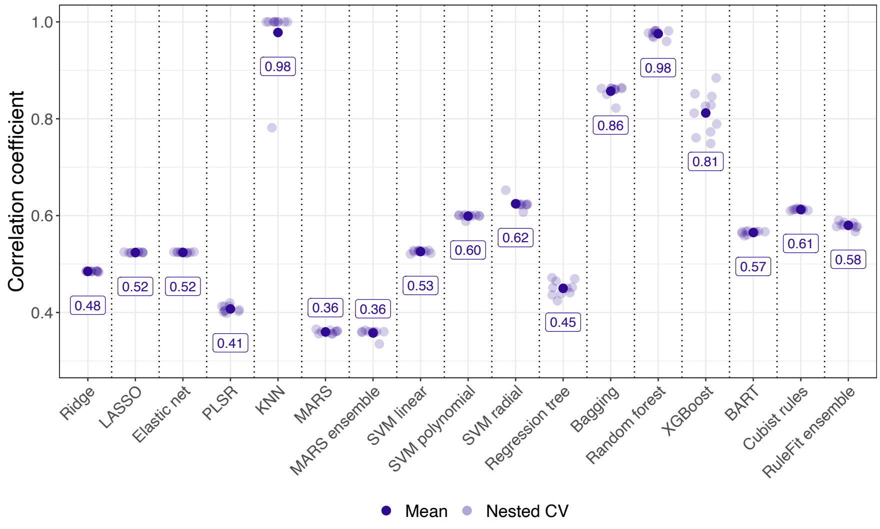

**Figure S40**. Nested cross-validation Pearson's correlation coefficient (*r*) for all models with tuned hyperparameter values in the 90% training sets. See Panel 1 for model abbreviations.

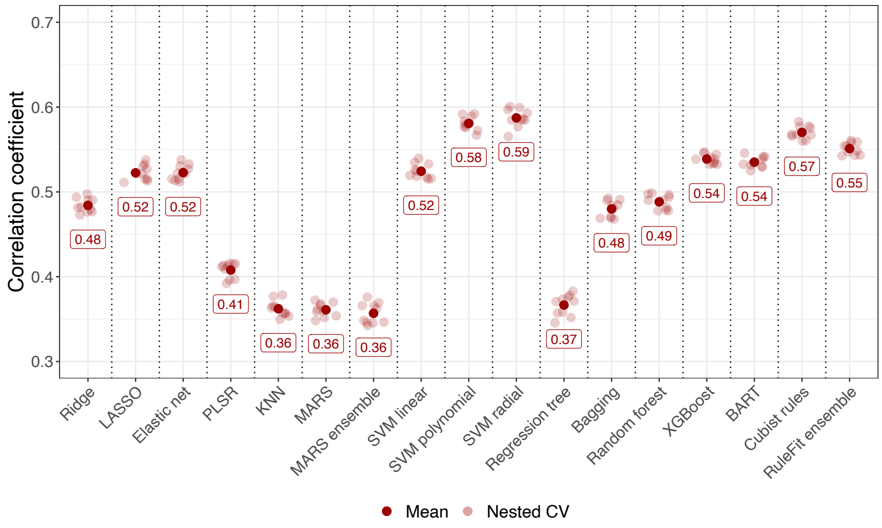

**Figure S41**. Nested cross-validation Pearson's correlation coefficient (*r*) for all models with tuned hyperparameter values in the 10% hold-out test sets. See Panel 1 for model abbreviations.

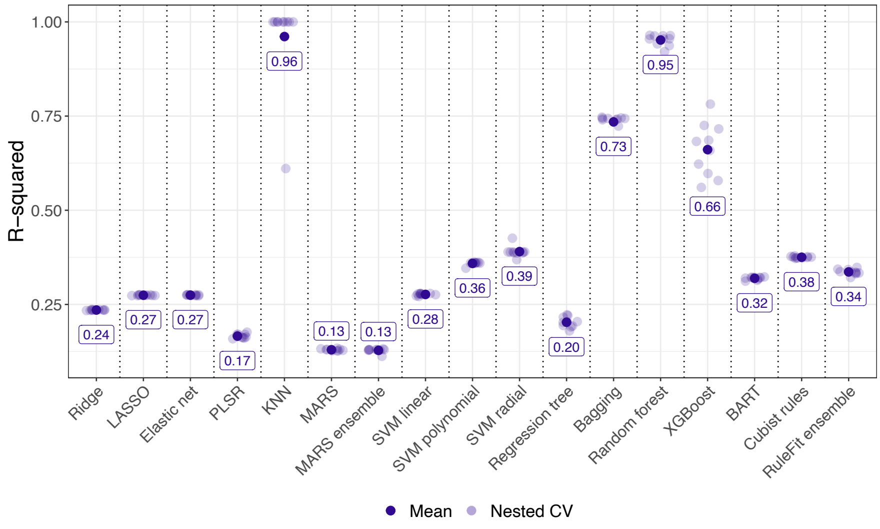

**Figure S42**. Nested cross-validation coefficient of determination ($R^2$) for all models with tuned hyperparameter values in the 90% training sets. See Panel 1 for model abbreviations.

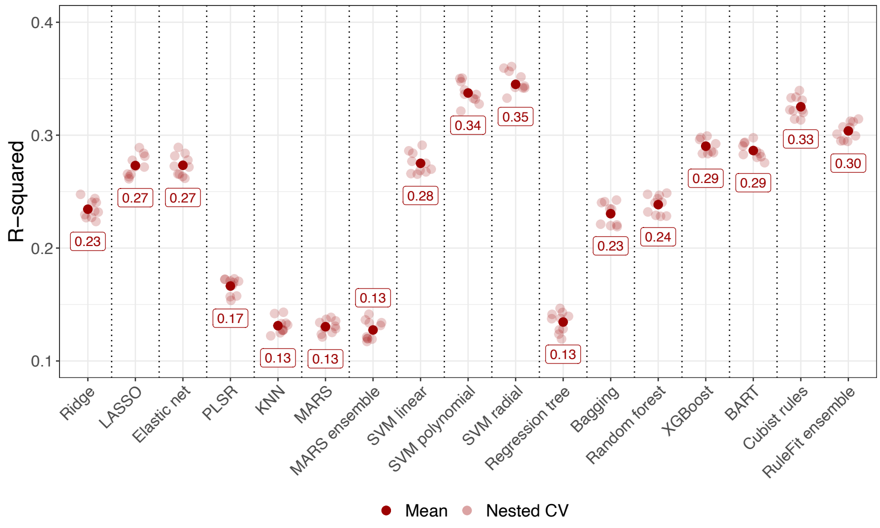

**Figure S43**. Nested cross-validation coefficient of determination ($R^2$) for all models with tuned hyperparameter values in the 10% hold-out test sets. See Panel 1 for model abbreviations.

### 7. Variable importance scores

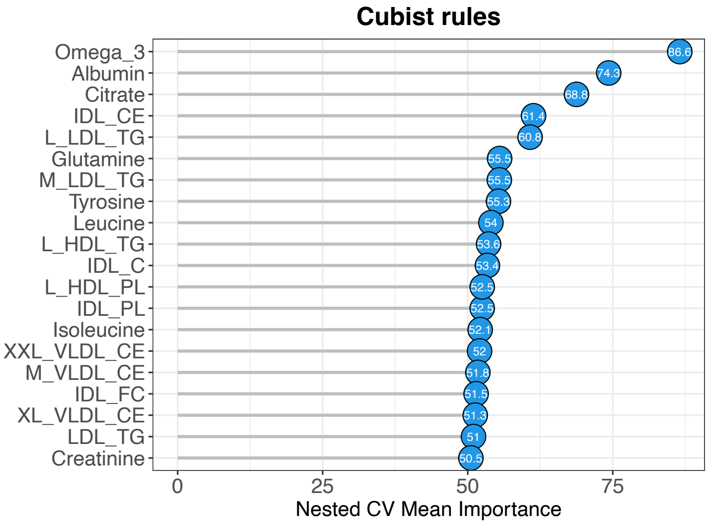

**Figure S44**. Variable importance plot, showing the top 20 metabolites ranked based on their influence on the Cubist rule-based regression model's predictive accuracy. Values shown represent averages across the 10 outer loops of the nested cross-validation. IDL = intermediate-density lipoprotein; CE = cholesteryl esters; LDL = low-density lipoprotein; TG = triglycerides; HDL = high-density lipoprotein; C = cholesterol; PL = phospholipids; VLDL = very low-density lipoprotein; FC = free cholesterol; CV = cross-validation.

#### 8. Correlation predicted and chronological age

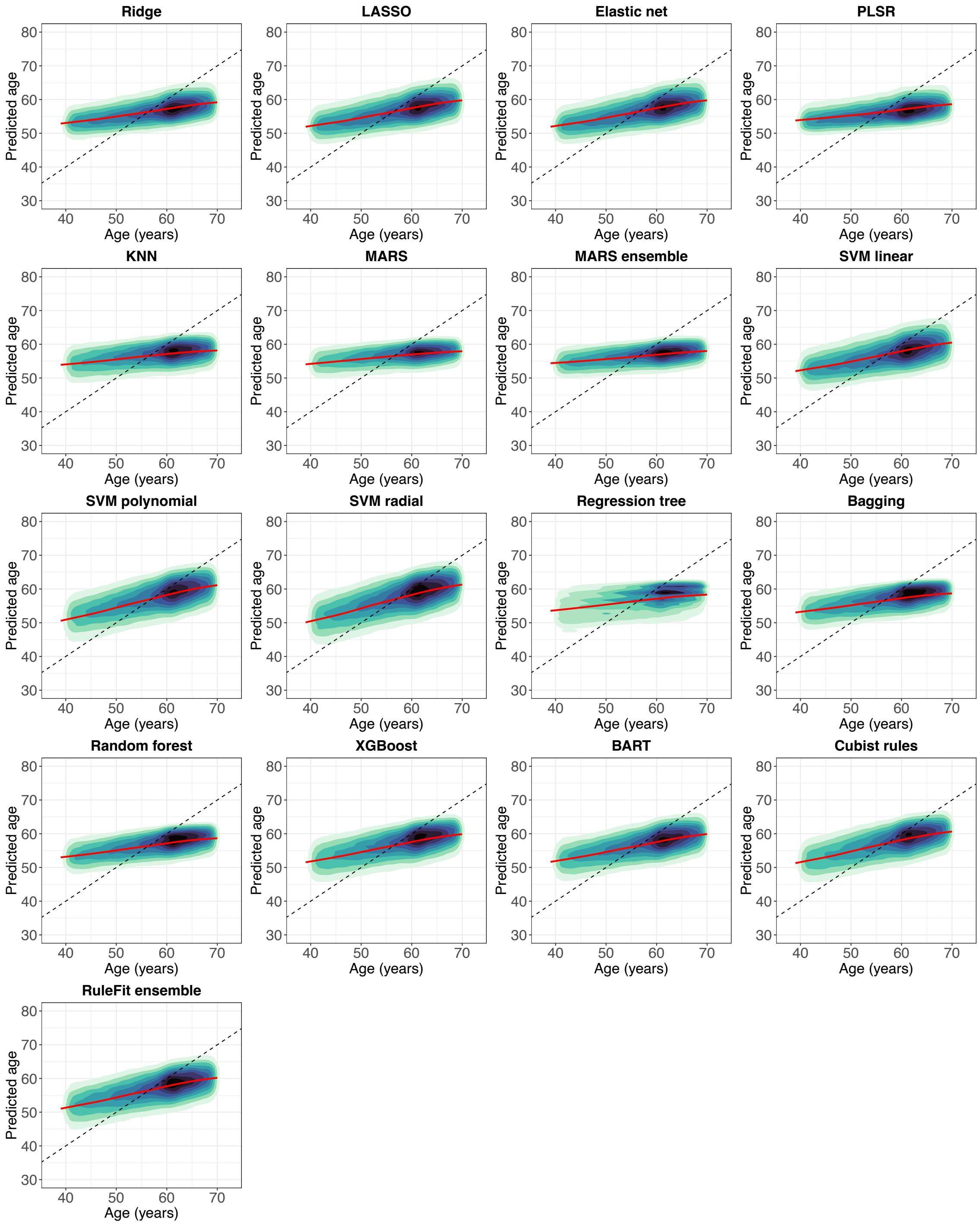

**Figure S45**. Scatter plots for all models showing the correlation between predicted age and chronological age before applying a statistical correction to the predicted age to remove the age bias (i.e., the systematic overestimation of age in young individuals and underestimation of age in older individuals). See Panel 1 for model abbreviations.

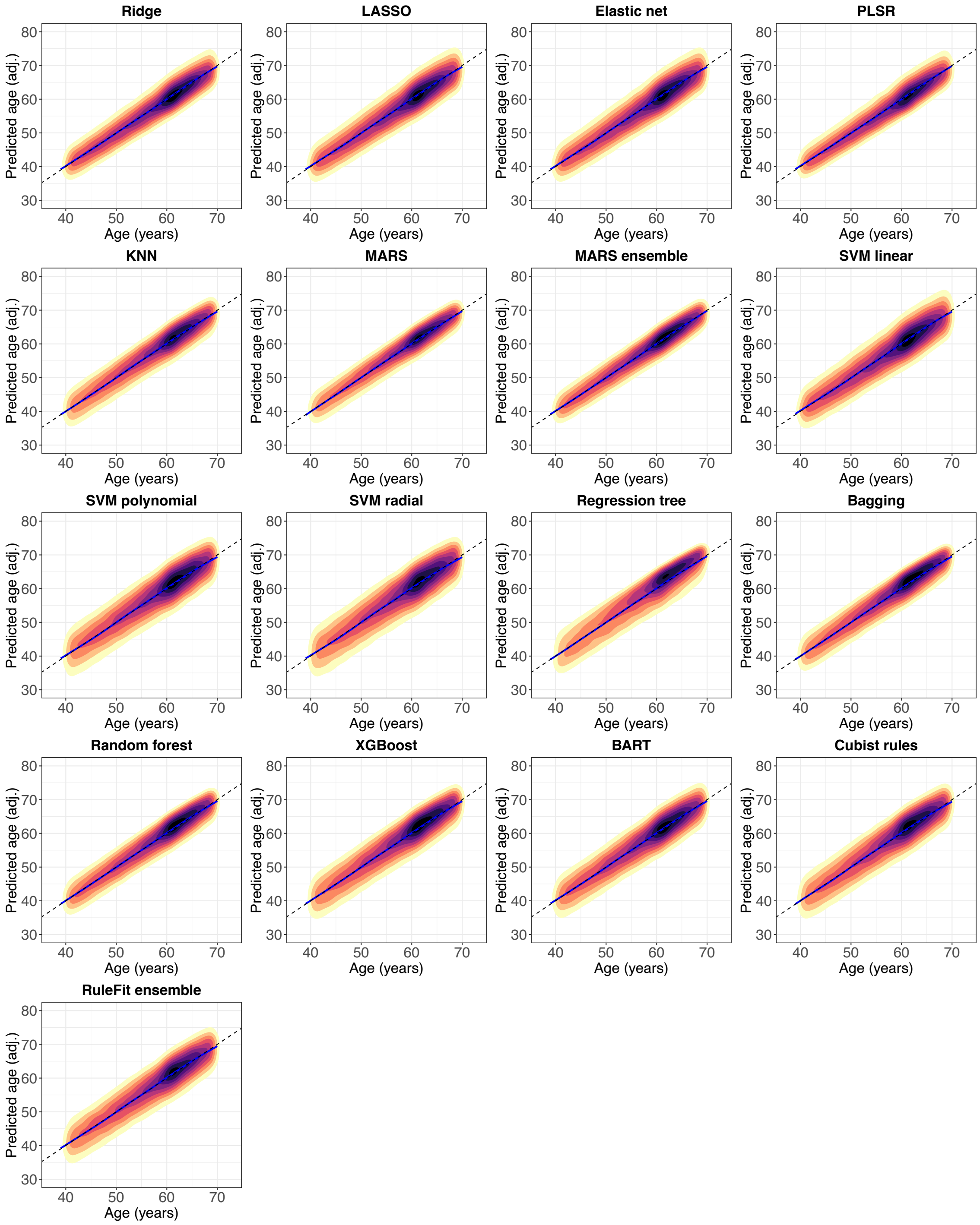

**Figure S46**. Scatter plots for all models showing the correlation between predicted age and chronological age after applying a statistical correction to the predicted age to remove the age bias (i.e., the systematic overestimation of age in young individuals and underestimation of age in older individuals). See Panel 1 for model abbreviations.

#### 9. Model extrapolation

**Table S4**. Model extrapolation beyond chronological age range (39 to 70 years)

| | Predicted age | | | | Extrapolation | | | |
|---|---|---|---|---|---|---|---|---|
| | MileAge | | MileAge (adj.) | | MileAge | | MileAge (adj.) | |
| Model | Min | Max | Min | Max | *n* < Min | *n* > Max | *n* < Min | *n* > Max |
| Ridge | 42.83 | 77.63 | 30.78 | 84.37 | 0 | 45 | 1694 | 3985 |
| LASSO | 37.37 | 77.76 | 27.57 | 82.30 | 2 | 111 | 2072 | 4533 |
| Elastic net | 37.36 | 77.68 | 27.78 | 82.37 | 2 | 107 | 2062 | 4535 |
| PLSR | 43.46 | 77.62 | 30.59 | 84.85 | 0 | 43 | 1397 | 3870 |
| KNN | 43.01 | 66.42 | 30.60 | 76.57 | 0 | 0 | 2240 | 3217 |
| MARS | 38.65 | 68.23 | 27.83 | 76.34 | 1 | 0 | 1921 | 2561 |
| MARS ensemble | 44.30 | 67.70 | 30.68 | 77.50 | 0 | 0 | 1408 | 2532 |
| SVM linear | 38.21 | 77.51 | 27.96 | 84.52 | 2 | 250 | 1887 | 4136 |
| SVM polynomial | 29.17 | 77.48 | 20.19 | 85.15 | 94 | 159 | 3379 | 4735 |
| SVM radial | 31.95 | 77.51 | 23.35 | 84.97 | 107 | 149 | 3570 | 4812 |
| Regression tree | 43.33 | 65.00 | 31.10 | 76.32 | 0 | 0 | 2727 | 3189 |
| Bagging | 45.30 | 64.63 | 32.60 | 74.83 | 0 | 0 | 2101 | 2683 |
| Random forest | 44.80 | 64.83 | 32.34 | 75.11 | 0 | 0 | 1928 | 2813 |
| XGBoost | 38.23 | 70.45 | 28.43 | 79.42 | 2 | 3 | 2777 | 3876 |
| BART | 38.23 | 74.63 | 27.26 | 82.23 | 2 | 29 | 2581 | 4165 |
| Cubist rules | 38.64 | 74.82 | 27.63 | 80.15 | 2 | 32 | 2841 | 4035 |
| RuleFit ensemble | 36.93 | 75.90 | 26.28 | 81.77 | 24 | 48 | 2870 | 4485 |

*Note:* Min = minimum; Max = maximum. See Panel 1 for model abbreviations.

#### 10. Predictive model performance after age-bias correction

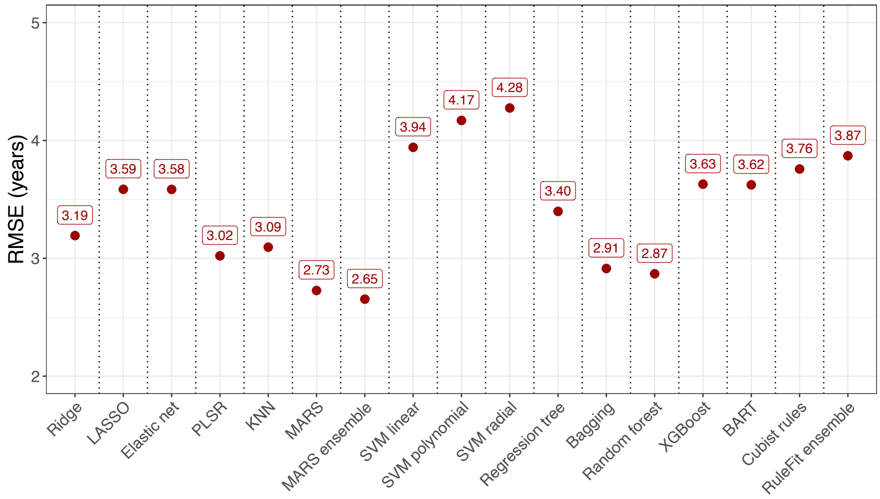

**Figure S47**. Root-mean-square error (RMSE) for all models with tuned hyperparameter values calculated in the nested cross-validation dataset after applying a statistical correction to the predicted age to remove the age bias (i.e., the systematic overestimation of age in young individuals and underestimation of age in older individuals). See Panel 1 for model abbreviations.

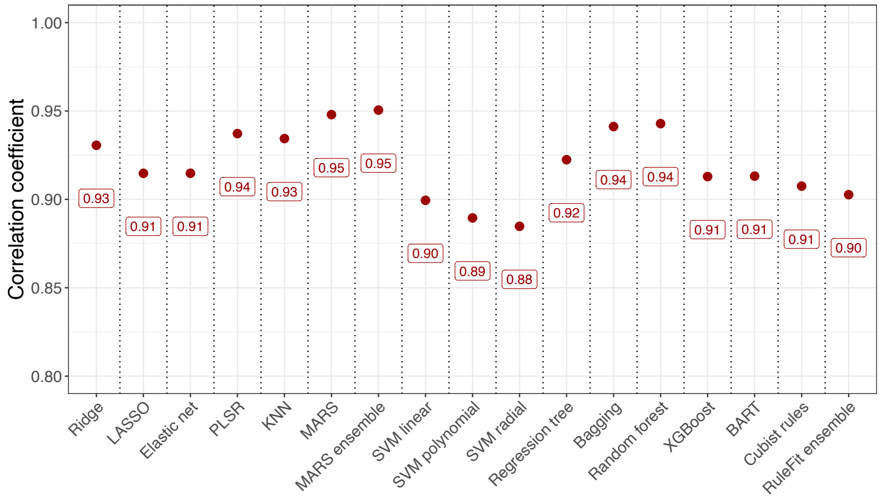

**Figure S48**. Pearson's correlation coefficient (*r*) for all models with tuned hyperparameter values calculated in the nested cross-validation dataset after applying a statistical correction to the predicted age to remove the age bias (i.e., the systematic overestimation of age in young individuals and underestimation of age in older individuals). See Panel 1 for model abbreviations.

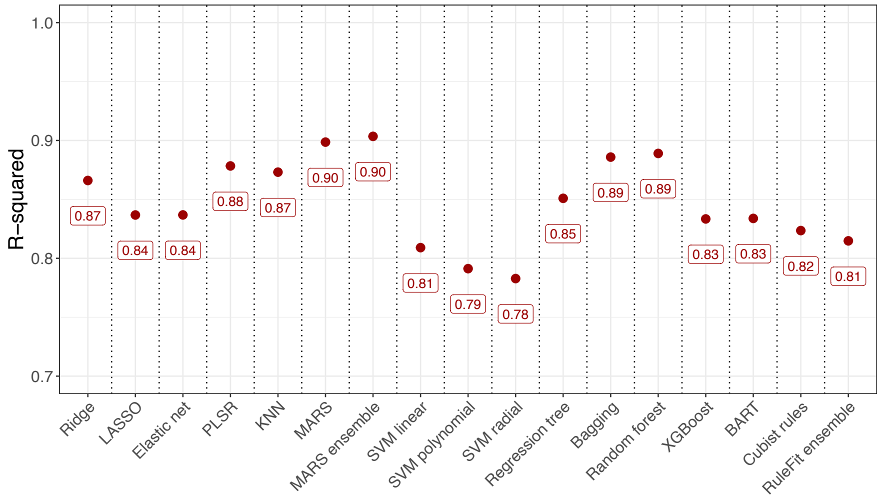

**Figure S49**. Coefficient of determination ($R^2$) for all models with tuned hyperparameter values calculated in the nested cross-validation dataset after applying a statistical correction to the predicted age to remove the age bias (i.e., the systematic overestimation of age in young individuals and underestimation of age in older individuals). See Panel 1 for model abbreviations.

#### 11. Descriptive statistics MileAge delta

**Table S5**. Descriptive statistics age delta before and after age-bias correction

| | MileAge delta | | | | | MileAge delta (adj.) | | | | |
|---|---|---|---|---|---|---|---|---|---|---|
| Model | Min | Med | Mean | Max | IQR | Min | Med | Mean | Max | IQR |
| Ridge | -21.29 | -0.62 | 0.00 | 30.10 | 10.74 | -12.01 | -0.14 | 0.00 | 20.20 | 4.20 |
| LASSO | -24.52 | -0.46 | 0.00 | 29.71 | 10.21 | -15.68 | -0.10 | 0.00 | 20.09 | 4.73 |
| Elastic net | -24.50 | -0.46 | 0.00 | 29.50 | 10.21 | -15.71 | -0.10 | 0.00 | 20.01 | 4.74 |
| PLSR | -20.54 | -0.74 | 0.00 | 30.08 | 11.37 | -10.41 | -0.20 | 0.00 | 21.25 | 3.94 |
| KNN | -21.63 | -0.66 | 0.06 | 23.58 | 11.51 | -11.79 | 0.17 | 0.00 | 10.90 | 4.27 |
| MARS | -22.62 | -0.85 | 0.00 | 24.04 | 11.68 | -16.17 | 0.16 | 0.00 | 11.07 | 3.63 |
| MARS ensemble | -21.46 | -0.88 | 0.00 | 25.87 | 11.75 | -11.58 | 0.09 | 0.00 | 11.49 | 3.62 |
| SVM linear | -24.60 | -0.02 | 0.41 | 32.11 | 10.13 | -17.03 | -0.12 | 0.00 | 20.04 | 5.19 |
| SVM polynomial | -28.12 | -0.01 | 0.37 | 30.68 | 9.38 | -23.22 | 0.16 | 0.00 | 19.78 | 5.52 |
| SVM radial | -28.83 | -0.03 | 0.30 | 27.87 | 9.17 | -22.03 | 0.17 | 0.00 | 18.07 | 5.73 |
| Regression tree | -26.67 | -0.73 | 0.01 | 25.00 | 11.29 | -15.35 | 0.42 | 0.00 | 11.27 | 4.94 |
| Bagging | -23.12 | -0.69 | 0.02 | 22.78 | 10.86 | -12.24 | 0.20 | 0.00 | 10.35 | 4.10 |
| Random forest | -23.21 | -0.78 | -0.07 | 23.14 | 10.83 | -12.39 | 0.14 | 0.00 | 10.69 | 4.01 |
| XGBoost | -25.36 | -0.51 | 0.00 | 25.06 | 9.89 | -15.84 | 0.19 | 0.00 | 14.85 | 4.95 |
| BART | -24.34 | -0.46 | 0.01 | 28.67 | 9.96 | -16.23 | 0.09 | 0.00 | 16.90 | 4.86 |
| Cubist rules | -27.06 | -0.01 | 0.41 | 27.14 | 9.55 | -18.94 | 0.14 | 0.00 | 16.50 | 5.10 |
| RuleFit ensemble | -25.02 | -0.41 | 0.00 | 26.10 | 9.68 | -18.69 | 0.12 | 0.00 | 16.77 | 5.19 |

*Note:* Min = minimum; Med = median; Max = maximum; IQR = interquartile range. See Panel 1 for model abbreviations.

#### 12. Sample characteristics health indicators

**Table S6**. Health indicators by MileAge delta (adj.) for the Cubist rules model

| | | MileAge delta (adj.) | | |
|---|---|---|---|---|
| | Full sample (*N*=101359) | ≤ 1SD below the mean (*n*=16204) | Middle (*n*=69166) | ≥ 1SD above the mean (*n*=15989) |
| Age | | | | |
| Chronological age | 56.44 (8.12) | 55.85 (8.33) | 56.75 (8.08) | 55.69 (8.00) |
| Predicted age | 56.85 (4.57) | 50.77 (3.24) | 57.04 (3.35) | 62.16 (2.80) |
| Predicted age (adj.) | 56.44 (8.95) | 49.96 (8.54) | 56.84 (8.39) | 61.24 (7.93) |
| MileAge delta (adj.) | 0.00 (3.76) | -5.89 (1.79) | 0.10 (2.04) | 5.55 (1.53) |
| Health indicators | | | | |
| Telomere length[1] | 0.00 (1.00) | -0.03 (0.99) | 0.00 (1.00) | 0.01 (1.00) |
| Frailty index[2] | 0.12 (0.07) | 0.11 (0.07) | 0.12 (0.07) | 0.14 (0.08) |
| Frailty phenotype | | | | |
| Non-frail | 50531 (49.9%) | 8344 (51.5%) | 34902 (50.5%) | 7285 (45.6%) |
| Pre-frail | 41718 (41.2%) | 6561 (40.5%) | 28195 (40.8%) | 6962 (43.5%) |
| Frail | 2812 (2.8%) | 340 (2.1%) | 1805 (2.6%) | 667 (4.2%) |
| Missing | 6298 (6.2%) | 959 (5.9%) | 4264 (6.2%) | 1075 (6.7%) |
| Long-standing illness | | | | |
| No | 67462 (66.6%) | 11502 (71.0%) | 46568 (67.3%) | 9392 (58.7%) |
| Yes | 31288 (30.9%) | 4326 (26.7%) | 20794 (30.1%) | 6168 (38.6%) |
| Missing | 2609 (2.6%) | 376 (2.3%) | 1804 (2.6%) | 429 (2.7%) |
| Health status | | | | |
| Healthy | 67275 (66.4%) | 11496 (70.9%) | 46331 (67.0%) | 9448 (59.1%) |
| Unhealthy | 31545 (31.1%) | 4287 (26.5%) | 21086 (30.5%) | 6172 (38.6%) |
| Missing | 2539 (2.5%) | 421 (2.6%) | 1749 (2.5%) | 369 (2.3%) |
| Self-rated health | | | | |
| Excellent | 16717 (16.5%) | 3060 (18.9%) | 11493 (16.6%) | 2164 (13.5%) |
| Good | 59059 (58.3%) | 9662 (59.6%) | 40730 (58.9%) | 8667 (54.2%) |
| Fair | 20860 (20.6%) | 2909 (18.0%) | 14001 (20.2%) | 3950 (24.7%) |
| Poor | 4153 (4.1%) | 482 (3.0%) | 2570 (3.7%) | 1101 (6.9%) |
| Missing | 570 (0.6%) | 91 (0.6%) | 372 (0.5%) | 107 (0.7%) |

*Note:* SD = standard deviation. MileAge delta (adj.) derived from the Cubist rule-based regression model. Numbers correspond to mean (standard deviation) or count (percentage). [1]*n* = 3395 missing. [2]*n* = 330 missing.

#### 13. Associations between MileAge delta and health indicators

**Table S7**. Association between frailty index scores and MileAge delta (adj.)

| Model | Level | *β* | 95% CI | | *p* |
|---|---|---|---|---|---|
| SVM radial | ≤ X̄-1SD | | Reference | | |
| | Middle | 0.006 | 0.005 | 0.007 | <0.001 |
| | ≥ X̄+1SD | 0.020 | 0.019 | 0.022 | <0.001 |
| SVM polynomial | ≤ X̄-1SD | | Reference | | |
| | Middle | 0.006 | 0.005 | 0.007 | <0.001 |
| | ≥ X̄+1SD | 0.021 | 0.020 | 0.023 | <0.001 |
| Cubist rules | ≤ X̄-1SD | | Reference | | |
| | Middle | 0.006 | 0.005 | 0.008 | <0.001 |
| | ≥ X̄+1SD | 0.023 | 0.021 | 0.024 | <0.001 |
| RuleFit ensemble | ≤ X̄-1SD | | Reference | | |
| | Middle | 0.005 | 0.003 | 0.006 | <0.001 |
| | ≥ X̄+1SD | 0.019 | 0.017 | 0.021 | <0.001 |
| XGBoost | ≤ X̄-1SD | | Reference | | |
| | Middle | 0.007 | 0.006 | 0.009 | <0.001 |
| | ≥ X̄+1SD | 0.021 | 0.019 | 0.023 | <0.001 |
| BART | ≤ X̄-1SD | | Reference | | |
| | Middle | 0.006 | 0.005 | 0.008 | <0.001 |
| | ≥ X̄+1SD | 0.020 | 0.019 | 0.022 | <0.001 |
| SVM linear | ≤ X̄-1SD | | Reference | | |
| | Middle | 0.006 | 0.004 | 0.007 | <0.001 |
| | ≥ X̄+1SD | 0.021 | 0.019 | 0.023 | <0.001 |
| Elastic net | ≤ X̄-1SD | | Reference | | |
| | Middle | 0.005 | 0.004 | 0.006 | <0.001 |
| | ≥ X̄+1SD | 0.019 | 0.017 | 0.021 | <0.001 |
| LASSO | ≤ X̄-1SD | | Reference | | |
| | Middle | 0.005 | 0.004 | 0.006 | <0.001 |
| | ≥ X̄+1SD | 0.019 | 0.018 | 0.021 | <0.001 |
| Ridge | ≤ X̄-1SD | | Reference | | |
| | Middle | 0.006 | 0.005 | 0.007 | <0.001 |
| | ≥ X̄+1SD | 0.019 | 0.018 | 0.021 | <0.001 |
| Random forest | ≤ X̄-1SD | | Reference | | |
| | Middle | 0.010 | 0.009 | 0.011 | <0.001 |
| | ≥ X̄+1SD | 0.019 | 0.018 | 0.021 | <0.001 |
| Bagging | ≤ X̄-1SD | | Reference | | |
| | Middle | 0.009 | 0.008 | 0.01 | <0.001 |
| | ≥ X̄+1SD | 0.02 | 0.019 | 0.022 | <0.001 |
| PLSR | ≤ X̄-1SD | | Reference | | |
| | Middle | 0.008 | 0.007 | 0.009 | <0.001 |
| | ≥ X̄+1SD | 0.020 | 0.018 | 0.021 | <0.001 |
| Regression tree | ≤ X̄-1SD | | Reference | | |
| | Middle | 0.005 | 0.004 | 0.006 | <0.001 |
| | ≥ X̄+1SD | 0.017 | 0.015 | 0.018 | <0.001 |
| KNN | ≤ X̄-1SD | | Reference | | |
| | Middle | 0.008 | 0.007 | 0.009 | <0.001 |
| | ≥ X̄+1SD | 0.020 | 0.018 | 0.021 | <0.001 |

| | | | | | |
|---|---|---|---|---|---|
| MARS | ≤ $\bar{X}$-1SD | Reference | | | |
| | Middle | 0.001 | 0.001 | 0.003 | 0.025 |
| | ≥ $\bar{X}$+1SD | 0.008 | 0.006 | 0.01 | <0.001 |
| MARS ensemble | ≤ $\bar{X}$-1SD | Reference | | | |
| | Middle | 0.002 | 0.001 | 0.003 | 0.003 |
| | ≥ $\bar{X}$+1SD | 0.012 | 0.011 | 0.014 | <0.001 |

*Note*: CI = confidence interval; SD = standard deviation. Models were adjusted for chronological age and sex. See Panel 1 for model abbreviations.

**Table S8**. Association between the frailty phenotype and MileAge delta (adj.)

| Model | Level | OR | 95% CI | | *p* |
|---|---|---|---|---|---|
| SVM radial | ≤ X̄-1SD | | Reference | | |
| | Middle | 1.01 | 0.97 | 1.05 | 0.608 |
| | ≥ X̄+1SD | 1.20 | 1.14 | 1.25 | <0.001 |
| SVM polynomial | ≤ X̄-1SD | | Reference | | |
| | Middle | 1.03 | 0.99 | 1.07 | 0.122 |
| | ≥ X̄+1SD | 1.25 | 1.19 | 1.31 | <0.001 |
| Cubist rules | ≤ X̄-1SD | | Reference | | |
| | Middle | 1.05 | 1.01 | 1.09 | 0.008 |
| | ≥ X̄+1SD | 1.29 | 1.23 | 1.35 | <0.001 |
| RuleFit ensemble | ≤ X̄-1SD | | Reference | | |
| | Middle | 1.03 | 1.00 | 1.07 | 0.055 |
| | ≥ X̄+1SD | 1.24 | 1.18 | 1.29 | <0.001 |
| XGBoost | ≤ X̄-1SD | | Reference | | |
| | Middle | 1.06 | 1.02 | 1.09 | 0.002 |
| | ≥ X̄+1SD | 1.25 | 1.19 | 1.31 | <0.001 |
| BART | ≤ X̄-1SD | | Reference | | |
| | Middle | 1.06 | 1.02 | 1.09 | 0.003 |
| | ≥ X̄+1SD | 1.28 | 1.23 | 1.34 | <0.001 |
| SVM linear | ≤ X̄-1SD | | Reference | | |
| | Middle | 1.06 | 1.02 | 1.11 | 0.003 |
| | ≥ X̄+1SD | 1.29 | 1.23 | 1.36 | <0.001 |
| Elastic net | ≤ X̄-1SD | | Reference | | |
| | Middle | 1.04 | 1.00 | 1.08 | 0.037 |
| | ≥ X̄+1SD | 1.26 | 1.21 | 1.32 | <0.001 |
| LASSO | ≤ X̄-1SD | | Reference | | |
| | Middle | 1.04 | 1.00 | 1.08 | 0.036 |
| | ≥ X̄+1SD | 1.27 | 1.21 | 1.33 | <0.001 |
| Ridge | ≤ X̄-1SD | | Reference | | |
| | Middle | 1.05 | 1.02 | 1.09 | 0.003 |
| | ≥ X̄+1SD | 1.25 | 1.20 | 1.31 | <0.001 |
| Random forest | ≤ X̄-1SD | | Reference | | |
| | Middle | 1.10 | 1.07 | 1.14 | <0.001 |
| | ≥ X̄+1SD | 1.20 | 1.15 | 1.25 | <0.001 |
| Bagging | ≤ X̄-1SD | | Reference | | |
| | Middle | 1.10 | 1.06 | 1.14 | <0.001 |
| | ≥ X̄+1SD | 1.21 | 1.15 | 1.26 | <0.001 |
| PLSR | ≤ X̄-1SD | | Reference | | |
| | Middle | 1.09 | 1.05 | 1.13 | <0.001 |
| | ≥ X̄+1SD | 1.27 | 1.21 | 1.33 | <0.001 |
| Regression tree | ≤ X̄-1SD | | Reference | | |
| | Middle | 1.04 | 1.01 | 1.08 | 0.014 |
| | ≥ X̄+1SD | 1.20 | 1.15 | 1.25 | <0.001 |
| KNN | ≤ X̄-1SD | | Reference | | |
| | Middle | 1.04 | 1.00 | 1.08 | 0.031 |
| | ≥ X̄+1SD | 1.17 | 1.12 | 1.22 | <0.001 |
| MARS | ≤ X̄-1SD | | Reference | | |

| | | | | | |
|---|---|---|---|---|---|
| | Middle | 0.99 | 0.96 | 1.03 | 0.667 |
| | ≥ $\bar{X}$+1SD | 1.12 | 1.07 | 1.17 | <0.001 |
| MARS ensemble | ≤ $\bar{X}$-1SD | | Reference | | |
| | Middle | 0.97 | 0.94 | 1.01 | 0.122 |
| | ≥ $\bar{X}$+1SD | 1.14 | 1.08 | 1.19 | <0.001 |

*Note*: OR = odds ratio; CI = confidence interval; SD = standard deviation. Proportional odds ratios indicate the changes in odds of being frailer associated with the MileAge delta (adj.) level relative to the reference group. Models were adjusted for chronological age and sex. See Panel 1 for model abbreviations.

**Table S9**. Association between telomere length and MileAge delta (adj.)

| Model | Level | *β* | 95% CI | | *p* |
|---|---|---|---|---|---|
| SVM radial | ≤ $\bar{X}$-1SD | | Reference | | |
| | Middle | 0.031 | 0.013 | 0.048 | <0.001 |
| | ≥ $\bar{X}$+1SD | 0.055 | 0.034 | 0.077 | <0.001 |
| SVM polynomial | ≤ $\bar{X}$-1SD | | Reference | | |
| | Middle | 0.031 | 0.013 | 0.048 | <0.001 |
| | ≥ $\bar{X}$+1SD | 0.057 | 0.035 | 0.079 | <0.001 |
| Cubist rules | ≤ $\bar{X}$-1SD | | Reference | | |
| | Middle | 0.015 | -0.002 | 0.032 | 0.088 |
| | ≥ $\bar{X}$+1SD | 0.052 | 0.030 | 0.073 | <0.001 |
| RuleFit ensemble | ≤ $\bar{X}$-1SD | | Reference | | |
| | Middle | 0.021 | 0.004 | 0.038 | 0.013 |
| | ≥ $\bar{X}$+1SD | 0.052 | 0.031 | 0.074 | <0.001 |
| XGBoost | ≤ $\bar{X}$-1SD | | Reference | | |
| | Middle | 0.024 | 0.007 | 0.041 | 0.006 |
| | ≥ $\bar{X}$+1SD | 0.050 | 0.029 | 0.072 | <0.001 |
| BART | ≤ $\bar{X}$-1SD | | Reference | | |
| | Middle | 0.021 | 0.004 | 0.038 | 0.015 |
| | ≥ $\bar{X}$+1SD | 0.052 | 0.031 | 0.074 | <0.001 |
| SVM linear | ≤ $\bar{X}$-1SD | | Reference | | |
| | Middle | 0.014 | -0.005 | 0.033 | 0.162 |
| | ≥ $\bar{X}$+1SD | 0.035 | 0.011 | 0.060 | 0.005 |
| Elastic net | ≤ $\bar{X}$-1SD | | Reference | | |
| | Middle | 0.022 | 0.005 | 0.040 | 0.011 |
| | ≥ $\bar{X}$+1SD | 0.052 | 0.030 | 0.074 | <0.001 |
| LASSO | ≤ $\bar{X}$-1SD | | Reference | | |
| | Middle | 0.022 | 0.005 | 0.039 | 0.013 |
| | ≥ $\bar{X}$+1SD | 0.052 | 0.030 | 0.074 | <0.001 |
| Ridge | ≤ $\bar{X}$-1SD | | Reference | | |
| | Middle | 0.033 | 0.016 | 0.050 | <0.001 |
| | ≥ $\bar{X}$+1SD | 0.052 | 0.030 | 0.074 | <0.001 |
| Random forest | ≤ $\bar{X}$-1SD | | Reference | | |
| | Middle | 0.009 | -0.008 | 0.026 | 0.292 |
| | ≥ $\bar{X}$+1SD | 0.020 | -0.001 | 0.042 | 0.062 |
| Bagging | ≤ $\bar{X}$-1SD | | Reference | | |
| | Middle | 0.008 | -0.009 | 0.025 | 0.357 |
| | ≥ $\bar{X}$+1SD | 0.020 | -0.002 | 0.041 | 0.071 |
| PLSR | ≤ $\bar{X}$-1SD | | Reference | | |
| | Middle | 0.029 | 0.011 | 0.046 | 0.001 |
| | ≥ $\bar{X}$+1SD | 0.036 | 0.013 | 0.058 | 0.002 |
| Regression tree | ≤ $\bar{X}$-1SD | | Reference | | |
| | Middle | 0.001 | -0.015 | 0.018 | 0.872 |
| | ≥ $\bar{X}$+1SD | 0.014 | -0.007 | 0.036 | 0.177 |
| KNN | ≤ $\bar{X}$-1SD | | Reference | | |
| | Middle | 0.005 | -0.012 | 0.022 | 0.563 |
| | ≥ $\bar{X}$+1SD | 0.030 | 0.009 | 0.051 | 0.006 |
| MARS | ≤ $\bar{X}$-1SD | | Reference | | |

| | | | | | |
|---|---|---|---|---|---|
| | Middle | -0.018 | -0.035 | -0.001 | 0.041 |
| | ≥ X̄+1SD | -0.010 | -0.032 | 0.012 | 0.369 |
| MARS ensemble | ≤ X̄-1SD | | Reference | | |
| | Middle | -0.015 | -0.033 | 0.003 | 0.095 |
| | ≥ X̄+1SD | -0.020 | -0.043 | 0.003 | 0.084 |

*Note*: CI = confidence interval; SD = standard deviation. Models were adjusted for chronological age and sex. See Panel 1 for model abbreviations.

**Table S10**. Association between having a long-standing illness and MileAge delta (adj.)

| Model | Level | OR | 95% CI | | *p* |
|---|---|---|---|---|---|
| SVM radial | ≤ X̄-1SD | | Reference | | |
| | Middle | 1.14 | 1.10 | 1.19 | <0.001 |
| | ≥ X̄+1SD | 1.71 | 1.63 | 1.80 | <0.001 |
| SVM polynomial | ≤ X̄-1SD | | Reference | | |
| | Middle | 1.13 | 1.09 | 1.18 | <0.001 |
| | ≥ X̄+1SD | 1.75 | 1.66 | 1.83 | <0.001 |
| Cubist rules | ≤ X̄-1SD | | Reference | | |
| | Middle | 1.17 | 1.12 | 1.21 | <0.001 |
| | ≥ X̄+1SD | 1.82 | 1.73 | 1.91 | <0.001 |
| RuleFit ensemble | ≤ X̄-1SD | | Reference | | |
| | Middle | 1.10 | 1.06 | 1.15 | <0.001 |
| | ≥ X̄+1SD | 1.65 | 1.57 | 1.73 | <0.001 |
| XGBoost | ≤ X̄-1SD | | Reference | | |
| | Middle | 1.17 | 1.13 | 1.22 | <0.001 |
| | ≥ X̄+1SD | 1.75 | 1.67 | 1.84 | <0.001 |
| BART | ≤ X̄-1SD | | Reference | | |
| | Middle | 1.17 | 1.13 | 1.22 | <0.001 |
| | ≥ X̄+1SD | 1.69 | 1.61 | 1.78 | <0.001 |
| SVM linear | ≤ X̄-1SD | | Reference | | |
| | Middle | 1.12 | 1.07 | 1.17 | <0.001 |
| | ≥ X̄+1SD | 1.62 | 1.54 | 1.71 | <0.001 |
| Elastic net | ≤ X̄-1SD | | Reference | | |
| | Middle | 1.11 | 1.07 | 1.16 | <0.001 |
| | ≥ X̄+1SD | 1.61 | 1.53 | 1.69 | <0.001 |
| LASSO | ≤ X̄-1SD | | Reference | | |
| | Middle | 1.11 | 1.07 | 1.16 | <0.001 |
| | ≥ X̄+1SD | 1.61 | 1.54 | 1.69 | <0.001 |
| Ridge | ≤ X̄-1SD | | Reference | | |
| | Middle | 1.13 | 1.09 | 1.18 | <0.001 |
| | ≥ X̄+1SD | 1.61 | 1.53 | 1.69 | <0.001 |
| Random forest | ≤ X̄-1SD | | Reference | | |
| | Middle | 1.25 | 1.21 | 1.30 | <0.001 |
| | ≥ X̄+1SD | 1.66 | 1.58 | 1.74 | <0.001 |
| Bagging | ≤ X̄-1SD | | Reference | | |
| | Middle | 1.22 | 1.17 | 1.27 | <0.001 |
| | ≥ X̄+1SD | 1.67 | 1.59 | 1.75 | <0.001 |
| PLSR | ≤ X̄-1SD | | Reference | | |
| | Middle | 1.18 | 1.13 | 1.23 | <0.001 |
| | ≥ X̄+1SD | 1.64 | 1.56 | 1.72 | <0.001 |
| Regression tree | ≤ X̄-1SD | | Reference | | |
| | Middle | 1.09 | 1.05 | 1.14 | <0.001 |
| | ≥ X̄+1SD | 1.52 | 1.45 | 1.59 | <0.001 |
| KNN | ≤ X̄-1SD | | Reference | | |
| | Middle | 1.19 | 1.14 | 1.24 | <0.001 |
| | ≥ X̄+1SD | 1.63 | 1.55 | 1.71 | <0.001 |
| MARS | ≤ X̄-1SD | | Reference | | |

| | | | | | |
|---|---|---|---|---|---|
| | Middle | 1.00 | 0.96 | 1.04 | 0.914 |
| | ≥ $\bar{X}$+1SD | 1.23 | 1.17 | 1.29 | <0.001 |
| MARS ensemble | ≤ $\bar{X}$-1SD | | Reference | | |
| | Middle | 1.05 | 1.01 | 1.10 | 0.013 |
| | ≥ $\bar{X}$+1SD | 1.43 | 1.36 | 1.51 | <0.001 |

*Note*: OR = odds ratio; CI = confidence interval; SD = standard deviation. Models were adjusted for chronological age and sex. See Panel 1 for model abbreviations.

**Table S11**. Association between health status and MileAge delta (adj.)

| Model | Level | OR | 95% CI | | *p* |
|---|---|---|---|---|---|
| SVM radial | ≤ X̄-1SD | | Reference | | |
| | Middle | 1.17 | 1.12 | 1.22 | <0.001 |
| | ≥ X̄+1SD | 1.79 | 1.70 | 1.88 | <0.001 |
| SVM polynomial | ≤ X̄-1SD | | Reference | | |
| | Middle | 1.14 | 1.10 | 1.19 | <0.001 |
| | ≥ X̄+1SD | 1.79 | 1.71 | 1.88 | <0.001 |
| Cubist rules | ≤ X̄-1SD | | Reference | | |
| | Middle | 1.18 | 1.14 | 1.23 | <0.001 |
| | ≥ X̄+1SD | 1.85 | 1.76 | 1.94 | <0.001 |
| RuleFit ensemble | ≤ X̄-1SD | | Reference | | |
| | Middle | 1.10 | 1.05 | 1.14 | <0.001 |
| | ≥ X̄+1SD | 1.70 | 1.62 | 1.79 | <0.001 |
| XGBoost | ≤ X̄-1SD | | Reference | | |
| | Middle | 1.14 | 1.10 | 1.19 | <0.001 |
| | ≥ X̄+1SD | 1.77 | 1.68 | 1.85 | <0.001 |
| BART | ≤ X̄-1SD | | Reference | | |
| | Middle | 1.17 | 1.12 | 1.22 | <0.001 |
| | ≥ X̄+1SD | 1.75 | 1.67 | 1.84 | <0.001 |
| SVM linear | ≤ X̄-1SD | | Reference | | |
| | Middle | 1.11 | 1.06 | 1.16 | <0.001 |
| | ≥ X̄+1SD | 1.59 | 1.51 | 1.68 | <0.001 |
| Elastic net | ≤ X̄-1SD | | Reference | | |
| | Middle | 1.13 | 1.09 | 1.18 | <0.001 |
| | ≥ X̄+1SD | 1.60 | 1.52 | 1.68 | <0.001 |
| LASSO | ≤ X̄-1SD | | Reference | | |
| | Middle | 1.13 | 1.08 | 1.17 | <0.001 |
| | ≥ X̄+1SD | 1.60 | 1.53 | 1.68 | <0.001 |
| Ridge | ≤ X̄-1SD | | Reference | | |
| | Middle | 1.10 | 1.06 | 1.15 | <0.001 |
| | ≥ X̄+1SD | 1.57 | 1.50 | 1.65 | <0.001 |
| Random forest | ≤ X̄-1SD | | Reference | | |
| | Middle | 1.21 | 1.16 | 1.25 | <0.001 |
| | ≥ X̄+1SD | 1.66 | 1.58 | 1.74 | <0.001 |
| Bagging | ≤ X̄-1SD | | Reference | | |
| | Middle | 1.17 | 1.12 | 1.21 | <0.001 |
| | ≥ X̄+1SD | 1.66 | 1.58 | 1.74 | <0.001 |
| PLSR | ≤ X̄-1SD | | Reference | | |
| | Middle | 1.19 | 1.15 | 1.24 | <0.001 |
| | ≥ X̄+1SD | 1.64 | 1.56 | 1.73 | <0.001 |
| Regression tree | ≤ X̄-1SD | | Reference | | |
| | Middle | 1.06 | 1.02 | 1.10 | 0.002 |
| | ≥ X̄+1SD | 1.53 | 1.46 | 1.60 | <0.001 |
| KNN | ≤ X̄-1SD | | Reference | | |
| | Middle | 1.15 | 1.11 | 1.19 | <0.001 |
| | ≥ X̄+1SD | 1.66 | 1.59 | 1.74 | <0.001 |
| MARS | ≤ X̄-1SD | | Reference | | |

| | | | | | |
|---|---|---|---|---|---|
| | Middle | 0.97 | 0.93 | 1.00 | 0.073 |
| | ≥ X̄+1SD | 1.11 | 1.05 | 1.16 | <0.001 |
| MARS ensemble | ≤ X̄-1SD | | Reference | | |
| | Middle | 1.04 | 1.00 | 1.08 | 0.052 |
| | ≥ X̄+1SD | 1.40 | 1.33 | 1.47 | <0.001 |

*Note*: OR = odds ratio; CI = confidence interval; SD = standard deviation. Models were adjusted for chronological age and sex. See Panel 1 for model abbreviations.

**Table S12**. Association between self-rated health and MileAge delta (adj.)

| Model | Level | OR | 95% CI | | *p* |
|---|---|---|---|---|---|
| SVM radial | ≤ $\bar{X}$-1SD | | | Reference | |
| | Middle | 1.16 | 1.12 | 1.20 | <0.001 |
| | ≥ $\bar{X}$+1SD | 1.62 | 1.55 | 1.69 | <0.001 |
| SVM polynomial | ≤ $\bar{X}$-1SD | | | Reference | |
| | Middle | 1.18 | | Middle | 1.18 |
| | ≥ $\bar{X}$+1SD | 1.67 | | ≥ $\bar{X}$+1SD | 1.67 |
| Cubist rules | ≤ $\bar{X}$-1SD | | | Reference | |
| | Middle | 1.18 | 1.14 | 1.22 | <0.001 |
| | ≥ $\bar{X}$+1SD | 1.72 | 1.65 | 1.80 | <0.001 |
| RuleFit ensemble | ≤ $\bar{X}$-1SD | | | Reference | |
| | Middle | 1.15 | 1.11 | 1.19 | <0.001 |
| | ≥ $\bar{X}$+1SD | 1.59 | 1.52 | 1.66 | <0.001 |
| XGBoost | ≤ $\bar{X}$-1SD | | | Reference | |
| | Middle | 1.22 | 1.18 | 1.26 | <0.001 |
| | ≥ $\bar{X}$+1SD | 1.66 | 1.59 | 1.73 | <0.001 |
| BART | ≤ $\bar{X}$-1SD | | | Reference | |
| | Middle | 1.21 | 1.17 | 1.25 | <0.001 |
| | ≥ $\bar{X}$+1SD | 1.65 | 1.58 | 1.72 | <0.001 |
| SVM linear | ≤ $\bar{X}$-1SD | | | Reference | |
| | Middle | 1.19 | 1.14 | 1.23 | <0.001 |
| | ≥ $\bar{X}$+1SD | 1.69 | 1.61 | 1.78 | <0.001 |
| Elastic net | ≤ $\bar{X}$-1SD | | | Reference | |
| | Middle | 1.15 | 1.11 | 1.19 | <0.001 |
| | ≥ $\bar{X}$+1SD | 1.61 | 1.54 | 1.68 | <0.001 |
| LASSO | ≤ $\bar{X}$-1SD | | | Reference | |
| | Middle | 1.14 | 1.11 | 1.18 | <0.001 |
| | ≥ $\bar{X}$+1SD | 1.61 | 1.54 | 1.68 | <0.001 |
| Ridge | ≤ $\bar{X}$-1SD | | | Reference | |
| | Middle | 1.18 | 1.14 | 1.23 | <0.001 |
| | ≥ $\bar{X}$+1SD | 1.64 | 1.57 | 1.72 | <0.001 |
| Random forest | ≤ $\bar{X}$-1SD | | | Reference | |
| | Middle | 1.32 | 1.28 | 1.36 | <0.001 |
| | ≥ $\bar{X}$+1SD | 1.61 | 1.54 | 1.68 | <0.001 |
| Bagging | ≤ $\bar{X}$-1SD | | | Reference | |
| | Middle | 1.30 | 1.26 | 1.35 | <0.001 |
| | ≥ $\bar{X}$+1SD | 1.66 | 1.59 | 1.73 | <0.001 |
| PLSR | ≤ $\bar{X}$-1SD | | | Reference | |
| | Middle | 1.25 | 1.21 | 1.30 | <0.001 |
| | ≥ $\bar{X}$+1SD | 1.71 | 1.63 | 1.78 | <0.001 |
| Regression tree | ≤ $\bar{X}$-1SD | | | Reference | |
| | Middle | 1.19 | 1.15 | 1.23 | <0.001 |
| | ≥ $\bar{X}$+1SD | 1.53 | 1.47 | 1.60 | <0.001 |
| KNN | ≤ $\bar{X}$-1SD | | | Reference | |
| | Middle | 1.22 | 1.18 | 1.26 | <0.001 |
| | ≥ $\bar{X}$+1SD | 1.60 | 1.53 | 1.67 | <0.001 |
| MARS | ≤ $\bar{X}$-1SD | | | Reference | |

| | | | | | |
|---|---|---|---|---|---|
| | Middle | 1.08 | 1.05 | 1.12 | <0.001 |
| | ≥ X̄+1SD | 1.34 | 1.29 | 1.40 | <0.001 |
| MARS ensemble | ≤ X̄-1SD | | Reference | | |
| | Middle | 1.05 | 1.01 | 1.08 | 0.014 |
| | ≥ X̄+1SD | 1.35 | 1.29 | 1.41 | <0.001 |

*Note*: OR = odds ratio; CI = confidence interval; SD = standard deviation. Proportional odds ratios indicate the changes in odds of reporting worse self-rated health associated with the MileAge delta (adj.) level relative to the reference group. Models were adjusted for chronological age and sex. See Panel 1 for model abbreviations.

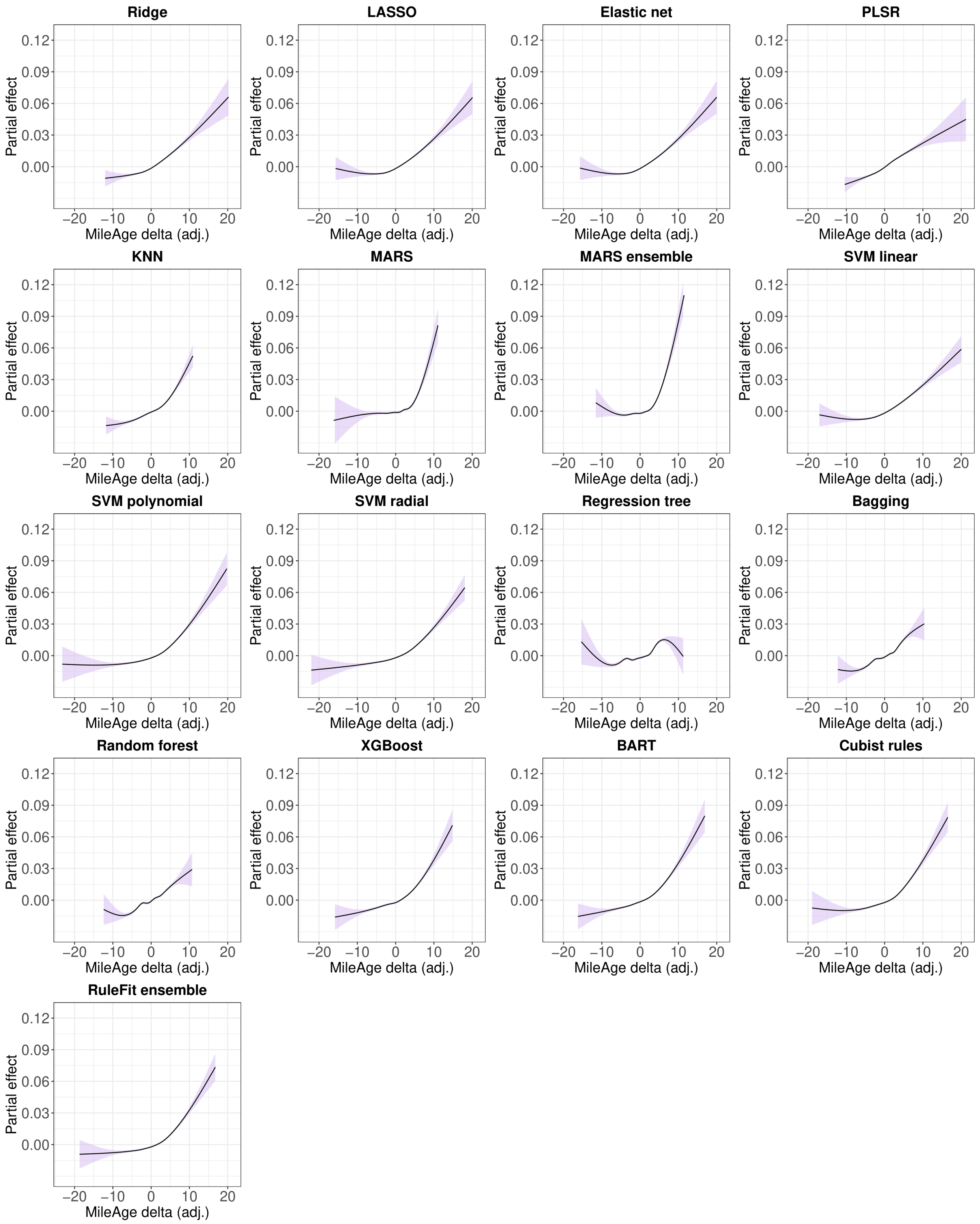

**Figure S50**. Partial effect plots of generalised additive models of the association between the frailty index and MileAge delta (adj.) for all models. Models were adjusted for chronological age and sex. The shaded areas correspond to 95% confidence intervals.

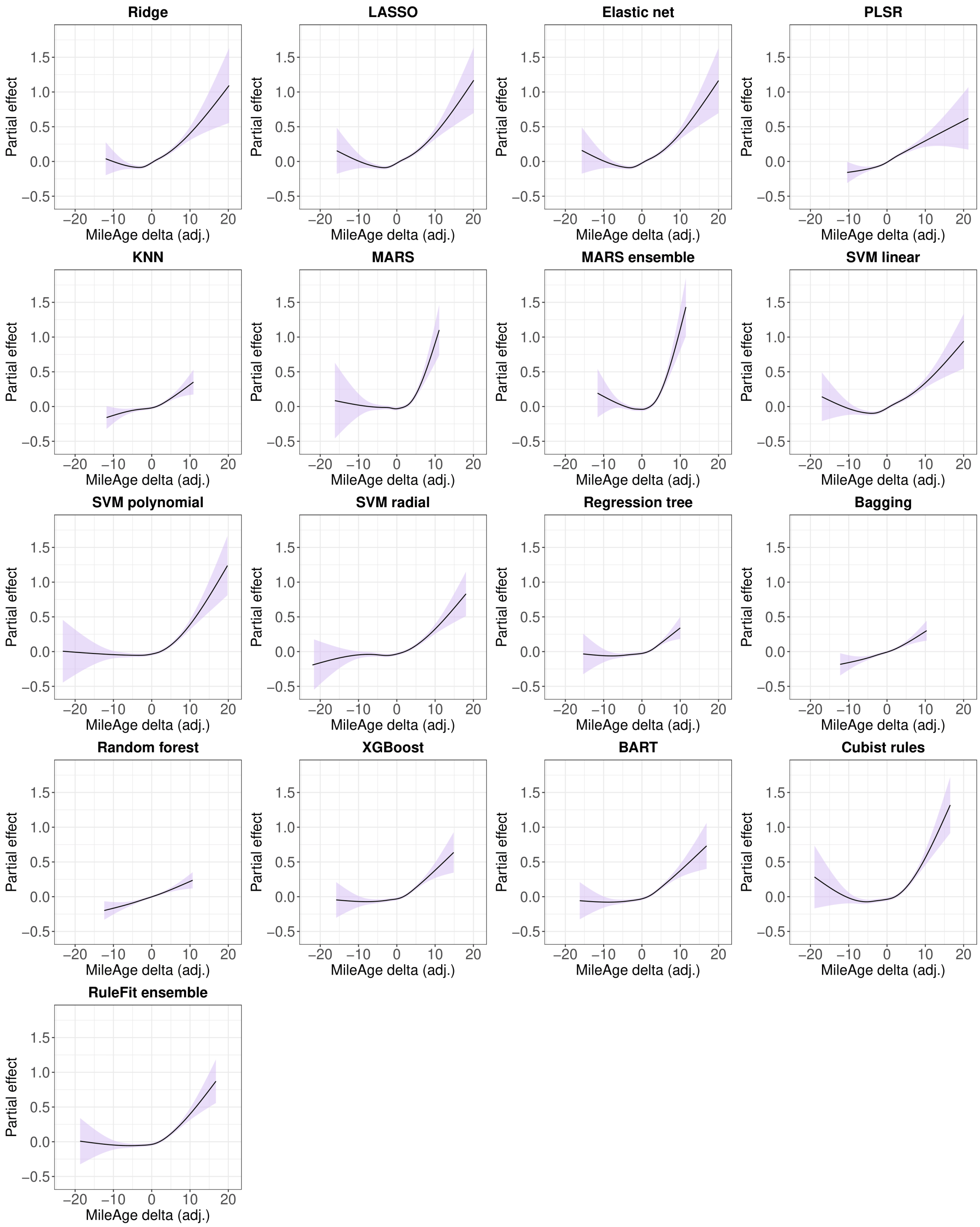

**Figure S51**. Partial effect plots of generalised additive models of the association between the frailty phenotype and MileAge delta (adj.) for all models. Models were adjusted for chronological age and sex. The shaded areas correspond to 95% confidence intervals.

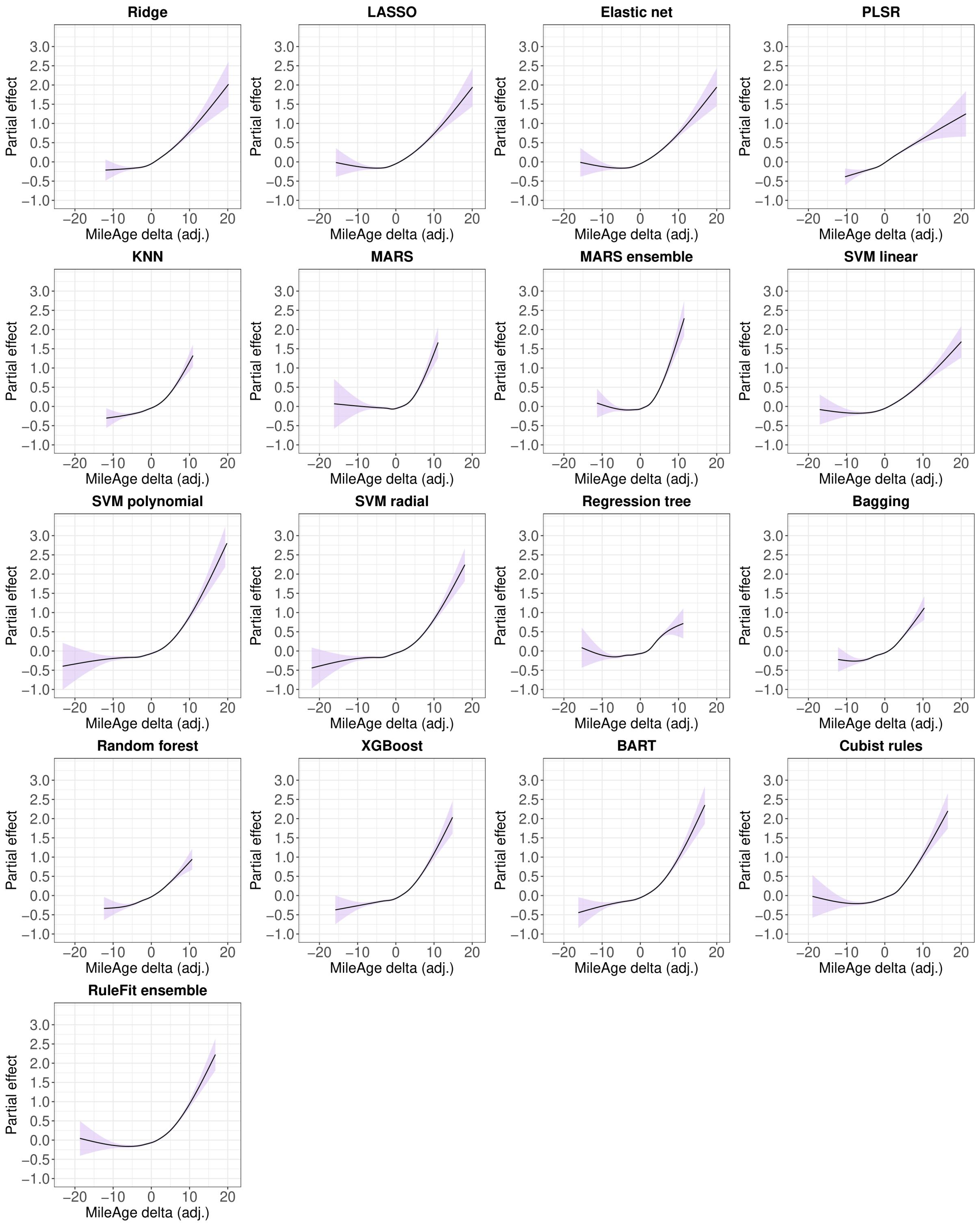

**Figure S52**. Partial effect plots of generalised additive models of the association between having a long-standing illness and MileAge delta (adj.) for all models. Models were adjusted for chronological age and sex. The shaded areas correspond to 95% confidence intervals.

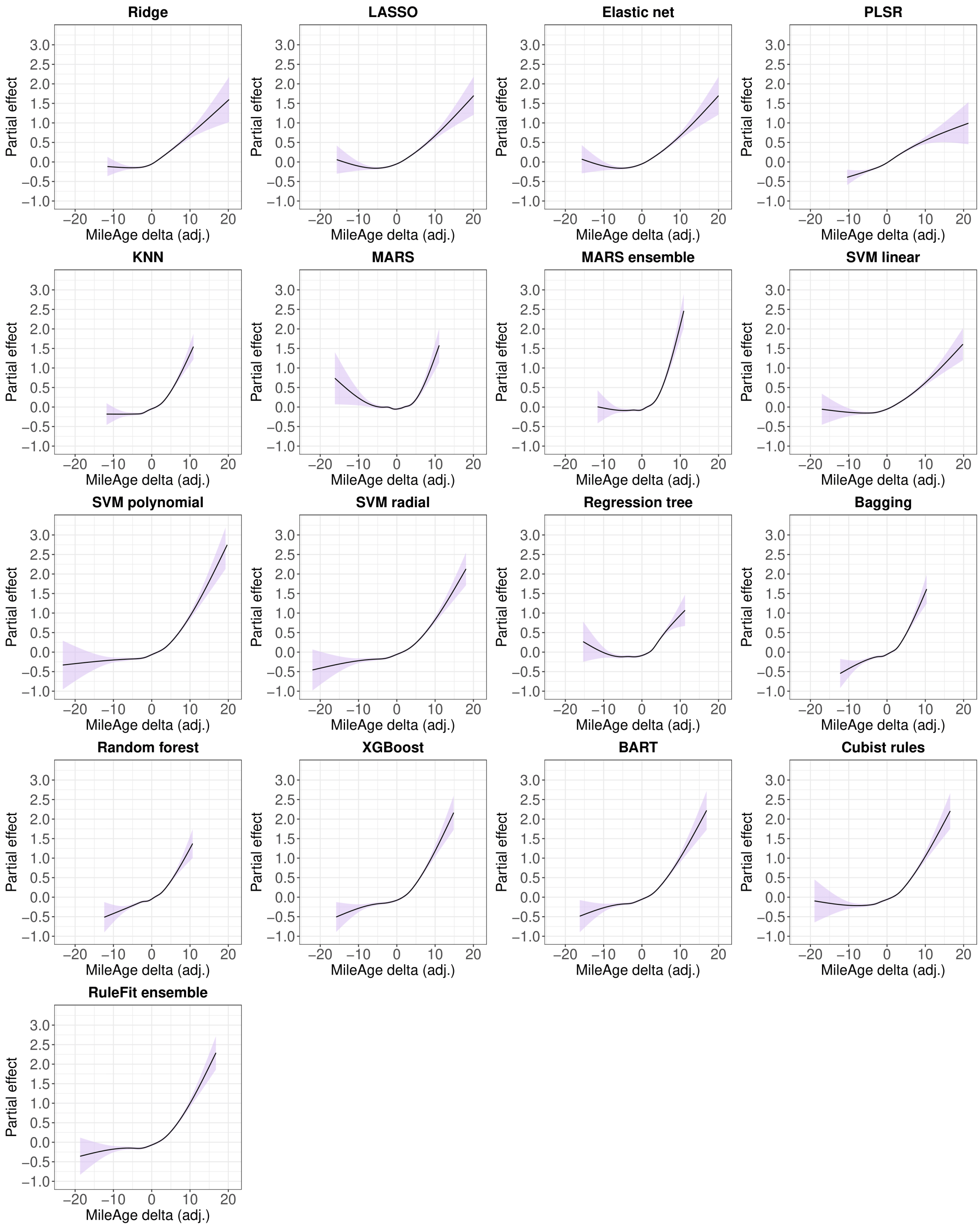

**Figure S53**. Partial effect plots of generalised additive models of the association between health status and MileAge delta (adj.) for all models. Models were adjusted for chronological age and sex. The shaded areas correspond to 95% confidence intervals.

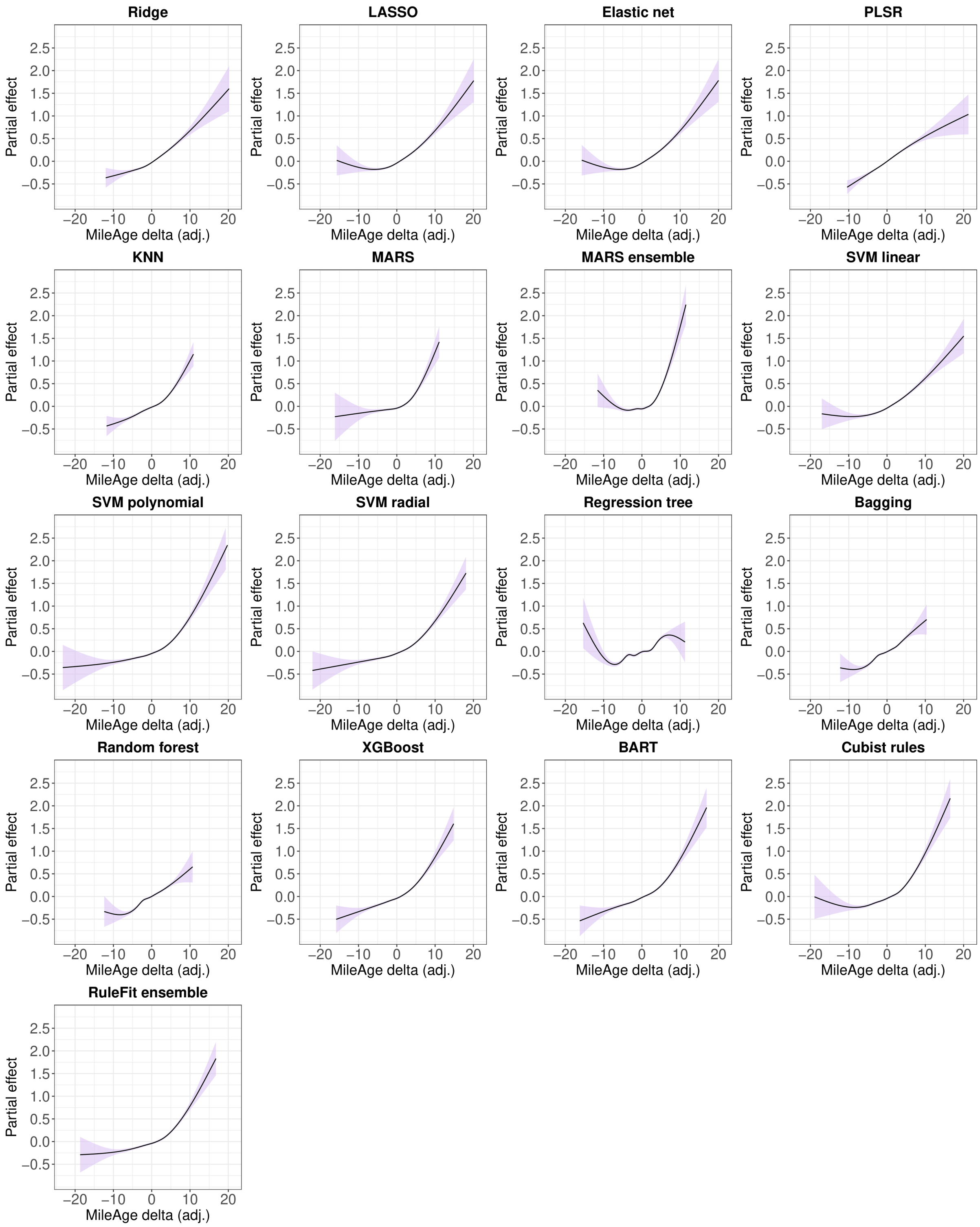

**Figure S54**. Partial effect plots of generalised additive models of the association between self-rated health and MileAge delta (adj.) for all models. Models were adjusted for chronological age and sex. The shaded areas correspond to 95% confidence intervals.

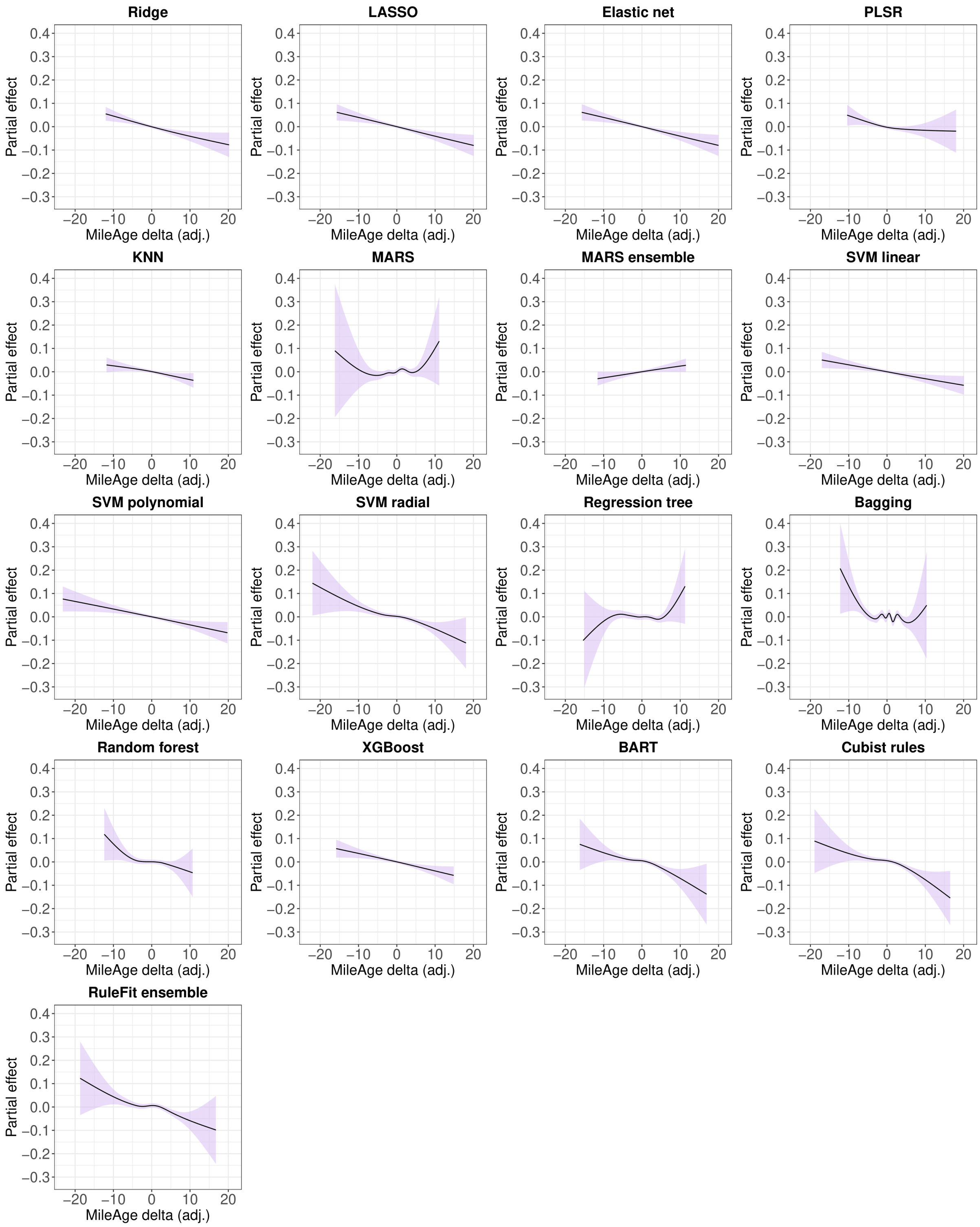

**Figure S55**. Partial effect plots of generalised additive models of the association between telomere length and MileAge delta (adj.) for all models. Models were adjusted for chronological age and sex. The shaded areas correspond to 95% confidence intervals.

#### 14. Age adjustment and time axis in the prospective analysis

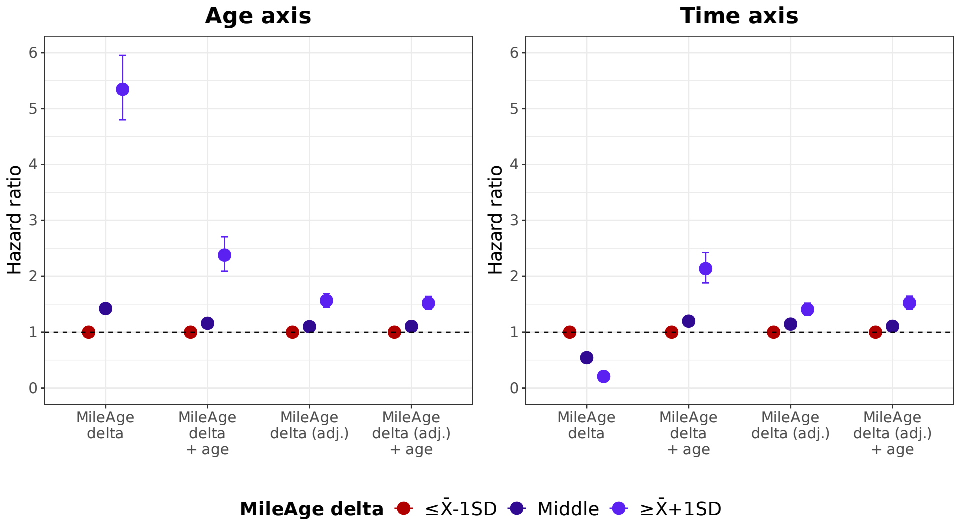

**Figure S56**. Hazard ratios (HR) and 95% confidence intervals from Cox proportional hazards models for all-cause mortality for unadjusted and age bias adjusted MileAge and MileAge delta with and without adjustment for chronological age as covariate, using age (in years) and time (in days) since the baseline assessment as the underlying time axis. All analyses were adjusted for sex. SD = standard deviation.

**Table S13**. All-cause mortality by age adjustment and time axis

| Predictor | Level | $N_{total}$ | $N_{died}$ | HR | 95% CI | | $p$ |
|---|---|---|---|---|---|---|---|
| Age (in years) | | | | | | | |
| MileAge delta | ≤ X̄-1SD | 16523 | 2392 | Reference | | | |
| | Middle | 67196 | 5252 | 1.42 | 1.35 | 1.50 | <0.001 |
| | ≥ X̄+1SD | 17640 | 554 | 5.35 | 4.80 | 5.95 | <0.001 |
| MileAge delta + age | ≤ X̄-1SD | 16523 | 2392 | Reference | | | |
| | Middle | 67196 | 5252 | 1.16 | 1.10 | 1.22 | <0.001 |
| | ≥ X̄+1SD | 17640 | 554 | 2.38 | 2.09 | 2.71 | <0.001 |
| MileAge delta (adj.) | ≤ X̄-1SD | 16204 | 1160 | Reference | | | |
| | Middle | 69166 | 5525 | 1.10 | 1.03 | 1.17 | 0.004 |
| | ≥ X̄+1SD | 15989 | 1513 | 1.57 | 1.45 | 1.69 | <0.001 |
| MileAge delta (adj.) + age | ≤ X̄-1SD | 16204 | 1160 | Reference | | | |
| | Middle | 69166 | 5525 | 1.10 | 1.04 | 1.18 | 0.003 |
| | ≥ X̄+1SD | 15989 | 1513 | 1.52 | 1.41 | 1.64 | <0.001 |
| Time since baseline assessment | | | | | | | |
| MileAge delta | ≤ X̄-1SD | 16523 | 2392 | Reference | | | |
| | Middle | 67196 | 5252 | 0.54 | 0.52 | 0.57 | <0.001 |
| | ≥ X̄+1SD | 17640 | 554 | 0.21 | 0.19 | 0.23 | <0.001 |
| MileAge delta + age | ≤ X̄-1SD | 16523 | 2392 | Reference | | | |
| | Middle | 67196 | 5252 | 1.20 | 1.13 | 1.26 | <0.001 |
| | ≥ X̄+1SD | 17640 | 554 | 2.14 | 1.88 | 2.43 | <0.001 |
| MileAge delta (adj.) | ≤ X̄-1SD | 16204 | 1160 | Reference | | | |
| | Middle | 69166 | 5525 | 1.14 | 1.07 | 1.22 | <0.001 |
| | ≥ X̄+1SD | 15989 | 1513 | 1.41 | 1.30 | 1.52 | <0.001 |
| MileAge delta (adj.) + age | ≤ X̄-1SD | 16204 | 1160 | Reference | | | |
| | Middle | 69166 | 5525 | 1.11 | 1.04 | 1.18 | 0.002 |
| | ≥ X̄+1SD | 15989 | 1513 | 1.52 | 1.41 | 1.64 | <0.001 |

*Note:* HR = hazard ratio; CI = confidence interval; SD = standard deviation. MileAge delta derived from Cubist rule-based regression model. All models were adjusted for sex.

#### 15. All-cause mortality by MileAge delta

**Table S14**. All-cause mortality by MileAge delta (adj.)

| Model | Level | $N_{total}$ | $N_{died}$ | HR | 95% CI | | *p* |
|---|---|---|---|---|---|---|---|
| SVM radial | ≤ X̄-1SD | 15973 | 1161 | | Reference | | |
| | Middle | 69614 | 5565 | 1.07 | 1.01 | 1.15 | 0.026 |
| | ≥ X̄+1SD | 15772 | 1472 | 1.43 | 1.32 | 1.54 | <0.001 |
| SVM polynomial | ≤ X̄-1SD | 15762 | 1143 | | Reference | | |
| | Middle | 69891 | 5539 | 1.08 | 1.01 | 1.15 | 0.017 |
| | ≥ X̄+1SD | 15706 | 1516 | 1.50 | 1.38 | 1.62 | <0.001 |
| Cubist rules | ≤ X̄-1SD | 16204 | 1160 | | Reference | | |
| | Middle | 69166 | 5525 | 1.10 | 1.04 | 1.18 | 0.003 |
| | ≥ X̄+1SD | 15989 | 1513 | 1.52 | 1.41 | 1.64 | <0.001 |
| RuleFit ensemble | ≤ X̄-1SD | 16116 | 1161 | | Reference | | |
| | Middle | 69394 | 5549 | 1.13 | 1.06 | 1.20 | <0.001 |
| | ≥ X̄+1SD | 15849 | 1488 | 1.45 | 1.34 | 1.57 | <0.001 |
| XGBoost | ≤ X̄-1SD | 16496 | 1178 | | Reference | | |
| | Middle | 68827 | 5550 | 1.10 | 1.03 | 1.17 | 0.005 |
| | ≥ X̄+1SD | 16036 | 1470 | 1.44 | 1.33 | 1.56 | <0.001 |
| BART | ≤ X̄-1SD | 16174 | 1187 | | Reference | | |
| | Middle | 69287 | 5492 | 1.09 | 1.03 | 1.17 | 0.005 |
| | ≥ X̄+1SD | 15898 | 1519 | 1.42 | 1.32 | 1.54 | <0.001 |
| SVM linear | ≤ X̄-1SD | 12579 | 949 | | Reference | | |
| | Middle | 56082 | 4308 | 1.08 | 1.01 | 1.16 | 0.030 |
| | ≥ X̄+1SD | 12431 | 1269 | 1.50 | 1.38 | 1.63 | <0.001 |
| Elastic net | ≤ X̄-1SD | 15614 | 1197 | | Reference | | |
| | Middle | 70068 | 5432 | 1.08 | 1.02 | 1.15 | 0.012 |
| | ≥ X̄+1SD | 15677 | 1569 | 1.46 | 1.36 | 1.58 | <0.001 |
| LASSO | ≤ X̄-1SD | 15593 | 1199 | | Reference | | |
| | Middle | 70114 | 5415 | 1.08 | 1.01 | 1.15 | 0.023 |
| | ≥ X̄+1SD | 15652 | 1584 | 1.48 | 1.37 | 1.59 | <0.001 |
| Ridge | ≤ X̄-1SD | 15554 | 1147 | | Reference | | |
| | Middle | 70334 | 5478 | 1.14 | 1.07 | 1.22 | <0.001 |
| | ≥ X̄+1SD | 15471 | 1573 | 1.54 | 1.42 | 1.66 | <0.001 |
| Random forest | ≤ X̄-1SD | 16785 | 1246 | | Reference | | |
| | Middle | 68089 | 5661 | 1.14 | 1.07 | 1.21 | <0.001 |
| | ≥ X̄+1SD | 16485 | 1291 | 1.24 | 1.15 | 1.34 | <0.001 |
| Bagging | ≤ X̄-1SD | 17017 | 1237 | | Reference | | |
| | Middle | 67816 | 5688 | 1.14 | 1.07 | 1.21 | <0.001 |
| | ≥ X̄+1SD | 16526 | 1273 | 1.26 | 1.17 | 1.37 | <0.001 |
| PLSR | ≤ X̄-1SD | 15278 | 1091 | | Reference | | |
| | Middle | 70592 | 5583 | 1.25 | 1.17 | 1.33 | <0.001 |
| | ≥ X̄+1SD | 15489 | 1524 | 1.58 | 1.46 | 1.71 | <0.001 |
| Regression tree | ≤ X̄-1SD | 17885 | 1319 | | Reference | | |
| | Middle | 67060 | 5646 | 1.06 | 1.00 | 1.13 | 0.059 |
| | ≥ X̄+1SD | 16414 | 1233 | 1.24 | 1.14 | 1.34 | <0.001 |
| KNN | ≤ X̄-1SD | 16669 | 1209 | | Reference | | |
| | Middle | 68220 | 5585 | 1.12 | 1.05 | 1.19 | <0.001 |
| | ≥ X̄+1SD | 16470 | 1404 | 1.31 | 1.21 | 1.42 | <0.001 |

| | | | | | | | |
|---|---|---|---|---|---|---|---|
| MARS | ≤ X̄-1SD | 15962 | 1353 | Reference | | | |
| | Middle | 69618 | 5615 | 0.94 | 0.89 | 1.00 | 0.054 |
| | ≥ X̄+1SD | 15779 | 1230 | 1.12 | 1.03 | 1.21 | 0.006 |
| MARS ensemble | ≤ X̄-1SD | 14718 | 1301 | Reference | | | |
| | Middle | 62021 | 4894 | 0.94 | 0.88 | 1.00 | 0.050 |
| | ≥ X̄+1SD | 14486 | 1196 | 1.10 | 1.02 | 1.19 | 0.017 |

*Note:* HR = hazard ratio; CI = confidence interval; SD = standard deviation. Models were adjusted for chronological age and sex. Age (in years) was used as the underlying time axis. See Panel 1 for model abbreviations.

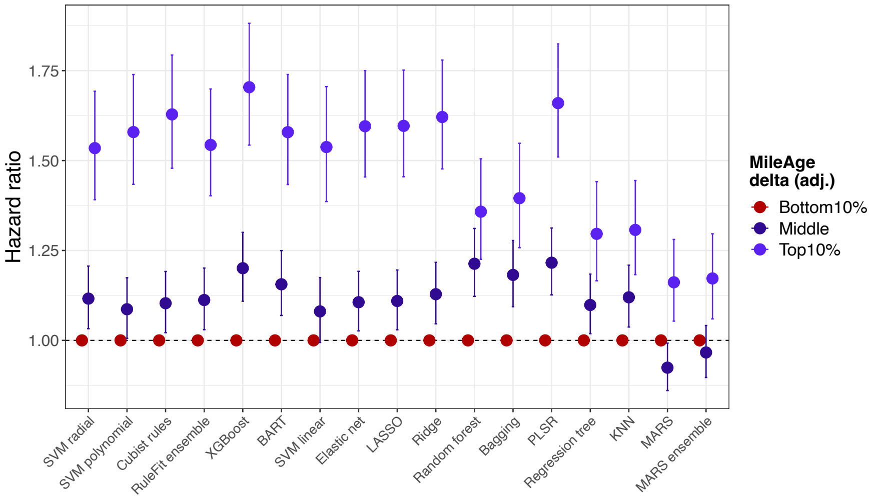

**Figure S57**. Hazard ratios (HR) and 95% confidence intervals from Cox proportional hazards models for all models. Models were adjusted for chronological age and sex. Age (in years) was used as the underlying time axis. Reference group: individuals with a MileAge delta (adj.) in the bottom 10% of the distribution. See Panel 1 for model abbreviations.

**Table S15**. All-cause mortality by MileAge delta (adj.) – distribution tails

| Model | Level | N total | N died | HR | 95% CI | | *p* |
|---|---|---|---|---|---|---|---|
| SVM radial | Bottom 10% | 10136 | 712 | | Reference | | |
| | Middle | 81087 | 6540 | 1.12 | 1.03 | 1.21 | 0.006 |
| | Top 10% | 10136 | 946 | 1.53 | 1.39 | 1.69 | <0.001 |
| SVM polynomial | Bottom 10% | 10136 | 722 | | Reference | | |
| | Middle | 81087 | 6468 | 1.09 | 1.01 | 1.17 | 0.036 |
| | Top 10% | 10136 | 1008 | 1.58 | 1.43 | 1.74 | <0.001 |
| Cubist rules | Bottom 10% | 10136 | 730 | | Reference | | |
| | Middle | 81087 | 6484 | 1.10 | 1.02 | 1.19 | 0.012 |
| | Top 10% | 10136 | 984 | 1.63 | 1.48 | 1.79 | <0.001 |
| RuleFit ensemble | Bottom 10% | 10136 | 733 | | Reference | | |
| | Middle | 81087 | 6465 | 1.11 | 1.03 | 1.20 | 0.007 |
| | Top 10% | 10136 | 1000 | 1.54 | 1.40 | 1.70 | <0.001 |
| XGBoost | Bottom 10% | 10136 | 677 | | Reference | | |
| | Middle | 81087 | 6562 | 1.20 | 1.11 | 1.30 | <0.001 |
| | Top 10% | 10136 | 959 | 1.70 | 1.54 | 1.88 | <0.001 |
| BART | Bottom 10% | 10136 | 711 | | Reference | | |
| | Middle | 81087 | 6476 | 1.16 | 1.07 | 1.25 | <0.001 |
| | Top 10% | 10136 | 1011 | 1.58 | 1.43 | 1.74 | <0.001 |
| SVM linear | Bottom 10% | 8110 | 626 | | Reference | | |
| | Middle | 64872 | 5042 | 1.08 | 0.99 | 1.17 | 0.069 |
| | Top 10% | 8110 | 858 | 1.54 | 1.39 | 1.71 | <0.001 |
| Elastic net | Bottom 10% | 10136 | 782 | | Reference | | |
| | Middle | 81087 | 6325 | 1.11 | 1.03 | 1.19 | 0.008 |
| | Top 10% | 10136 | 1091 | 1.60 | 1.45 | 1.75 | <0.001 |
| LASSO | Bottom 10% | 10136 | 781 | | Reference | | |
| | Middle | 81087 | 6330 | 1.11 | 1.03 | 1.20 | 0.006 |
| | Top 10% | 10136 | 1087 | 1.60 | 1.46 | 1.75 | <0.001 |
| Ridge | Bottom 10% | 10136 | 761 | | Reference | | |
| | Middle | 81087 | 6340 | 1.13 | 1.05 | 1.22 | 0.002 |
| | Top 10% | 10136 | 1097 | 1.62 | 1.48 | 1.78 | <0.001 |
| Random forest | Bottom 10% | 10136 | 716 | | Reference | | |
| | Middle | 81087 | 6709 | 1.21 | 1.12 | 1.31 | <0.001 |
| | Top 10% | 10136 | 773 | 1.36 | 1.23 | 1.51 | <0.001 |
| Bagging | Bottom 10% | 10136 | 714 | | Reference | | |
| | Middle | 81087 | 6740 | 1.18 | 1.09 | 1.28 | <0.001 |
| | Top 10% | 10136 | 744 | 1.40 | 1.26 | 1.55 | <0.001 |
| PLSR | Bottom 10% | 10136 | 746 | | Reference | | |
| | Middle | 81087 | 6391 | 1.22 | 1.13 | 1.31 | <0.001 |
| | Top 10% | 10136 | 1061 | 1.66 | 1.51 | 1.82 | <0.001 |
| Regression tree | Bottom 10% | 10136 | 768 | | Reference | | |
| | Middle | 81085 | 6776 | 1.10 | 1.02 | 1.18 | 0.015 |
| | Top 10% | 10138 | 654 | 1.30 | 1.17 | 1.44 | <0.001 |
| KNN | Bottom 10% | 10136 | 730 | | Reference | | |
| | Middle | 81087 | 6633 | 1.12 | 1.04 | 1.21 | 0.004 |
| | Top 10% | 10136 | 835 | 1.31 | 1.18 | 1.44 | <0.001 |
| MARS | Bottom 10% | 10136 | 873 | | Reference | | |

| | | | | | | | |
|---|---|---|---|---|---|---|---|
| | Middle | 81087 | 6528 | 0.92 | 0.86 | 0.99 | 0.030 |
| | Top 10% | 10136 | 797 | 1.16 | 1.05 | 1.28 | 0.003 |
| MARS ensemble | Bottom 10% | 9123 | 804 | | Reference | | |
| | Middle | 72979 | 5825 | 0.97 | 0.90 | 1.04 | 0.369 |
| | Top 10% | 9123 | 762 | 1.17 | 1.06 | 1.30 | 0.002 |

*Note:* HR = hazard ratio; CI = confidence interval; SD = standard deviation. Models were adjusted for chronological age and sex. Age (in years) was used as the underlying time axis. See Panel 1 for model abbreviations.

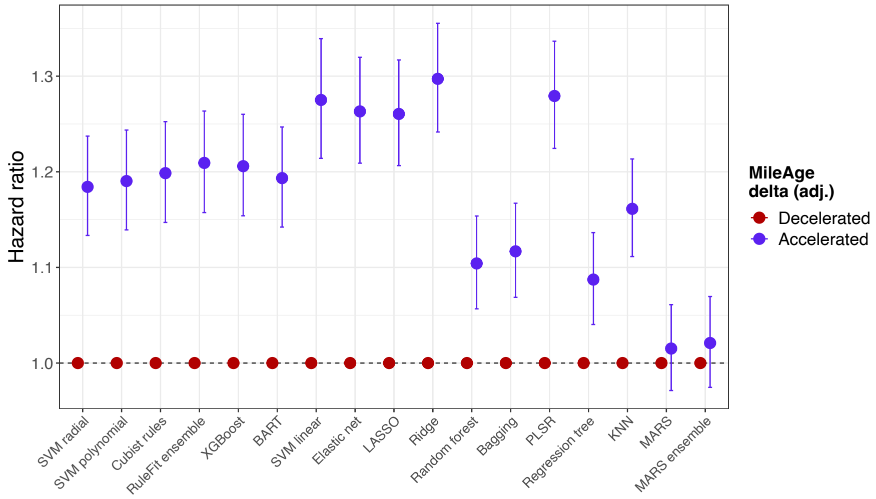

**Figure S58**. Hazard ratios (HR) and 95% confidence intervals from Cox proportional hazards models for all models. Models were adjusted for chronological age and sex. Age (in years) was used as the underlying time axis. Reference group: individuals with a negative MileAge delta (adj.), i.e., individuals with a younger predicted than true chronological age. See Panel 1 for model abbreviations.

**Table S16**. All-cause mortality by MileAge delta (adj.) – effect direction

| Model | Level | $N_{total}$ | $N_{died}$ | HR | 95% CI | | *p* |
|---|---|---|---|---|---|---|---|
| SVM radial | Decelerated | 48972 | 3709 | | Reference | | |
| | Accelerated | 52387 | 4489 | 1.18 | 1.13 | 1.24 | <0.001 |
| SVM polynomial | Decelerated | 49118 | 3718 | | Reference | | |
| | Accelerated | 52241 | 4480 | 1.19 | 1.14 | 1.24 | <0.001 |
| Cubist rules | Decelerated | 49242 | 3704 | | Reference | | |
| | Accelerated | 52117 | 4494 | 1.20 | 1.15 | 1.25 | <0.001 |
| RuleFit ensemble | Decelerated | 49343 | 3705 | | Reference | | |
| | Accelerated | 52016 | 4493 | 1.21 | 1.16 | 1.26 | <0.001 |
| XGBoost | Decelerated | 48623 | 3623 | | Reference | | |
| | Accelerated | 52736 | 4575 | 1.21 | 1.15 | 1.26 | <0.001 |
| BART | Decelerated | 49738 | 3765 | | Reference | | |
| | Accelerated | 51621 | 4433 | 1.19 | 1.14 | 1.25 | <0.001 |
| SVM linear | Decelerated | 41541 | 3025 | | Reference | | |
| | Accelerated | 39551 | 3501 | 1.28 | 1.21 | 1.34 | <0.001 |
| Elastic net | Decelerated | 51816 | 3830 | | Reference | | |
| | Accelerated | 49543 | 4368 | 1.26 | 1.21 | 1.32 | <0.001 |
| LASSO | Decelerated | 51821 | 3833 | | Reference | | |
| | Accelerated | 49538 | 4365 | 1.26 | 1.21 | 1.32 | <0.001 |
| Ridge | Decelerated | 52407 | 3821 | | Reference | | |
| | Accelerated | 48952 | 4377 | 1.30 | 1.24 | 1.36 | <0.001 |
| Random forest | Decelerated | 48858 | 3831 | | Reference | | |
| | Accelerated | 52501 | 4367 | 1.10 | 1.06 | 1.15 | <0.001 |
| Bagging | Decelerated | 47986 | 3726 | | Reference | | |
| | Accelerated | 53373 | 4472 | 1.12 | 1.07 | 1.17 | <0.001 |
| PLSR | Decelerated | 53397 | 3930 | | Reference | | |
| | Accelerated | 47962 | 4268 | 1.28 | 1.22 | 1.34 | <0.001 |
| Regression tree | Decelerated | 46221 | 3550 | | Reference | | |
| | Accelerated | 55138 | 4648 | 1.09 | 1.04 | 1.14 | <0.001 |
| KNN | Decelerated | 48566 | 3729 | | Reference | | |
| | Accelerated | 52793 | 4469 | 1.16 | 1.11 | 1.21 | <0.001 |
| MARS | Decelerated | 48232 | 4063 | | Reference | | |
| | Accelerated | 53127 | 4135 | 1.02 | 0.97 | 1.06 | 0.504 |
| MARS ensemble | Decelerated | 44397 | 3695 | | Reference | | |
| | Accelerated | 46828 | 3696 | 1.02 | 0.97 | 1.07 | 0.382 |

*Note:* HR = hazard ratio; CI = confidence interval; SD = standard deviation. Models were adjusted for chronological age and sex. Age (in years) was used as the underlying time axis. See Panel 1 for model abbreviations.

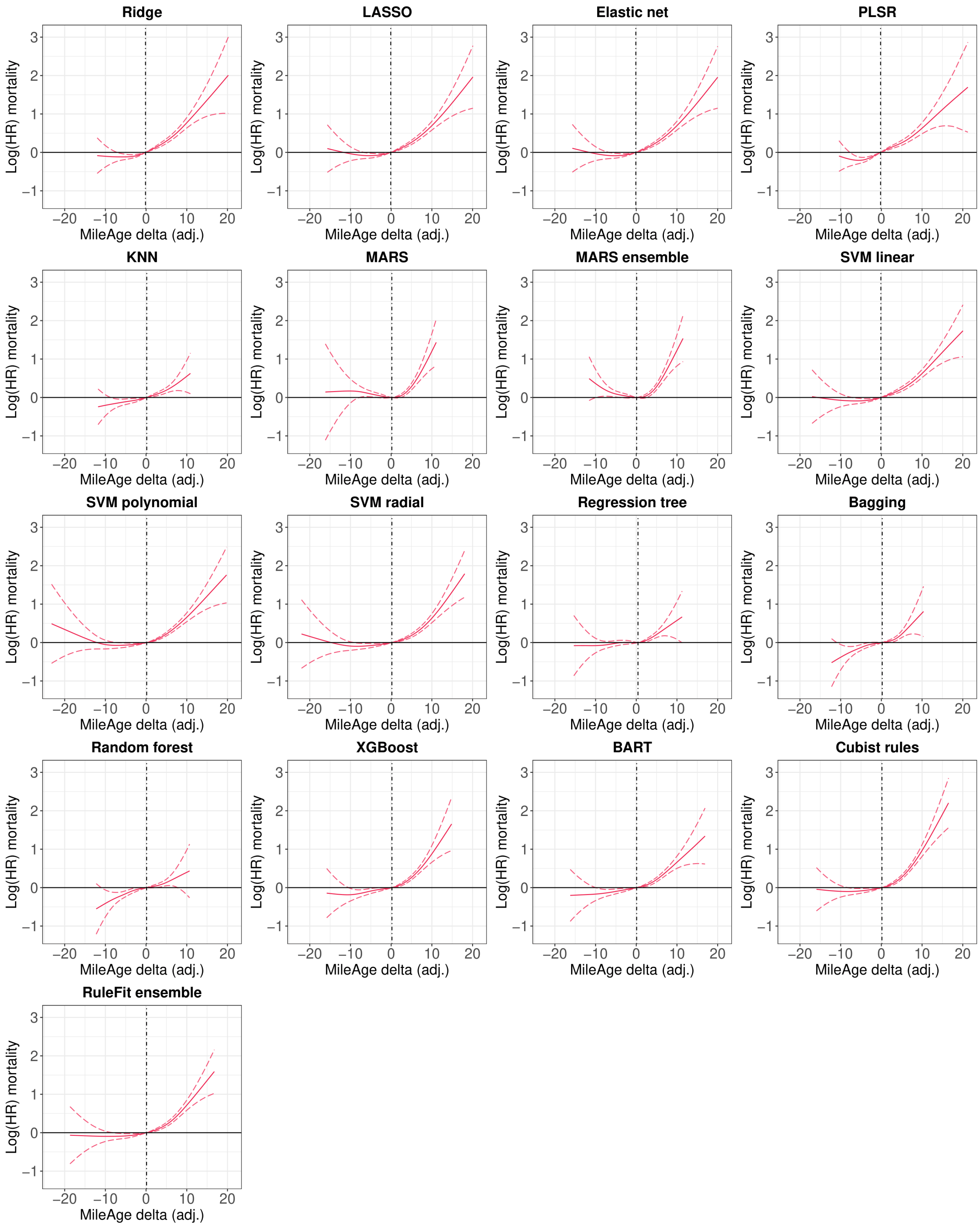

**Figure S59**. Log(HRs) and 95% confidence intervals from Cox proportional hazards models for all-cause mortality. Models were adjusted for chronological age and sex. Age (in years) was used as the underlying time axis. Vertical lines indicate the median of the distribution which represents the reference for interpreting the estimates shown. HR = hazard ratio.

### 16. MileAge delta stratified by sex

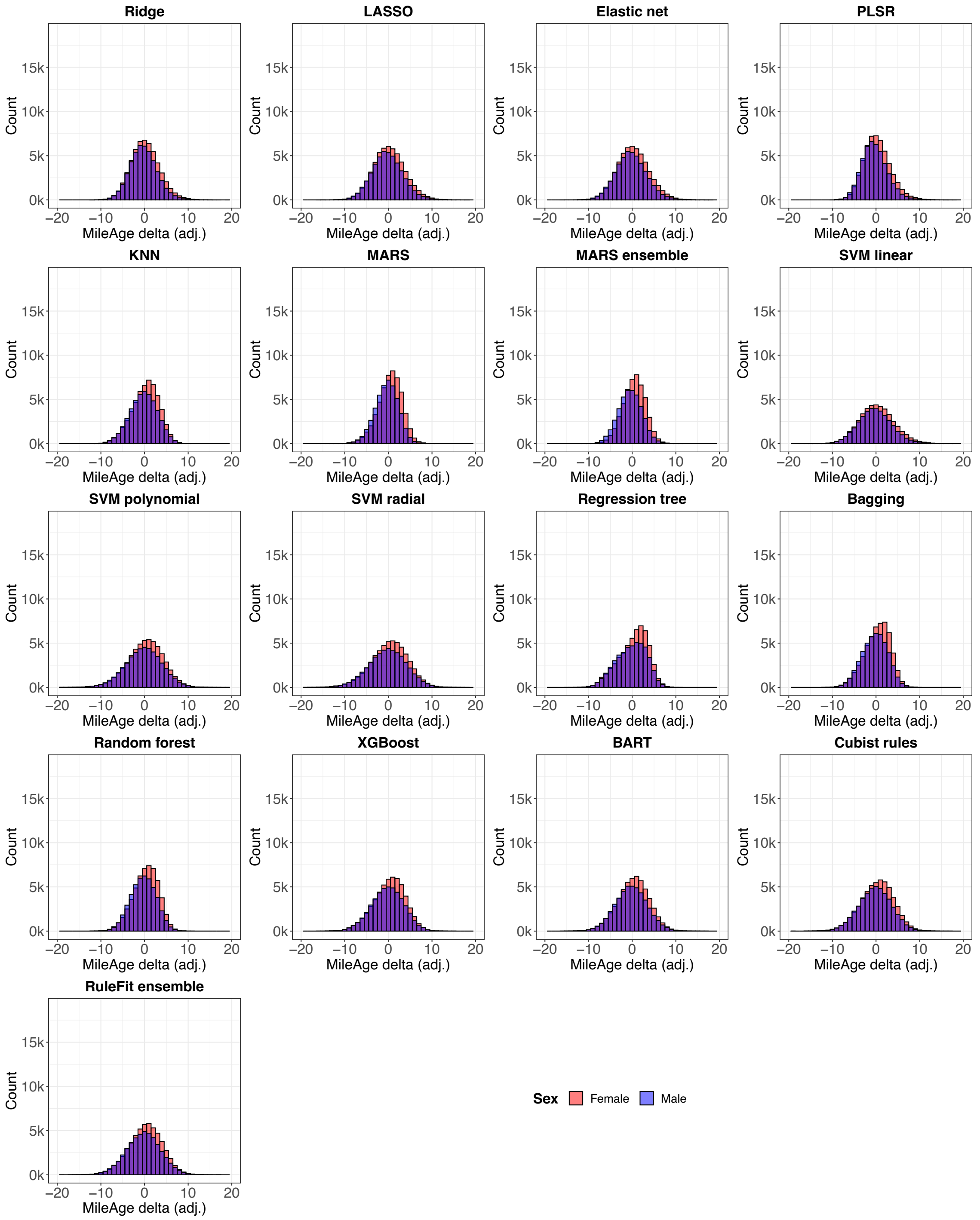

**Figure S60**. Histograms showing the distribution of MileAge delta (adj.) for all models, stratified by sex. See Panel 1 for model abbreviations.

#### 17. All-cause mortality by MileAge delta stratified

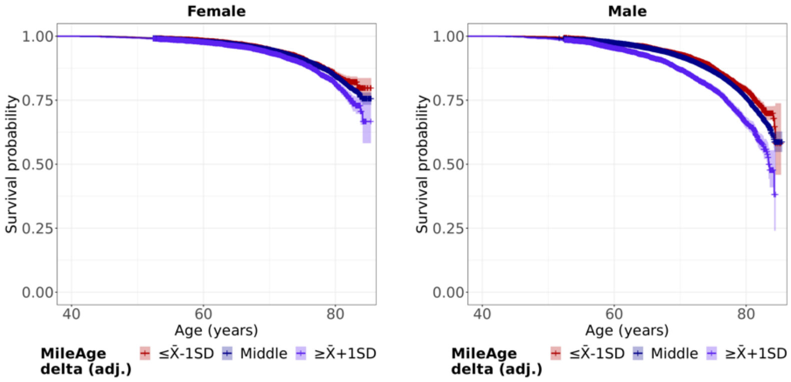

**Figure S61**. Kaplan-Meier survival probabilities for all-cause mortality by MileAge delta (adj.) derived from the Cubist rule-based regression model stratified by sex. Log-rank test *p*-values < 0.001. Age (in years) was used as the underlying time axis. Reference group: individuals with a MileAge delta (adj.) smaller than one standard deviation below the mean. SD = standard deviation.

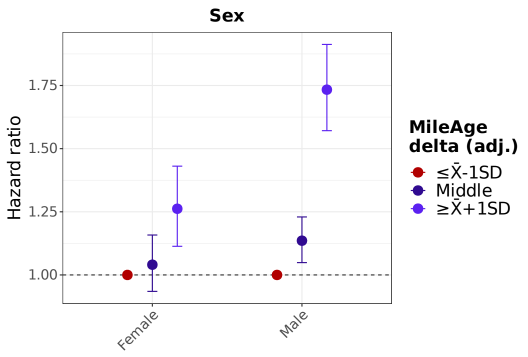

**Figure S62**. Hazard ratios (HR) and 95% confidence intervals from Cox proportional hazards models derived from the Cubist rule-based regression model stratified by sex. Models were adjusted for chronological age. Age (in years) was used as the underlying time axis. Reference group: individuals with a MileAge delta (adj.) smaller than one standard deviation below the mean. SD = standard deviation.

**Table S17**. All-cause mortality by MileAge delta (adj.) stratified

| Sex | Level | $N_{total}$ | $N_{died}$ | HR | 95% CI | | *p* |
|---|---|---|---|---|---|---|---|
| Female | ≤ X̄-1SD | 8062 | 397 | | Reference | | |
| | Middle | 37002 | 2301 | 1.04 | 0.93 | 1.16 | 0.467 |
| | ≥ X̄+1SD | 9420 | 652 | 1.26 | 1.11 | 1.43 | <0.001 |
| Male | ≤ X̄-1SD | 8142 | 763 | | Reference | | |
| | Middle | 32164 | 3224 | 1.14 | 1.05 | 1.23 | 0.002 |
| | ≥ X̄+1SD | 6569 | 861 | 1.73 | 1.57 | 1.91 | <0.001 |
| Self-rated health | | | | | | | |
| Poor | ≤ X̄-1SD | 482 | 96 | | Reference | | |
| | Middle | 2570 | 528 | 1.00 | 0.80 | 1.24 | 0.972 |
| | ≥ X̄+1SD | 1101 | 284 | 1.41 | 1.11 | 1.78 | 0.004 |
| Fair | ≤ X̄-1SD | 2909 | 329 | | Reference | | |
| | Middle | 14001 | 1606 | 1.01 | 0.89 | 1.13 | 0.913 |
| | ≥ X̄+1SD | 3950 | 507 | 1.26 | 1.10 | 1.45 | 0.001 |
| Good | ≤ X̄-1SD | 9662 | 597 | | Reference | | |
| | Middle | 40730 | 2805 | 1.10 | 1.01 | 1.20 | 0.038 |
| | ≥ X̄+1SD | 8667 | 607 | 1.29 | 1.15 | 1.45 | <0.001 |
| Excellent | ≤ X̄-1SD | 3060 | 130 | | Reference | | |
| | Middle | 11493 | 539 | 1.06 | 0.87 | 1.28 | 0.561 |
| | ≥ X̄+1SD | 2164 | 96 | 1.24 | 0.95 | 1.62 | 0.111 |
| Age group | | | | | | | |
| 39-49 years | ≤ X̄-1SD | 4406 | 90 | | Reference | | |
| | Middle | 15658 | 338 | 1.00 | 0.79 | 1.27 | 0.978 |
| | ≥ X̄+1SD | 4132 | 111 | 1.29 | 0.97 | 1.70 | 0.075 |
| 50-59 years | ≤ X̄-1SD | 5346 | 240 | | Reference | | |
| | Middle | 22689 | 1088 | 1.09 | 0.95 | 1.25 | 0.236 |
| | ≥ X̄+1SD | 5671 | 411 | 1.77 | 1.50 | 2.07 | <0.001 |
| 60-71 years | ≤ X̄-1SD | 6452 | 830 | | Reference | | |
| | Middle | 30819 | 4099 | 1.13 | 1.05 | 1.22 | 0.002 |
| | ≥ X̄+1SD | 6186 | 991 | 1.49 | 1.36 | 1.63 | <0.001 |

*Note:* HR = hazard ratio; CI = confidence interval; SD = standard deviation. MileAge delta (adj.) derived from Cubist rule-based regression model. Models were adjusted for chronological age (sex stratified), sex (age group stratified) and age and sex (self-rated health stratified). Age (in years) was used as the underlying time axis.

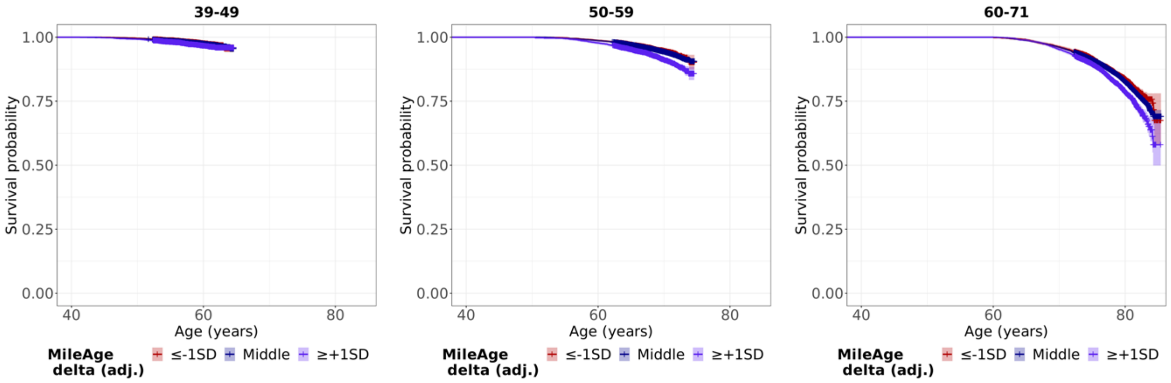

**Figure S63**. Kaplan-Meier survival probabilities for all-cause mortality by MileAge delta (adj.) derived from the Cubist rule-based regression model stratified by chronological age group. Log-rank test *p*-values: 0.058, < 0.001 and < 0.001. Age (in years) was used as the underlying time axis. Reference group: individuals with a MileAge delta (adj.) smaller than one standard deviation below the mean. SD = standard deviation.

#### 18. All-cause mortality by MileAge delta and other ageing markers

**Table S18**. All-cause mortality by MileAge delta (adj.) compared with other predictors

| Predictor | Level | $N_{total}$ | $N_{died}$ | HR | 95% CI | | *p* |
|---|---|---|---|---|---|---|---|
| MileAge delta (adj.) | $\leq \bar{X}$-1SD | 16204 | 1160 | | Reference | | |
| | Middle | 69166 | 5525 | 1.10 | 1.04 | 1.18 | 0.003 |
| | $\geq \bar{X}$+1SD | 15989 | 1513 | 1.52 | 1.41 | 1.64 | <0.001 |
| Frailty index | $\leq \bar{X}$-1SD | 14653 | 651 | | Reference | | |
| | Middle | 70990 | 5153 | 1.46 | 1.34 | 1.58 | <0.001 |
| | $\geq \bar{X}$+1SD | 15386 | 2363 | 2.90 | 2.65 | 3.16 | <0.001 |
| Grip strength | $\leq \bar{X}$-1SD | 18454 | 1256 | | Reference | | |
| | Middle | 66081 | 5344 | 1.38 | 1.29 | 1.47 | <0.001 |
| | $\geq \bar{X}$+1SD | 16460 | 1556 | 1.90 | 1.74 | 2.08 | <0.001 |
| Telomere length | $\leq \bar{X}$-1SD | 14589 | 828 | | Reference | | |
| | Middle | 68746 | 5381 | 1.08 | 1.01 | 1.17 | 0.033 |
| | $\geq \bar{X}$+1SD | 14629 | 1707 | 1.31 | 1.21 | 1.43 | <0.001 |

*Note:* HR = hazard ratio; CI = confidence interval; SD = standard deviation. MileAge delta (adj.) derived from Cubist rule-based regression model. Models were adjusted for chronological age and sex. Age (in years) was used as the underlying time axis.

**Table S19**. C-index for all-cause mortality

| Model | C-index | 95% CI | |
|---|---|---|---|
| Base model (age + sex) | 0.716 | 0.711 | 0.722 |
| + Telomere length | 0.717 | 0.711 | 0.722 |
| + MileAge delta (adj.) | 0.719 | 0.713 | 0.724 |
| + Grip strength | 0.721 | 0.716 | 0.727 |
| + Frailty index | 0.737 | 0.732 | 0.742 |

*Note:* CI = confidence interval. MileAge (adj.) derived from Cubist rule-based regression model. Age (in years) was used as the underlying time axis.
